# Integrating GWAS and Transcriptomic Data Using PrediXcan and Multimodal Deep Learning Reveals Genetic Basis and Drug Repositioning Opportunities for Alzheimer’s Disease

**DOI:** 10.1101/2025.01.02.25319880

**Authors:** Xuecong Tian, Ying Su, Sizhe Zhang, Chen Chen, Yaolei Ma, Haiqing Sun, Cheng Chen, Wei Zhou, Yue Gao, Luyu Zhou, Xiaoyi Lv, Panos Roussos, Wen Zhang

## Abstract

Alzheimer’s disease (AD), the leading cause of dementia, imposes a significant societal and economic burden; however, its complex molecular mechanisms remain unclear. This study integrates multi-omics data with advanced artificial intelligence (AI) methods to uncover the molecular basis underlying AD phenotype regulation and explore personalized drug repositioning strategies based on individual genetic backgrounds. First, we applied the PrediXcan method to identify candidate genes closely associated with AD cognitive diagnosis, selecting from 61 brain-related traits. We validated these findings through individual-level analysis using gene expression and genotype data from 553 dorsolateral prefrontal cortex samples in the ROSMAP database. Simultaneously, we constructed a deep, multi-layer information fusion model (AD-MIF) by integrating genotype and gene expression data and employing autoencoders as well as graph autoencoders for multi-modal feature extraction. The results revealed a 10–20% improvement in the Area Under the Curve (AUC) for predicting AD-related phenotypes. Both approaches showed high consistency across cellular structures, brain regions, and neurobiological pathways, demonstrating their complementary advantages. Gene enrichment analysis indicated that APOE and its interacting gene APOC1 play a central role in cholesterol metabolism, lipid transport, and immune regulation, while genes such as SCIMP and KAT8 are involved in immune signaling, epigenetic regulation, and neuroprotection. After incorporating attention mechanisms, AD-MIF highlighted the importance of key genes, such as POLR2C and TRAPPC4, in regulating neuronal function. Based on predictive results and enrichment analysis, we further identified candidate drugs, including sirolimus, dasatinib, and MGCD-265. In vivo experiments confirmed that MGCD-265, also known as Glesatinib, and dasatinib significantly improve cognitive deficits in the SAMP8 AD model mice by inhibiting neuroinflammation, pathological tau phosphorylation, and Aβ deposition. This study demonstrates the complementary advantages of bioinformatics pipelines and AI-based multi-modal fusion methods in elucidating the complex pathological mechanisms of AD and enhancing phenotype prediction accuracy. It also provides new theoretical support for personalized drug interventions based on individual genetic characteristics, laying a solid foundation for optimizing early screening, prediction, and personalized treatment strategies.

**Graphical Abstract:** 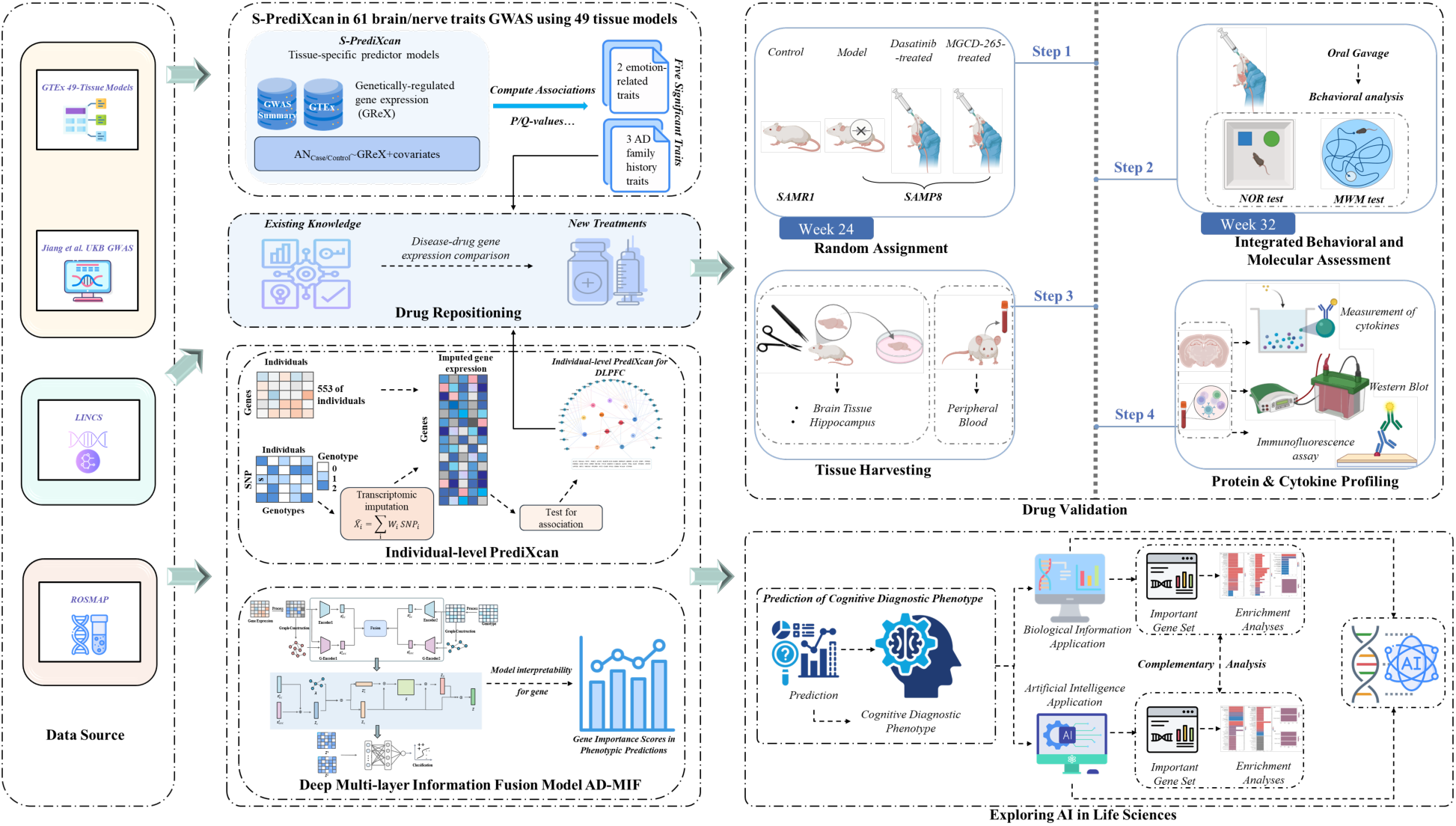

## Main

Alzheimer’s disease (AD) is the leading cause of dementia, which is rapidly emerging as one of the most costly, deadly, and burdensome diseases of the 21st century [1, 2]. This neurodegenerative disorder impairs cognitive functions such as memory, behavior, and emotions, eventually leading to the loss of the ability to perform daily activities and engage in social interactions [3]. AD primarily affects the cerebral cortex and hippocampal regions, with pathological features including the accumulation of insoluble amyloid-beta (Aβ) plaques and tau protein aggregation within neurons [4]. Currently, approximately 6.9 million Americans aged 65 and older are affected by AD, with medical and caregiving costs projected to reach $360 billion by 2024 [5]. To alleviate the substantial burden of AD, it is crucial to explore its etiology and identify potential therapeutic drugs.

Genome-wide association studies (GWAS) are experimental designs used to detect associations between genetic variations and traits, with results dependent on factors such as sample size and allele frequency [6]. To date, GWAS has identified approximately 70 genetic loci associated with AD risk, providing crucial insights into the genetic mechanisms underlying AD [7]. However, the interpretation of GWAS loci remains challenging, and it is difficult to precisely identify functional genes and regulatory mechanisms [8]. Transcriptome-wide association studies (TWAS) integrate GWAS data with gene expression changes, using reference gene expression data to predict the expression levels of disease-related genes. This approach identifies genes associated with traits across the genome and reveals potential biological mechanisms [9]. Recently, Parrish et al. developed a new stacked regression transcriptome-wide association analysis (SR-TWAS) method using data from the GTEx project (Version 8). By utilizing multi-tissue gene expression prediction models, they identified six AD-related genes in the motor cortex and nine Parkinson’s disease (PD)-related genes in the substantia nigra, providing new insights into the genetic mechanisms of both diseases [10]. Guo et al. applied BGW-TWAS and aggregated Cauchy testing methods to integrate cis- and trans-eQTL data from brain and blood tissues with GWAS summary data, identifying 141 TWAS risk genes for AD dementia [11]. Mews et al. used the OTTERS TWAS pipeline, combining cis-eQTL data from cortical brain tissue (MetaBrain) and blood (eQTLGen), to discover and validate five novel AD gene associations (PRKAG1, C3orf62, LYSMD4, ZNF439, SLC11A2) and six additional genes near known GWAS loci [12]. TWAS, by utilizing multi-tissue data analysis, provides a novel perspective for understanding the functional implications of AD-associated variants and further expands our understanding of the genetic mechanisms of the disease. However, while TWAS can reveal associations between gene expression and disease risk, it cannot directly prove causality [9].

To further explore the biological mechanisms of diseases, Barbeira et al. developed the PrediXcan method, which combines SNP gene expression prediction models with GWAS data to infer gene expression levels and identify genes associated with diseases [13]. Yanfa Sun et al. proposed the UTMOST strategy, based on the S-PrediXcan method, which integrates multi-tissue gene expression data to identify 53 genes associated with AD risk [8]. PrediXcan not only overcomes the limitations of traditional GWAS but also reveals the mediating role of gene expression in phenotypes. As a widely used individual genotype-to-transcriptome inference method, PrediXcan estimates gene expression values determined by genetic traits and explores the potential mechanisms underlying these associations by testing the relationship between predicted values and target phenotypes [14]. Furthermore, integrating multi-omics data, such as gene expression and genotype, with artificial intelligence (AI) methods can capture complex nonlinear relationships, aiding in the understanding of disease mechanisms [15]. For example, the GenNet framework [16] developed by Hilten et al. and the DeepGAMI model [17] proposed by Chandrashekar et al. have both demonstrated outstanding predictive performance as well as the ability to interpret underlying mechanisms in disease research. By integrating gene expression and genotype data using deep learning models, these approaches show great promise for improving the AD risk prediction, providing new directions for early diagnosis and the identification of potential therapeutic targets.

Due to the lack of effective therapeutic drugs for AD and the high costs and lengthy development cycles associated with traditional drug discovery, coupled with an unclear disease mechanism [18, 19], drug repositioning has emerged as a strategy to accelerate therapeutic development [20]. The risk of many complex traits is driven by the accumulation of variations with relatively low significance, which presents a challenge for drug repositioning [21]. To address this, Hon-Cheong So et al. proposed a novel strategy that utilizes GWAS data to infer gene expression profiles and compares these profiles with drug-induced gene expression changes [22]. This method overcomes the confounding effects of drugs and environmental factors in traditional gene expression studies and has shown progress in research on complex diseases such as AD.

Based on the aforementioned research, this study utilized GWAS summary statistics provided by Jiang et al. [23], along with gene expression prediction models from 49 tissues in the GTEx database. Using the S-PrediXcan method, we conducted a comprehensive TWAS on 61 brain-related traits, ultimately identifying five significant traits, three of which were related to a family history of AD. Additionally, we employed gene expression and genotype data from 553 individuals in the ROSMAP database, focusing on the dorsolateral prefrontal cortex. By applying the individual-level PrediXcan method, we correlated predicted gene expression levels with AD-related phenotypes to further validate the reliability of these significant traits. Notably, this study introduced an innovative deep multi-layer information fusion model, AD-MIF, designed to leverage high-quality multimodal data (such as gene expression and gene phenotypes) for the accurate prediction of a range of important phenotypes. Using deep learning techniques, AD-MIF enables comprehensive integration of multimodal data, extracting more representative features to significantly enhance prediction accuracy. Based on core AD regulatory genes identified in our research, we integrated the strategy proposed by Hon-Cheong So et al. to compare GWAS-inferred gene expression profiles with drug-induced gene expression changes, identifying a series of potential candidate drugs. To further validate our computational results, we employed a comprehensive set of behavioral, molecular, and histopathological assessments in the SAMP8 AD mouse model to evaluate the selected candidate drugs. In vivo results demonstrated that treatment with MGCD-265 and dasatinib significantly improved cognitive abilities and notably reduced neuroinflammation. Moreover, both compounds effectively alleviated key neuropathological changes, significantly reducing tau hyperphosphorylation and amyloid-beta deposition. These findings provide strong experimental support for the therapeutic potential of MGCD-265 and dasatinib in AD. This study not only offers new perspectives for early diagnosis and intervention in AD but also paves the way for future research, advancing the field.

## Results

### S-PrediXcan Analysis of AD Family History and Related Traits

#### Gene Expression and Significant Gene Count for AD Family History-Related Traits

A TWAS was conducted using the S-PrediXcan method to analyze gene expression levels across 49 different tissues for 61 traits related to brain and mental health (Supplementary Table S1). Supplementary Figure S1a illustrates that, after adjusting for q-values and applying a false discovery rate (FDR) threshold of 0.01, 18 significant traits were identified. The number of significant genes in different tissues was visualized (see Figure 1a). In the figure, red and blue represent high and low counts of significant genes, respectively, revealing the pleiotropy of gene expression across various tissues. Notably, traits such as GCST90042665: Father’s Disease – AD/Dementia, GCST90042678: Mother’s Disease – AD/Dementia, GCST90042691: Sibling’s Disease – AD/Dementia, GCST90041866: Irritability, and GCST90041879: Worry/Anxiety exhibit a relatively rich distribution of significant genes.

**Figure 1.**
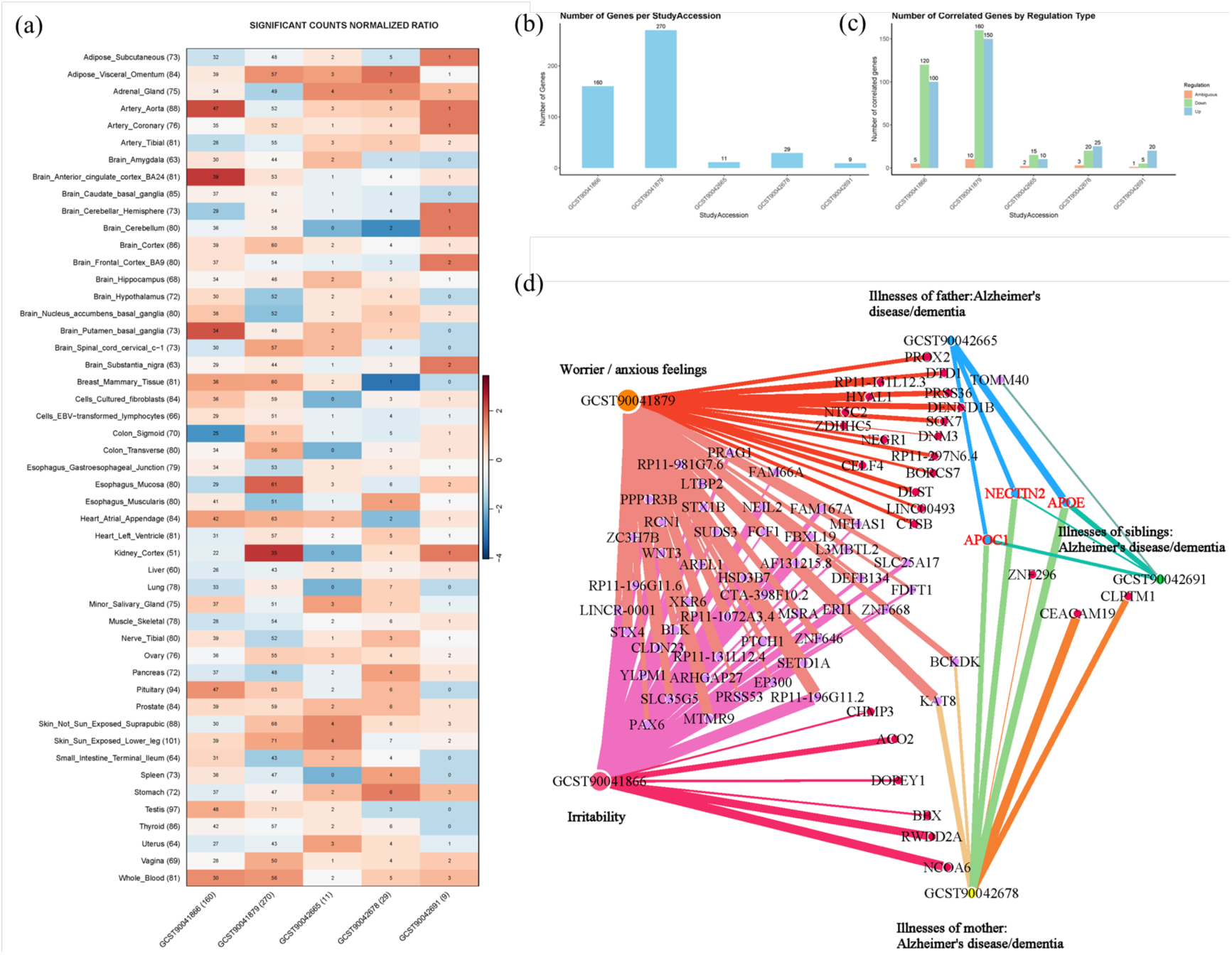
A Comprehensive Overview of Significant Genes Associated with Brain and Psychiatric Traits: Multi-Tissue Expression Distribution, Regulatory States, and Interaction Network Analysis. **a) Distribution of Significant Genes for Five Brain and Psychiatric Traits (Including AD Family History and Emotional Traits) Across 49 Tissues Identified via S-PrediXcan Analysis.** The heatmap illustrates the distribution of significant genes for five brain- and psychiatric-related traits, selected from an initial analysis of 61 traits, using S-PrediXcan across 49 different tissues. These traits include: GCST90042665 (Father’s Disease: Alzheimer’s/Dementia), GCST90042678 (Mother’s Disease: Alzheimer’s/Dementia), GCST90042691 (Sibling’s Disease: Alzheimer’s/Dementia), GCST90041866 (Irritability), and GCST90041879 (Worry/Anxiety). Rows represent different tissues, while columns represent these five traits. The colors indicate standardized significant gene counts (Z-scores), with red representing higher gene counts and blue representing lower counts. The results highlight the pleiotropy of these traits and their tissue-specific associations, particularly in brain regions such as the hippocampus and frontal cortex, where significant gene concentrations are notably higher. **b) Number of significant genes in each trait.** This figure displays the total number of significant genes linked to each AD-related trait. The significant genes for each trait are determined based on gene expression differences across multiple tissues. Here, the count represents the number of unique significant genes per trait, meaning each gene is counted only once. **c) The Number of Genes Whose Gene Expression is Up-regulated, Down-regulated and Ambiguous in Each Trait.** This figure illustrates the number of genes with up-regulated, down-regulated, and ambiguous (i.e., equal numbers of up-regulated and down-regulated) gene expressions for each AD-related trait. The statistics are based on the performance of each gene across multiple tissues. In some cases, a gene may exhibit different expression statuses (up-regulated or down-regulated) in different tissues, resulting in a single gene being counted in multiple categories (up-regulated or down-regulated). **d) Gene Expression Network for Brain- and Psychiatric-Related Traits at a q-value Threshold of 0.05.** This figure illustrates the core genes with the strongest associations identified at a q-value (pva.qval) threshold of 0.05. These genes demonstrate the highest significance in traits related to familial AD/dementia history, highlighting their critical roles in the genetic mechanisms underlying AD family history.

Among traits related to a family history of AD, key brain regions such as the hippocampus (Brain_Hippocampus) and the frontal cortex (Brain_Frontal_Cortex_BA9) demonstrated significant pleiotropy (Supplementary Figure S1b). This suggests that these brain regions may play critical roles in the onset and progression of AD. Other brain regions, including the anterior cingulate cortex (Brain_Anterior_Cingulate_Cortex_BA24) and the cerebral cortex (Brain_Cortex), showed strong gene associations with emotional traits (e.g., irritability and worry/anxiety), indicating that these areas may be closely related to the emotional and behavioral regulation mechanisms associated with AD. It is noteworthy that the distribution of significant genes is not limited to the brain; peripheral tissues such as the liver (Liver), lung (Lung), and arteries (Artery_Aorta, Artery_Coronary) also exhibited a number of significant genes. Although the number of significant genes is lower compared to brain tissues, these peripheral tissues still show associations with traits related to a family history of AD (Figure 1b and Figure 1c). This suggests that AD is not solely a neurodegenerative disease; its development may also involve pathological changes in the metabolic and vascular systems, particularly in cases with a familial genetic background.

Overall, significant differences were observed in the number of significant genes across different traits within the same tissue. For example, the hippocampus and frontal cortex exhibit a high number of significant genes across multiple AD family history traits and emotional traits, suggesting that these brain regions play a central role in the various genetic mechanisms of AD. In contrast, although peripheral tissues contain fewer significant genes, their association with familial AD suggests that these tissues may play important roles in the disease through metabolic or vascular-related pathways.

In summary, the number of significant genes in brain tissues is markedly higher than in non-brain tissues, which aligns with AD being characterized as a neurodegenerative disease. However, the observed significance in peripheral tissues suggests that AD may involve more extensive systemic involvement, particularly with potential key roles in metabolic, inflammatory, and vascular pathological processes. Therefore, exploring the synergistic effects between brain and peripheral tissues may provide new insights into the systemic mechanisms of AD.

#### Gene Expression and Enrichment Analysis for AD Family History-Related Traits

In this study, a comprehensive analysis of gene expression was conducted for five traits related to a family history of AD using the S-PrediXcan method. Three screening analyses were performed with different q-value thresholds (q-value ≤ 0.05, q-value ≤ 0.1, and q-value ≤ 0.5), and gene network diagrams were constructed. As the q-value threshold was lowered, the selected genes became increasingly concentrated around a few core genes strongly associated with AD. At a q-value ≤ 0.5, the gene distribution was relatively broad, encompassing multiple traits. However, at q-values ≤ 0.1 and ≤ 0.05, the results progressively focused on key genes with strong associations. Detailed information is provided in Supplementary Tables S2–S4.

#### Gene Expression Networks at q-value ≤ 0.5

In the gene expression network analysis with a q-value ≤ 0.5, shared genes were identified across multiple traits. These genes are not only closely associated with familial AD but also exhibit significant associations with emotional and behavioral characteristics, such as irritability and worry/anxiety. These shared genes reveal complex genetic links between AD family history traits and emotional and behavioral features, indicating their important roles in polygenic regulatory processes. Supplementary Figure S1c illustrates the distribution of significantly associated genes across multiple traits. APOE is a core gene closely related to AD risk, showing strong associations with three family history traits (father’s, mother’s, and siblings’ history of AD/dementia), emphasizing its importance in the genetic mechanisms of AD. APOC1 is also related to all three family history traits of AD, while SCIMP is associated with two family histories of AD and irritability, suggesting that these genes may play roles in AD progression and emotion regulation. KAT8 is significantly associated with AD family history, irritability, and anxiety traits, indicating that it may have dual functions in emotion regulation and AD genetic mechanisms. Other genes, such as RP11-196G11.6, VKORC1, RP11-196G11.2, PRSS53, and ZNF646, also show associations with familial AD history, irritability, and anxiety traits, further suggesting their potential importance in hereditary AD and emotion regulation. Detailed networks are presented in Supplementary Figure S1d.

#### Gene Expression Networks at q-value ≤ 0.1

When the q-value threshold is set to ≤ 0.1, the number of significant genes identified decreases, but the associations of these genes become stronger. In this analysis, core genes that are significantly associated with multiple familial AD history traits were identified, such as APOE, NECTIN2, and TOMM40. The gene network becomes more concentrated under this threshold, particularly in the traits of irritability and worry/anxiety, with a significant increase in the number of shared genes. Supplementary Figure S1e shows that these core genes are widely present across multiple familial AD history traits, especially those related to AD-associated emotional and cognitive symptoms. TOMM40 and APOE show strong associations across all three family history traits, further confirming their central roles in the genetic mechanisms of AD. Other genes, such as RP11-196G11.6, ZNF646, and ZNF668, exhibit shared associations between familial AD history and emotional traits (e.g., irritability, anxiety), suggesting that these genes may play roles in the genetic mechanisms through which emotional and behavioral traits influence AD.

#### Gene Expression Networks at q-value ≤ 0.05

At a q-value threshold of ≤ 0.05, the number of genes identified is significantly reduced. At this threshold, many genes are related to irritability and anxious feelings. Among these, BCKDK and KAT8 link anxious feelings traits to maternal AD family history traits, suggesting a connection between family history traits and irritability/anxiety traits (Figure 1d). APOE and NECTIN2 show the strongest associations under this threshold, especially exhibiting higher significance in traits related to familial AD/dementia history. These genes are widely involved in the core genetic mechanisms of familial AD. Figure 1d illustrates that the most strongly associated genes are concentrated in a few key traits, particularly showing pronounced significance in traits such as the father’s disease – AD/dementia and the mother’s disease – AD/dementia, revealing core genetic loci associated with AD family history. APOE and NECTIN2 continue to show strong associations with multiple familial AD history traits, while APOC1 also exhibits high significance across multiple AD-related traits, further indicating its crucial role in familial AD. Detailed information is available in Supplementary Table S5.

#### GO and KEGG Enrichment Analysis of AD-Related Genes in Significant Tissues

Based on a p-value and q-value threshold of ≤ 0.05, GO and KEGG enrichment analyses were performed on genes associated with familial AD traits in tissues such as the aorta (Artery_Aorta), anterior cingulate cortex BA24 (Brain_Anterior_cingulate_cortex_BA24), putamen/basal ganglia (Brain_Putamen_basal_ganglia), esophageal mucosa (Esophagus_Mucosa), and kidney cortex (Kidney_Cortex). The results of GO enrichment (see Figure 2a) showed that gene expression related to familial AD involved changes in many cellular biological processes, cell components, and molecular functions. These changes spanned immune response, lipid metabolism, amyloid deposition, cell stress response, and other key processes. The enrichment of natural killer cells and tumor immune response pathways may reveal immune system changes in AD. Immune response could play a dual role in the occurrence of neurodegenerative diseases, both providing disease resistance and contributing to neuroinflammation. The combination of amyloid β-protein and its related proteins, along with the enrichment of the amyloid aggregation regulation pathway, indicates that the pathological characteristics of AD are primarily exacerbated by the accumulation of amyloid and the dysfunction of intracellular clearance systems. KEGG enrichment analysis results (see Figure 2c) identified a series of pathways related to metabolism, cell adhesion, sugar biosynthesis, and transmembrane transport. The enrichment of cholesterol metabolism suggests that lipid metabolism disorders in AD may worsen amyloid protein deposition. Cell adhesion molecules and glycosylation may regulate nerve cell function by affecting adhesion and signal transmission between cells, as well as influencing neurotransmitter release and conduction through receptor glycosylation and synaptic plasticity. Dysregulated mechanisms may impair signal transmission between neurons, further aggravating neurotransmitter imbalances and cognitive decline. The enrichment of ABC transporters highlights the critical role of lipid and metabolite transport in the cell membrane, which may be a key factor in cholesterol metabolism disorder.

**Figure 2.**
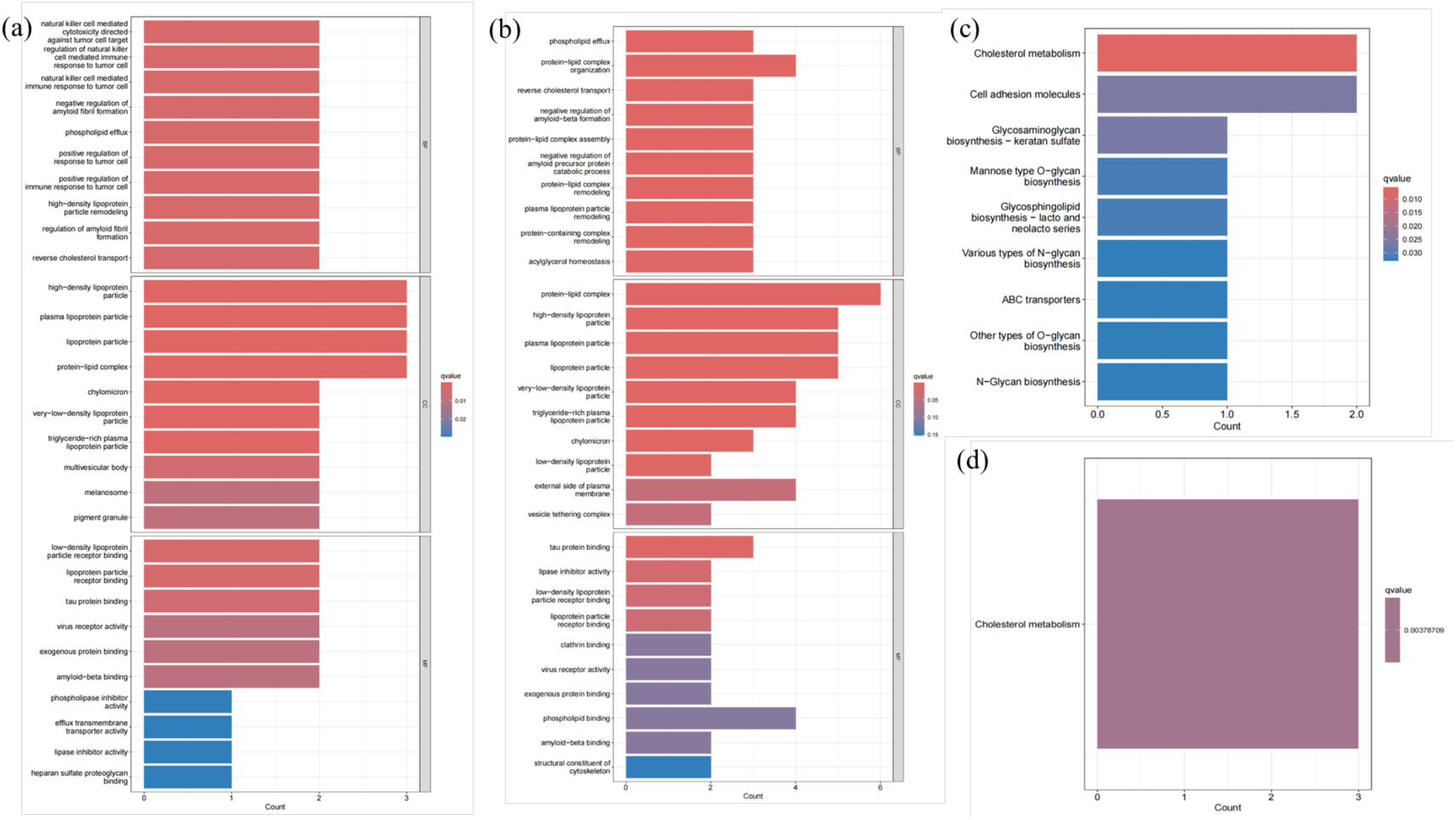
GO Biological Process and KEGG Pathway Enrichment Analysis Across Significant Tissues (p-value and q-value ≤ 0.05). **a) GO biological process enrichment analysis in key tissues of interest. b) GO biological process enrichment analysis in other significant tissues. c) KEGG pathway enrichment analysis in key tissues of interest. d) GO biological process enrichment analysis in other significant tissues.**

#### GO and KEGG Enrichment Analysis of AD-Related Genes in Other Significant Tissues

For tissues outside the primary focus, GO and KEGG enrichment analyses were also conducted based on p-value and q-value ≤ 0.05. The GO enrichment analysis (see Figure 2b) in other important tissues is consistent with the enrichment results from the aforementioned tissues, including lipid metabolism disorders, abnormalities of amyloid and tau proteins, changes in cell membrane function, and immune response regulation. These findings confirm the critical role of lipid metabolism, amyloid β-protein accumulation, and tau protein aggregation in multiple tissues of AD, suggesting that their interactions may play a central role in nerve cell injury and cognitive decline. This is consistent with existing research. The KEGG pathway analysis (see Figure 2d) indicates that cholesterol metabolism remains the most significantly enriched pathway in non-primary tissues. Although these tissues have received less research attention, their lipid metabolic activity may still hold potential significance in the pathological mechanisms of AD.

Using the screening criteria of p-value and q-value ≤ 0.05, functional differences in AD-related genes across various tissues were uncovered. Key significant tissues play central roles in immune responses and lipid metabolism, while other tissues contribute to auxiliary lipid metabolic regulation. These findings offer a deeper understanding of the complex genetic background and multiple pathological mechanisms of AD.

#### GO and KEGG Enrichment Analysis of AD-Related Genes in the Anterior Cingulate Cortex BA24

To further explore the potential functions of genes associated with familial AD traits in the anterior cingulate cortex (Brain Anterior Cingulate Cortex, BA24), GO and KEGG enrichment analyses were conducted on the selected key genes. The anterior cingulate cortex plays a crucial role in cognitive function, emotion regulation, and the pathogenesis of AD. Using a screening criterion of p-value ≤ 0.01, multiple biological processes and pathways related to AD pathology were identified.

The GO analysis results (see Figure 3a) show that the relevant genes are mainly enriched in processes such as pigment organization, cell adhesion, detoxification, and natural killer cell-mediated immune responses. Specifically, the APOE and ABCB6 genes are closely related to the formation and metabolism of pigment granules, while the CEACAM19 and PVR genes play key roles in intercellular adhesion and immune functions. Additionally, the enrichment of the APOE gene in processes related to cellular responses to toxic substances further supports its multifaceted role in the pathogenesis of AD. KEGG pathway analysis (see Figure 3c) highlights the central roles of cholesterol metabolism and ABC transporter proteins in AD. The significant enrichment of the APOE gene in the cholesterol metabolism pathway indicates its critical role in regulating lipid metabolism and amyloid protein deposition. The ABCB6 gene is enriched in the ABC transporter protein pathway, potentially affecting AD pathology by regulating the transport of substances in and out of cells. These results suggest that AD-related genes in the anterior cingulate cortex play important roles in lipid metabolism, immune responses, cell adhesion, and detoxification processes, further elucidating their key roles in the complex pathological mechanisms of AD.

**Figure 3.**
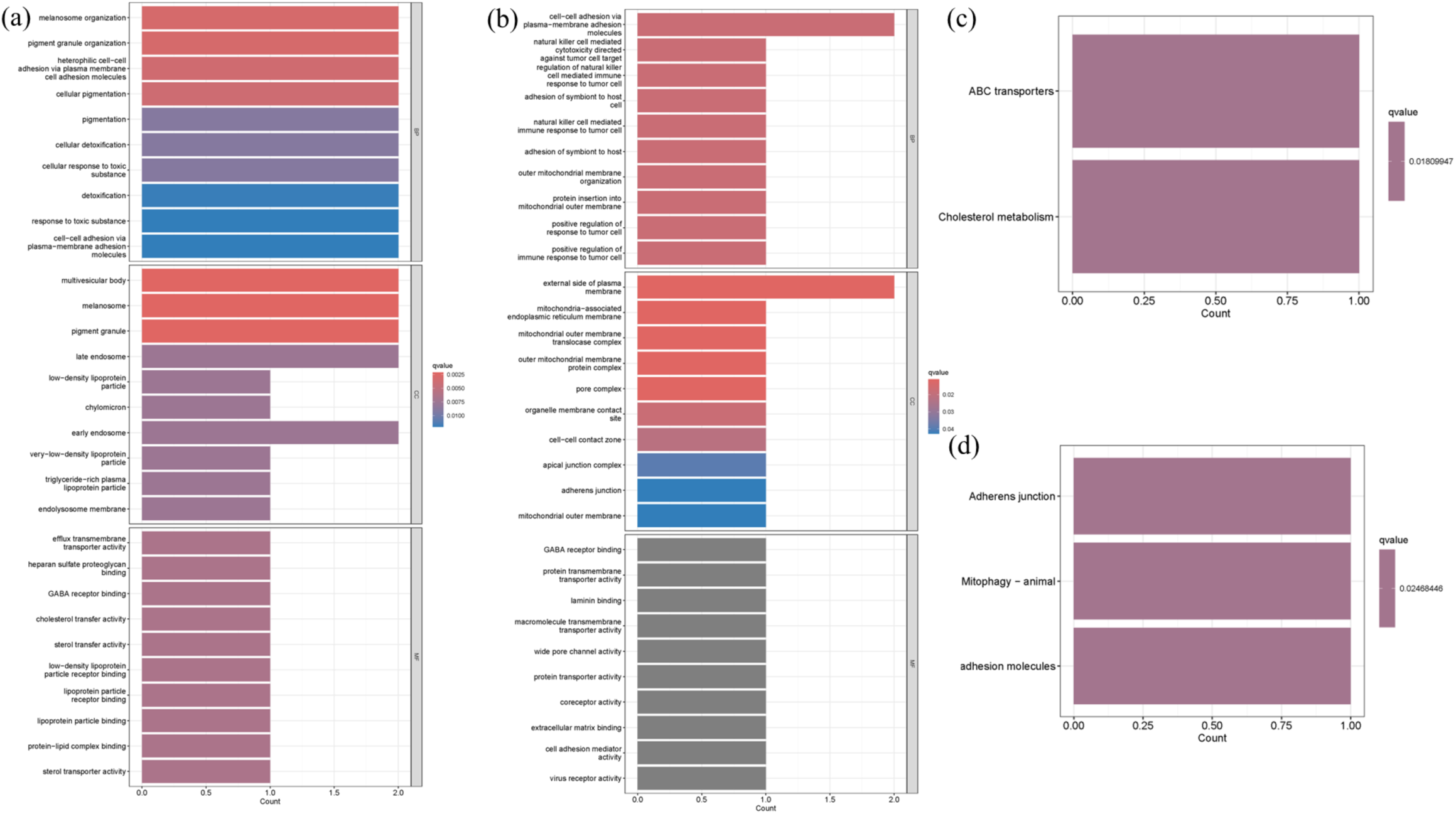
GO Biological Process and KEGG Pathway Enrichment Analysis in Brain Anterior Cingulate Cortex (BA24) and Artery Aorta (q-value ≤ 0.01). **a) GO Biological Process Enrichment Analysis in Brain Anterior Cingulate Cortex (BA24). b) GO Biological Process Enrichment Analysis in Artery Aorta. c) KEGG Pathway Enrichment Analysis in Brain Anterior Cingulate Cortex (BA24). d) KEGG Pathway Enrichment Analysis in Artery Aorta.**

#### GO and KEGG Enrichment Analysis of AD-Related Genes in the Aorta

To investigate the potential functions of genes associated with familial AD in peripheral tissues, we selected the aorta (Artery Aorta) for GO and KEGG enrichment analyses. This analysis explored the roles of these genes outside of brain tissues. The results revealed several important biological processes and signaling pathways related to cell adhesion, immune responses, and mitochondrial function, using a screening criterion of p-value ≤ 0.01. These findings indicate that these processes are significantly enriched in the aorta, suggesting that the peripheral vascular system may play a significant role in the pathological mechanisms of AD.

The GO analysis (see Figure 3b) shows that genes such as NECTIN2 and CEACAM19 are closely related to cell–cell adhesion, which could affect AD pathology by maintaining vascular structure and function. Additionally, NECTIN2 is enriched in biological processes related to natural killer cell-mediated immune responses, highlighting its critical role in immune dysfunction associated with AD. The TOMM40 gene is involved in mitochondrial outer membrane organization and the insertion of proteins into the mitochondrial outer membrane, suggesting that mitochondrial function may be crucial in the energy metabolism dysregulation observed in AD. KEGG analysis (see Figure 3d) further supports these findings, showing that the NECTIN2 gene is enriched in adhesion junctions and cell adhesion molecule pathways, underscoring its crucial role in maintaining vascular wall stability and regulating cell–cell interactions. Additionally, the mitochondrial autophagy pathway of TOMM40 is enriched, implying its significant role in aortic cell homeostasis and AD-related metabolic abnormalities. Overall, these results indicate that AD-related genes in the aorta may have a substantial impact on the pathological progression of AD by regulating cell adhesion, immune responses, and mitochondrial function.

### Individual-level PrediXcan Analysis of the ROSMAP Dataset and APOE Association Results

#### Gene Expression Network Analysis of AD-Related Phenotypes

A shared gene expression network analysis was conducted across nine phenotypes related to AD (phenotype annotation information is provided in Supplementary Table S6). Among these, the APOE genotype is strongly associated with the genetic risk of AD and, as a key genetic marker, its impact on AD pathology has garnered significant attention. As shown in Supplementary Figure S2a, the shared network analysis of gene expression across the phenotypes revealed several key genes shared among multiple phenotypes. Notably, the genes NDUFA13, C17orf58, and BTG2 were identified as shared across multiple AD phenotypes, including age at death, MMSE scores at the last follow-up, and Braak pathological staging. The presence of these genes suggests that they may play multiple roles at different stages of AD progression (see Supplementary Table S7).

For instance, NDUFA13 is essential for mitochondrial function and energy metabolism, processes that are likely linked to the metabolic dysregulation observed in AD. C17orf58 and BTG2 are involved in the regulation of cell proliferation and apoptosis, suggesting that they may play critical roles in the survival and death processes of neural cells. The shared nature of these genes underscores their central roles in regulating multiple AD phenotypes and provides valuable insights for further functional validation.

#### Functional Enrichment Analysis of Phenotype-related Genes in Cognitive Diagnosis

To further investigate cognitive diagnostic scores closely associated with the risk of AD, enrichment analyses were conducted on related genes. These analyses revealed the significant functions of multiple genes across different cellular locations, tissues, and subcellular structures, particularly highlighting their potential roles in cognitive function and the nervous system.

Through Gene Ontology (GO) analysis (Supplementary Figure S2b), several GO terms, such as “intracellular anatomical structure,” “membrane-bound organelle,” and “cytoplasm,” exhibited significant enrichment of genes (e.g., MTMR7, PPP1R13B, UQCRC1) that play critical roles in fundamental biological processes such as neural signal transduction, metabolism, and protein synthesis. These genes may be involved in the cognitive regulation of the nervous system by modulating cellular structures and functions. Additionally, Tissue Expression analysis revealed that several genes are highly expressed in key tissues, including the brain, nervous system, and endocrine glands, further suggesting their close association with neural development, neuroprotection, and cognitive function. Subcellular Localization analysis further demonstrated that these enriched genes are widely distributed across various intracellular regions, particularly in the cytoplasm and organelles, and are involved in key processes such as metabolism, protein synthesis, and cell signal transduction. These processes are crucial for neuronal function, synaptic plasticity, and cognitive health. UniProt Keywords analysis revealed post-translational modifications of these genes, including splice variants, phosphorylation, and acetylation, which may play important roles in synaptic plasticity and neural signal regulation by modulating protein stability and activity (Supplementary Table S8).

In summary, the individual-level PrediXcan analysis, combined with enrichment results, identified key genes shared across multiple phenotypes, particularly emphasizing the core roles of NDUFA13, C17orf58, and BTG2. Furthermore, the significant roles of the enriched genes in various cellular functions and processes suggest that they may influence cognitive functions of the nervous system through multiple mechanisms. These genes not only serve as potential biomarkers but may also act as therapeutic targets for AD.

### Performance Evaluation of Comparison Model AD-MIF

In this study, the multimodal AD gene data were randomly divided into training sets (80%), validation sets (10%), and test sets (10%). These subsets were used for training and evaluating the AD-MIF model. To assess the predictive performance of AD-MIF on various downstream tasks for AD patients, such as cognitive level, Braak stage, and APOE, AD-MIF was compared with general machine learning and deep learning prediction models based on omics data. To ensure the reliability of the results, the baseline models employed the same data splitting method as AD-MIF and used the average values from five-fold cross-validation as the final outcomes. Regarding performance evaluation metrics, the primary focus was on the Area Under the Curve (AUC), as it provides a comprehensive assessment of the model’s classification performance under varying conditions.

As shown in Table 1, AD-MIF achieved the best average performance in predicting various downstream tasks related to AD. Compared to traditional machine learning algorithms such as Support Vector Machine (SVM), Logistic Regression (LR), and Random Forest (RF), AD-MIF exhibited an average AUC increase of 10%-20% across different downstream tasks. This indicates that AD-MIF effectively captures and utilizes more complex feature information, demonstrating a stronger ability to differentiate gene expression and genotype features associated with AD. Compared to deep learning algorithms like ResNet and Recurrent Neural Networks (RNN), AD-MIF achieved an average AUC improvement of over 15%, suggesting that AD-MIF’s specific fusion modules, which integrate potential feature information from multiple omics datasets, further optimize feature representation and model learning. Notably, when compared to modality imputation deep learning methods such as DeepGAMI and GenNet, AD-MIF also improved accuracy by approximately 1%-5%. This suggests that AD-MIF enhances the model’s generalization ability and accuracy by establishing more effective associations between the original omics features and neighborhood features.

**Table 1.**
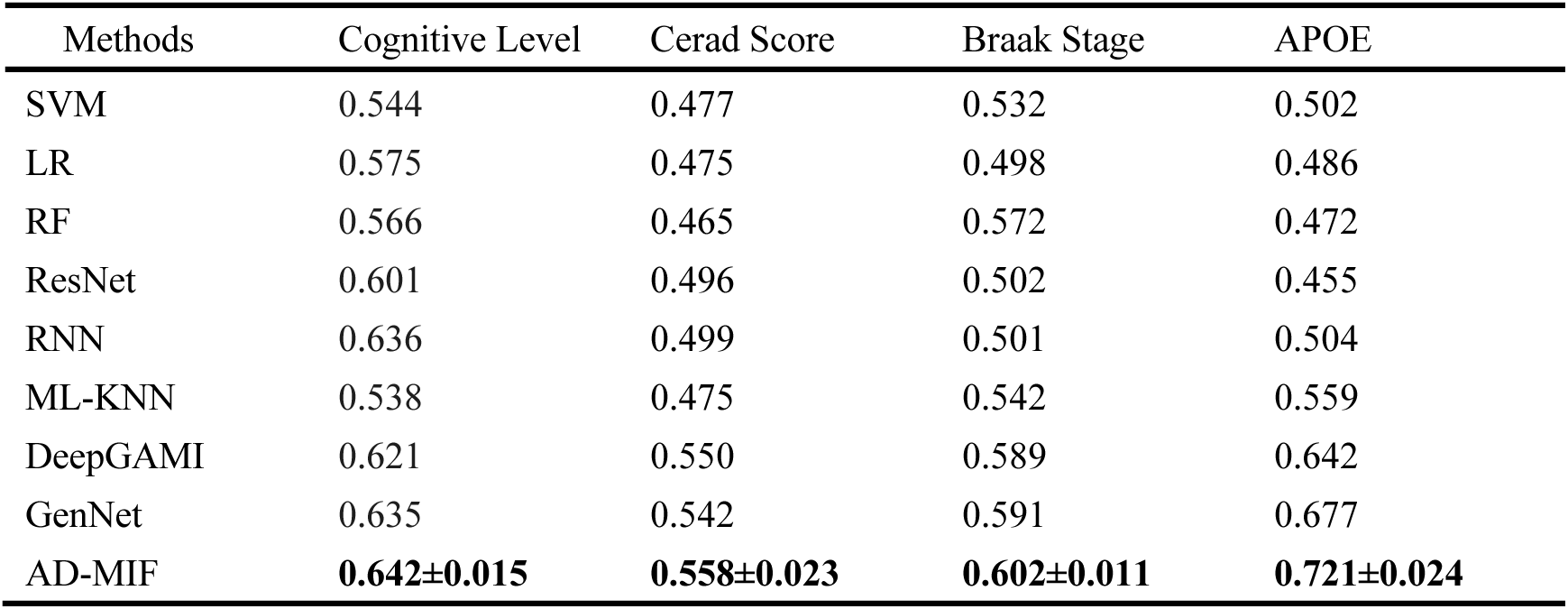
Performance of AD-MIF.

| Methods | Cognitive Level | Cerad Score | Braak Stage | APOE |
| --- | --- | --- | --- | --- |
| SVM | 0.544 | 0.477 | 0.532 | 0.502 |
| LR | 0.575 | 0.475 | 0.498 | 0.486 |
| RF | 0.566 | 0.465 | 0.572 | 0.472 |
| ResNet | 0.601 | 0.496 | 0.502 | 0.455 |
| RNN | 0.636 | 0.499 | 0.501 | 0.504 |
| ML-KNN | 0.538 | 0.475 | 0.542 | 0.559 |
| DeepGAMI | 0.621 | 0.550 | 0.589 | 0.642 |
| GenNet | 0.635 | 0.542 | 0.591 | 0.677 |
| AD-MIF | **0.642±0.015** | **0.558±0.023** | **0.602±0.011** | **0.721±0.024** |

In summary, AD-MIF enhances the expression capability and information utilization efficiency of different modal features by integrating and learning various omics characteristics. This provides robust theoretical and technical support for the practical applications of AD-MIF in the early detection, progression prediction, and personalized treatment of AD.

Additionally, to further examine the effectiveness of different configurations of AD-MIF, this study established the following AD-MIF variants:

- AD-MIF without genotype data (w/o GT): Genotype data input was removed.
- AD-MIF without modal fusion (w/o MF): The modal fusion algorithm was removed.

All AD-MIF variants were processed using the same data splitting method, and their effectiveness was evaluated using the AUC metric. The experimental results are presented in Table 2. The AD-MIF variant without genotype data (w/o GT) led to a decrease in model accuracy of approximately 5.5% across all AD-related prediction tasks. This indicates that AD-related gene phenotype data provide valuable biological guidance to the model, enabling it to more comprehensively capture the complex biological features of AD and thus improve prediction accuracy. The results from removing the genetic/SNP neighborhood feature graph representation (w/o Gen-G/SNP-G) demonstrate that the various interaction relationships between neighborhoods enhance AD-MIF’s ability to represent and utilize features extracted from omics data efficiently.

**Table 2.** AD-MIF Ablation Experiment Results.

| Methods | Cognitive Level | Cerad Score | Braak Stage | Apoe |
| --- | --- | --- | --- | --- |
| AD-MIF(w/o GT) | 0.582 | 0.552 | 0.534 | 0.634 |
| AD-MIF(w/o Gen-G /SNP-G) | 0.560 | 0.557 | 0.531 | 0.584 |
| AD-MIF(w/o MF) | 0.561 | 0.544 | 0.530 | 0.592 |
| AD-MIF | **0.642±0.015** | **0.558±0.023** | **0.602±0.011** | **0.721±0.024** |

Notably, the AD-MIF variant without modal fusion (w/o MF) caused the largest decrease in model prediction accuracy, with an average reduction of 7.3%. This suggests that the information fusion module employed by AD-MIF, by integrating local and global features, generates more consistent feature representations. This, in turn, provides higher-quality features for AD-related tasks, enhancing both the prediction accuracy and stability of the model.

### Visualization and Analysis of AD-MIF Model Results Based on Gene Expression Data

To investigate the specific roles of related genes in AD-related phenotype prediction tasks, the experiment extracted neighborhood feature graph representations of gene expression under different AD tasks (Gen-G) and analyzed the importance scores of each gene across various tasks. Specifically, the AD-MIF model calculated the attention weights for each gene based on Gen-G. By analyzing these weight scores, deeper insights can be gained into the relative contributions of genes across different tasks, revealing their biological significance and potential functional roles in AD prediction. As shown in Figures 4a-d, the results indicate that different genes make significant contributions to various phenotypes of AD, including cognitive level, CERAD score, Braak stage, and APOE genotype. Key genes such as ADRA1A, CAMKK1, SLC30A4, and PNKD are likely closely related to neural signal transmission, amyloid plaque formation, neurofibrillary tangle progression, and APOE-related risk regulation. These genes provide essential insights into understanding the phenotypic differences of AD, its underlying mechanisms, and the exploration of diagnostic and therapeutic targets.

**Figure 4.**
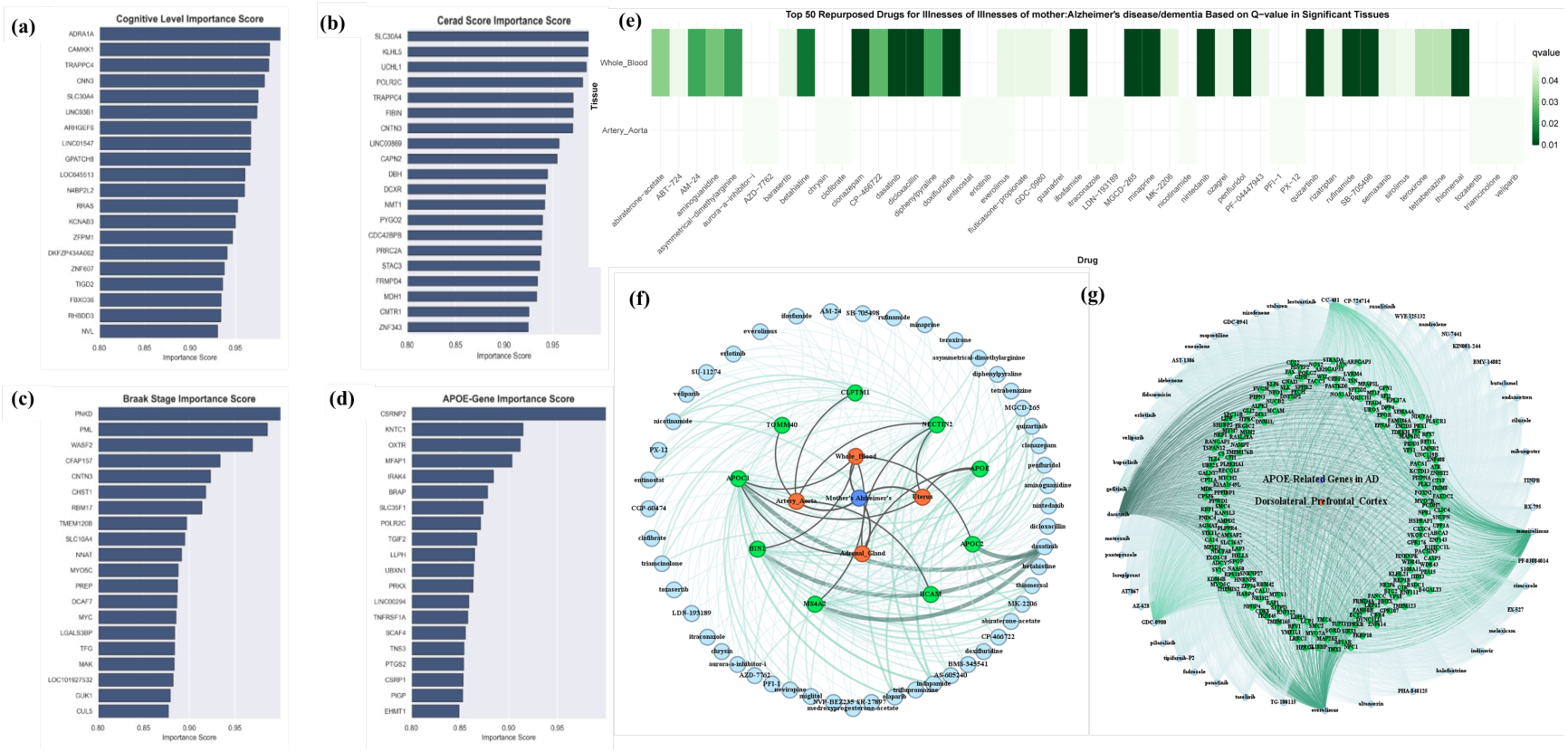
AD Phenotype Prediction and Drug Repurposing: A Comprehensive Analysis of Multi-layered Gene, Tissue, and Drug Interaction Networks. **a) Importance scores of genes in the Cognitive Level prediction task; b) Importance scores of genes in the CERAD Score prediction task; c) Importance scores of genes in the Braak Stage prediction task; d) Importance scores of genes in the APOE prediction task; e) Top 50 Repurposed Drugs for Mother’s AD/Dementia Based on Q-value in Significant Tissues.** This figure illustrates the top 50 potential repurposed drugs identified in significant tissues (Whole Blood and Artery Aorta) for maternal AD/Dementia history, visualizing the drug effects based on q-values. Due to the small sample size and the presence of numerous duplicate values in the data, some correlation coefficients could not be calculated; these data were saved in non-significant tissue files. **f) Mother’s AD: Trait-Tissue-Gene-Drug Interaction Network.** This network visualizes the interactions between Mother’s AD, various tissues, key genes, and potential repurposed drugs. The tissues involved are Artery Aorta, Adrenal Gland, Uterus, and Whole Blood, which play important roles in gene-tissue interactions related to AD. Key genes include CLPTM1, NECTIN2, APOE, TOMM40, APOC1, BIN1, MS4A2, BCAM, and APOC2, all linked to Alzheimer’s pathology. Drug-Gene interactions are weighted by −*log*_10_(*p*) to reflect the strength of the associations, while Tissue-Gene interactions have a fixed weight of 8. This network offers insights into drug repurposing strategies by focusing on crucial tissue and gene interactions in AD. **g) APOE-Related Trait-Tissue-Gene-Drug Network for AD Drug Repurposing.** This network illustrates interactions between APOE-related traits, tissues, genes, and repurposed drugs for AD. Nodes represent genes linked to APOE-related phenotypes (from PrediXcan analysis), the dorsolateral prefrontal cortex, and potential repurposed drugs. Edges indicate drug-gene associations weighted by −*log*_10_(*p*) and tissue-gene connections with a fixed weight of 8. Drugs were selected based on significant associations with APOE-related genes (p ≤ 0.05). The network reveals how genetic and tissue-specific interactions inform drug repurposing strategies for AD.

Additionally, to explore the feasibility and effectiveness of artificial intelligence methods in studying disease mechanisms, enrichment analyses were conducted on key genes closely related to cognitive diagnostic scores associated with AD risk (Supplementary Table S9). The genes predicted by the model to be associated with cognition were significantly enriched in GO components, such as intracellular anatomical structures (e.g., organelle maintenance and signal transduction), nucleoplasm, and nuclear lumen. This suggests that these genes play crucial roles in key biological processes, such as gene expression regulation, signal transduction, and cellular function maintenance, which support normal neuronal functions and cognitive processes (Supplementary Figure S2c). Furthermore, tissue enrichment analysis revealed that these genes are highly expressed in the brain and nervous system, further confirming their critical roles in neurosignal transmission, synaptic plasticity, and neuronal development—biological processes related to cognition. These findings are consistent with previous studies, emphasizing the importance of intracellular, particularly nuclear, functions and gene expression in the central nervous system for cognitive regulation. This provides a solid foundation for the in-depth exploration of the molecular mechanisms underlying cognitive processes.

### Drug Repositioning Analysis and Potential Therapeutic Drugs for AD

#### Drug Repositioning Analysis for AD Family History-Related Traits

This study utilized the S-PrediXcan method to analyze gene expression levels associated with three AD family history traits: a history of AD/dementia in the father, mother, and siblings. Significant genes were identified with a p-value ≤ 0.01, followed by a drug repositioning analysis. By calculating q-values and performing manual filtering, the top 50 most promising drugs from each trait and tissue were extracted. The results are presented in heatmaps and bar charts. Additionally, the top 20 most significant candidate drugs for each AD family history trait are listed in Table 3, Supplementary Table S10, and Supplementary Table S11. Intersection analysis further revealed common drugs and tissues across the three AD family history traits, suggesting that these drugs may have significant potential in treating multiple AD-related symptoms (Supplementary Figure S2d).

**Table 3.** Top Repositioning Hits for Mother’s AD/Dementia (Ranked by Q-values and P-values)

| Drug | Tissue | Rank | P-value | Q-value | Z-score | Brief description |
| --- | --- | --- | --- | --- | --- | --- |
| MGCD-265 | Whole Blood | 1 | 9.31e-08 | 9.92e-03 | -1.756 | Identified only through virtual screening, no psychiatric relevance. |
| Nintedanib | Whole Blood | 2 | 1.86e-07 | 9.92e-03 | -1.756 | Identified as AD repurposing candidate due to proximity to oligodendrocytes disease modules[24]. |
| Quizartinib | Whole Blood | 3 | 2.79e-07 | 9.92e-03 | -1.756 | No psychiatric relevance. Protects against TNF-induced inflammation in mice via *Ripk1* inhibition[25]. |
| SB-705498 | Whole Blood | 4 | 3.72e-07 | 9.92e-03 | -1.756 | No psychiatric relevance, only related to TRP channels. |
| Dasatinib | Whole Blood | 5 | 4.66e-07 | 9.92e-03 | -1.756 | Targets tyrosine kinase activity, key for AD. Clarifies microglial activation. Promising in age-related disease therapies[26]. |
| Clonazepam | Whole Blood | 6 | 5.59e-07 | 9.92e-03 | -1.756 | Reduces NO release and microglial activity in LPS-induced conditions[27]. |
| Rufinamide | Whole Blood | 7 | 6.52e-07 | 9.92e-03 | -1.756 | Suppresses neuronal hyperexcitability, approved for Lennox-Gastaut Syndrome[28]. |
| Doxifluridine | Whole Blood | 8 | 7.45e-07 | 9.92e-03 | -1.756 | No known relevance, identified through virtual screening. |
| Thiomersal | Whole Blood | 9 | 8.38e-07 | 9.92e-03 | -1.756 | No psychiatric relevance. Controversial mercury-based preservative, possible toxicity. |
| Dicloxacillin | Whole Blood | 10 | 9.31e-07 | 9.92e-03 | -1.756 | No psychiatric relevance. Effective against MSSA bacteraemia with a good safety profile[29]. |
| Minaprine | Whole Blood | 11 | 1.02e-06 | 9.92e-03 | -1.756 | No psychiatric relevance, identified through virtual screening. |
| Penfluridol | Whole Blood | 12 | 1.12e-06 | 9.92e-03 | -1.756 | An antipsychotic drug with potential anti-cancer properties[30]. |
| MK-2206 | Whole Blood | 13 | 1.21e-06 | 9.92e-03 | -1.756 | No psychiatric relevance. Inhibits cell viability in DLBCL lines via Akt inhibition[31]. |
| Ifosfamide | Whole Blood | 14 | 1.30e-06 | 9.92e-03 | -1.756 | Identified through virtual screening, no further relevance. |
| Betahistine | Whole Blood | 15 | 2.33e-06 | 1.65e-02 | -1.756 | May reduce vertigo symptoms, generally well-tolerated[32]. |
| Asymmetrical-Dimethylarginine | Whole Blood | 16 | 3.35e-06 | 2.23e-02 | -1.756 | No psychiatric relevance. Endogenous NO synthesis inhibitor. |
| AM-24 | Whole Blood | 17 | 3.91e-06 | 2.45e-02 | -1.756 | No psychiatric relevance. |
| Diphenylpyraline | Whole Blood | 18 | 4.19e-06 | 2.48e-02 | -1.756 | No psychiatric relevance. Inhibits Ebola virus proliferation[33]. |
| CP-466722 | Whole Blood | 19 | 4.93e-06 | 2.77e-02 | -1.756 | No psychiatric relevance. Identified as an ATM inhibitor, no psychiatric relevance. |
| Aminoguanidine | Whole Blood | 20 | 5.68e-06 | 3.02e-02 | -1.756 | No psychiatric relevance. Potent AGE inhibitor, alleviates diabetic complications[34]. |

‘Rank’ refers to the ranking within each tissue for the disorder under study.

#### Drug Repositioning Analysis for Maternal AD/Dementia History

In the drug repositioning analysis for maternal AD/dementia family history, drugs such as Dasatinib and Sirolimus exhibited significant potential in tissues like whole blood and the aorta (see Figure 4e). Dasatinib shows promise in AD treatment by clearing senescent cells, thereby alleviating neurodegeneration and associated mood symptoms. Sirolimus, on the other hand, slows neuroinflammation in AD by modulating immune responses, further validating its broad potential in various neurodegenerative diseases. Additionally, drugs like Guanidine, Mianpril, and others demonstrated potential in treating AD, particularly showing positive effects in regulating the vascular and circulatory systems.

#### Drug Repositioning Analysis for Paternal AD/Dementia History

In the drug repositioning analysis for paternal AD/dementia history, drugs identified in adrenal gland tissue, such as Transitro, Isoxsuprine, and Tamoxifen, also showed potential in treating AD or cognitive decline (Supplementary Figure S3c). The mechanisms of these drugs may involve improving metabolic functions and regulating blood pressure, which could positively impact AD-related vascular lesions and the cognitive impairments they cause.

#### Drug Repositioning Analysis for Sibling AD/Dementia History

In the drug repositioning analysis for sibling AD/dementia history (Supplementary Figure S3d), drugs identified in gastric tissue, including Catechin, Imipramine, and Pioglitazone, demonstrated significant repositioning potential in the digestive system and metabolic regulation. Imipramine, a commonly used antidepressant, plays a positive role in AD’s pathological processes by reducing inflammatory responses and improving neuroprotection. The identification of these drugs provides potential therapeutic targets, particularly in alleviating AD symptoms by regulating metabolism and reducing neuroinflammation.

Furthermore, based on drug repositioning analyses for emotion-related features such as irritability and anxiety/worry, several drugs demonstrated significant repositioning potential across multiple tissues, including Pantoprazole, Mitoxantrone, and Balaglitazone (see Supplementary Figures S3a and S3b). Further analysis results are presented in Supplementary Tables S12 and S13, which list the drug repositioning outcomes for irritability and anxiety/worry based on Q-values and P-values rankings, respectively. These drugs exhibit significant potential in regulating neurological functions and cardiovascular metabolism, offering new possibilities for managing patients’ emotional symptoms.

The drug repositioning analysis also revealed the reutilization potential of various existing drugs. For example, Nintedanib, primarily used to treat fibrotic diseases, may exhibit neuroprotective effects in AD by reducing myelin oligodendrocyte cell degeneration. Quizartinib, by inhibiting RIPK1 signaling, reduces inflammatory responses, suggesting its potential application in neuroinflammation. Additionally, MK-2206, an Akt inhibitor, may protect against neurodegenerative changes in AD by regulating cell survival and apoptosis. Drugs such as Sirolimus and Dasatinib showed significant efficacy across multiple AD-related phenotypes, particularly excelling in immune regulation and clearing senescent cells. The multiple mechanisms of action of these drugs provide strong support for their application in AD treatment.

#### Drug Repurposing Analysis through Trait-Tissue-Gene-Drug Networks

To further validate the reliability of the drug repositioning results, this study conducted an in-depth analysis of the potential drugs identified for AD family history-related traits by constructing trait-tissue-gene-drug interaction networks. These networks integrated gene expression data from the LINCS L1000 database, providing systematic insights into AD drug repositioning. By analyzing drugs with a significance level of q ≤ 0.001 associated with maternal, paternal, and sibling AD-related traits, we revealed the action pathways of these drugs across various AD-related tissues. Interestingly, the reverse targeting results differed from the forward drug repositioning results, indicating drug-tissue specificity and distinct gene-associated pathways.

In the interaction network for maternal AD traits, tissues such as the aorta, adrenal glands, uterus, and whole blood exhibited significant gene-tissue associations (Figure 4f). Key genes, including CLPTM1, NECTIN2, APOE, and TOMM40, are closely related to the pathological processes of AD. These genes may play crucial roles in the onset and progression of AD by regulating biological processes such as cholesterol metabolism, immune responses, and cell adhesion. For example, TOMM40 showed strong significance in the aorta, suggesting its potential core role in regulating vascular function and metabolism.

In the network for paternal AD traits, adrenal gland tissue showed close interactions with key genes APOE, APOC1, and NECTIN2 (Supplementary Figure S4a). APOE and APOC1 play critical roles in regulating lipid metabolism and clearing amyloid plaques, both of which are central to the pathogenesis of AD. The associations of these genes with multiple drugs were also significant, suggesting that these drugs may have therapeutic potential in reducing neuroinflammation and regulating metabolism. Through network analysis, we were able to visualize how these drugs interact with key genes in specific tissues, further supporting the potential application of these drugs in AD treatment.

The interaction network for sibling AD traits indicated that the stomach and adrenal glands are important target tissues (Supplementary Figure S4b). Key genes associated with these tissues include TOMM40, APOE, APOC1, and ABCB6. TOMM40 and APOE exhibited strong significance in their interactions with drugs across multiple tissues, particularly in the stomach. These genes may play dual roles in the pathological processes of AD by regulating metabolism and immune functions. Additionally, the enrichment of the mitochondrial autophagy pathway for the ABCB6 gene suggests its potential role in cellular homeostasis and metabolic regulation. These findings provide new targets for AD treatment, particularly by improving metabolism and enhancing cellular homeostasis to alleviate the disease.

Interaction networks for emotion-related traits such as irritability and anxiety/worry also demonstrated potential for drug repositioning. Supplementary Figures S4c and S4d detail the trait-tissue-gene-drug interaction networks for these traits. The results indicate that some drugs show consistent significance across multiple tissues, particularly in the nervous and cardiovascular systems. The multi-tissue and multi-gene associations of these drugs suggest that they may influence emotional symptoms through multiple pathways, offering new therapeutic approaches for managing emotions in AD patients.

These trait-tissue-gene-drug networks reveal the complexity of the pathological mechanisms of AD, emphasizing the importance of adopting tissue-specific approaches in drug repositioning. The results of the network analysis provide insights into how to utilize gene-tissue-specific interactions for drug screening, thereby facilitating the development of personalized treatment plans. These findings also suggest that future research should place greater emphasis on tissue and gene specificity to more accurately select targeted drugs, thereby enhancing the efficacy and safety of AD treatments.

#### Drug Repositioning Analysis for APOE Genotype

This study focused on significant genes associated with the APOE genotype and conducted drug repositioning analyses on these genes (Supplementary Figure S3e). By performing a detailed analysis of the genes significantly associated with the APOE genotype, we identified a series of drugs with potential therapeutic effects for AD (Table 4). These drugs may effectively intervene in APOE genotype-related AD pathological processes through various pathways, including the regulation of metabolism, immune responses, and neuroprotection.

**Table 4.** Top Repositioning Hits for APOE Genotype-Related Genes in AD/Dementia (Ranked by P-values)

| Drug | Tissue | Rank | P-value | Brief description |
| --- | --- | --- | --- | --- |
| Gefitinib | Dorsolateral Prefrontal Cortex | 1 | 2.38E-05 | EGFR inhibitor, reduces AD pathology and improves cognitive function[35]. |
| Idebenone | Dorsolateral Prefrontal Cortex | 1 | 2.38E-05 | Ameliorates memory impairment by promoting Aβ42 clearance and restoring antioxidant status[36]. |
| Rimcazole | Dorsolateral Prefrontal Cortex | 2 | 3.33E-05 | No psychiatric relevance. |
| Tipifarnib-P2 | Dorsolateral Prefrontal Cortex | 3 | 4.93E-05 | No psychiatric relevance. |
| BX-795 | Dorsolateral Prefrontal Cortex | 4 | 8.25E-05 | No psychiatric relevance. |
| Pantoprazole | Dorsolateral Prefrontal Cortex | 5 | 8.98E-05 | Linked to a reduced risk of AD in follow-up studies[37]. |
| Butaclamol | Dorsolateral Prefrontal Cortex | 6 | 1.03E-04 | No psychiatric relevance. |
| Dasatinib | Dorsolateral Prefrontal Cortex | 7 | 1.04E-04 | Preclinical evidence for senescent cell clearance, reducing AD pathology[38]. |
| Taselisib | Dorsolateral Prefrontal Cortex | 7 | 1.04E-04 | No psychiatric relevance. |
| Motesanib | Dorsolateral Prefrontal Cortex | 8 | 1.07E-04 | No psychiatric relevance. |
| Everolimus | Dorsolateral Prefrontal Cortex | 9 | 1.16E-04 | No psychiatric relevance. |
| Meloxicam | Dorsolateral Prefrontal Cortex | 10 | 1.34E-04 | No psychiatric relevance. |
| Dasatinib | Dorsolateral Prefrontal Cortex | / | / | / |
| Temsirolimus | Dorsolateral Prefrontal Cortex | 11 | 1.59E-04 | Reduces neurofibrillary tangle density and improves memory[39]. |
| TTNPB | Dorsolateral Prefrontal Cortex | 12 | 1.62E-04 | Promotes glia-neuron conversion to replace impaired neurons[40]. |
| TG-100115 | Dorsolateral Prefrontal Cortex | 13 | 1.73E-04 | No relevant findings. |
| EX-527 | Dorsolateral Prefrontal Cortex | 14 | 1.95E-04 | No relevant findings. |
| CC-401 | Dorsolateral Prefrontal Cortex | 15 | 1.98E-04 | Trials terminated, possible toxicity. |
| AST-1306 | Dorsolateral Prefrontal Cortex | 16 | 2.01E-04 | No relevant findings. |
| GDC-0980 | Dorsolateral Prefrontal Cortex | 17 | 2.26E-04 | No relevant findings. |

**Table 5.** Drug enrichment analyses P values with drug-sets defined in the ATC classification system.

| Disorder | Self (Fisher) | Compet (Fisher) | Self (minP) | Compet (minP) |
| --- | --- | --- | --- | --- |
| **Enrichment for ATC antiepileptics (N03)** |  |  |  |  |
| Irritability | 0 | 0.653 | 1.78E-91 | 0.397 |
| Worrier / anxious feelings | 0 | 1.56E-05 | 1.91E-92 | 5.80E-3 |
| Illnesses of father: AD/dementia | 0 | 0.221 | 1.08E-121 | 0.465 |
| Illnesses of mother: AD/dementia | 0 | 0.411 | 5.63E-117 | 0.661 |
| Illnesses of siblings: AD/dementia | 0 | 0.789 | 4.30E-114 | 0.816 |
| **Enrichment for ATC anti-Parkinson (N04)** |  |  |  |  |
| Irritability | 0 | 0.156 | 4.34E-12 | 0.770123035 |
| Worrier / anxious feelings | **0** | **6.37E-2** | **3.80E-14** | **2.07E-2** |
| Illnesses of father: AD/dementia | **6.51E-101** | **0.459** | **1.07E-13** | **0.106** |
| Illnesses of mother: AD/dementia | 3.21E-299 | 0.860 | 9.35E-15 | 0.851 |
| Illnesses of siblings: AD/dementia | 5.47E-39 | 0.417 | 4.88E-14 | 0.553 |
| **Enrichment for ATC antipsychotics (N05A)** |  |  |  |  |
| Irritability | 0 | 0.223 | 1.58E-20 | 0.604 |
| Worrier / anxious feelings | 0 | 8.83E-2 | 8.63E-20 | 0.832 |
| Illnesses of father: AD/dementia | 7.59E-188 | 0.780 | 1.89E-24 | 0.614 |
| Illnesses of mother: AD/dementia | 0 | 0.950 | 3.33E-24 | 0.996 |
| Illnesses of siblings: AD/dementia | 1.44E-67 | 0.617 | 2.83E-23 | 0.631 |
| **Enrichment for ATC anxiolytics or antidepressants (N05B or N06A)** |  |  |  |  |
| Irritability | 0 | 1.01E-06 | 6.30E-112 | 0.257 |
| Worrier / anxious feelings | **0** | **8.11E-3** | **3.06E-101** | **9.76E-2** |
| Illnesses of father: AD/dementia | 0 | 0.155 | 9.54E-141 | 0.471 |
| Illnesses of mother: AD/dementia | 0 | 0.711 | 1.24E-145 | 0.356 |
| Illnesses of siblings: AD/dementia | 0 | 0.967 | 1.49E-136 | 0.961 |
| **Enrichment for all ATC psychiatric drugs (N05 or N06)** |  |  |  |  |
| Irritability | 0 | 0.885 | 8.60E-146 | 1.00 |
| Worrier / anxious feelings | 0 | 0.949 | 5.97E-134 | 1.00 |
| Illnesses of father: AD/dementia | 0 | 0.792 | 1.72E-179 | 0.980 |
| Illnesses of mother: AD/dementia | 0 | 0.928 | 2.28E-186 | 1.00 |
| Illnesses of siblings: AD/dementia | 0 | 0.701 | 4.64E-176 | 0.829 |

In the dorsolateral prefrontal cortex, several important drugs were identified through the gene-drug interaction network (Figure 4g). These drugs showed significant associations with key genes such as APOE and TOMM40. Notably, Sirolimus and Dasatinib exhibited strong significance across multiple genes, potentially playing critical roles in alleviating neuroinflammation and clearing senescent cells. Additionally, Everolimus and Temsirolimus demonstrated potential in AD treatment by regulating the mTOR signaling pathway, which helps mitigate neurodegenerative lesions.

During the drug screening for the APOE genotype, several drugs with specific neuroprotective effects were also identified. For example, Gefitinib, an EGFR inhibitor, showed potential in reducing AD pathology and improving cognitive function. Pantoprazole, a proton pump inhibitor, demonstrated a role in decreasing AD risk in follow-up studies. Moreover, preclinical evidence supporting Dasatinib’s ability to clear senescent cells further endorses its potential application as a therapeutic agent for AD.

The significance analysis of these drugs was weighted based on -log₁₀(p) to reflect the strength of the association between the drugs and the genes. By visualizing the multiple action pathways of these drugs in specific tissues, a better understanding of their therapeutic mechanisms can be gained. These findings highlight the potential of drug repositioning in a personalized context, especially for patients with the APOE genotype, where these drugs may offer more effective intervention strategies.

In summary, through drug repositioning analysis related to the APOE genotype, we identified a series of drugs with potential therapeutic effects. These drugs may provide effective treatment options for AD patients by regulating metabolism, reducing neuroinflammation, and clearing senescent cells through various mechanisms. Future research should further explore the in vivo and in vitro experimental validation of these drugs and evaluate their safety and efficacy in patients with specific APOE genotypes through clinical trials, thereby advancing the development of personalized treatments.

#### Drug Enrichment Analysis Results

In this study, we conducted a comprehensive analysis of drug repositioning enrichment to assess the potential roles of various drug categories in the treatment of AD. Using drug sets based on the ATC classification system and MEDI-HPS definitions, we performed a series of enrichment tests to identify the significance of specific drug categories in relation to AD-related traits.

### Drug enrichment analysis based on ATC classification system

The results of the drug enrichment analysis, based on the ATC classification system, revealed that antiepileptic drugs (N03), antiparkinsonian drugs (N04), antipsychotic drugs (N05A), and anxiolytic/antidepressant drugs (N05B or N06A) were significantly enriched across various AD-related traits. Notably, the significance of these drugs was particularly evident in emotion-related traits, such as irritability and anxiety/worry. These findings suggest that psychotropic medications may play a significant role in alleviating AD-related emotional symptoms by modulating neurotransmitters or reducing neuroinflammation.

#### Drug enrichment analysis based on the definition of MEDI-HPS

In the enrichment analysis of drug sets defined by MEDI-HPS, we further explore d the roles of drugs associated with specific mental illnesses, such as depression and anxiety disorders, in the treatment of AD (Supplementary Table S14). The results revealed that drugs linked to depression and anxiety disorders were significantly enriched across multiple AD-related traits. Notably, in traits associated with a parental history of AD or dementia, antidepressant drugs showed a higher level of significance. This suggests that these medications may positively impact the emotional symptoms of AD patients by regulating mood and improving cognitive function.

#### Drug Enrichment Analysis Based on ClinicalTrials

The drug enrichment analysis based on clinical trial data revealed that drugs associated with irritability and AD were significantly enriched across multiple tissues, while drugs related to anxiety/worry exhibited relatively lower enrichment (Supplementary Table S15). This finding suggests that certain medications may be more effective for targeting specific AD symptoms, whereas others may demonstrate better efficacy for different symptoms.

Overall, the results of the drug enrichment analysis offer valuable insights into selecting the most appropriate medications for addressing various AD symptoms. In particular, the significant enrichment of psychotropic drugs in emotion management suggests that they may play a crucial role in the personalized treatment of AD. Future research should further validate the mechanisms of these drugs and evaluate their efficacy and safety in AD patients through clinical trials, providing more comprehensive and effective treatment options for patients.

### MGCD-265 and Dasatinib Ameliorate Cognitive, Inflammatory, and Alzheimer-like Pathologies in SAMP8 Mice

#### MGCD-265 and Dasatinib Improved Cognitive Function in SAMP8 Mice

The experimental timeline and mouse treatment protocol are illustrated in Figure 5A. Following open-field training conducted one day prior, no significant differences in spatial exploration ability were observed between the groups (Supplementary Figure S5 A-B). In the novel object recognition test, SAMP8 model mice exhibited significantly lower entry frequency, exploration time, and exploration distance toward the novel object compared to the SAMR1 group (Figure 5B). However, oral administration of MGCD-265 or Dasatinib over the treatment period improved novel object exploration in SAMP8 mice to varying degrees (Figure 5C-F), suggesting that both compounds enhance cognitive function in this model. During the Morris water maze test, the SAMR1, MGCD-265, and Dasatinib-treated groups located the hidden platform significantly faster than the SAMP8 model group (Figure 5H). On day 6, the platform was removed for a 1-minute probe trial. While total swimming distance did not differ significantly between groups (Figure 5I), mice treated with MGCD-265 or Dasatinib showed increased platform crossings, indicating improved spatial memory retention. Collectively, these findings demonstrate that both MGCD-265 and Dasatinib ameliorate cognitive deficits in SAMP8 mice.

**Figure 5.**
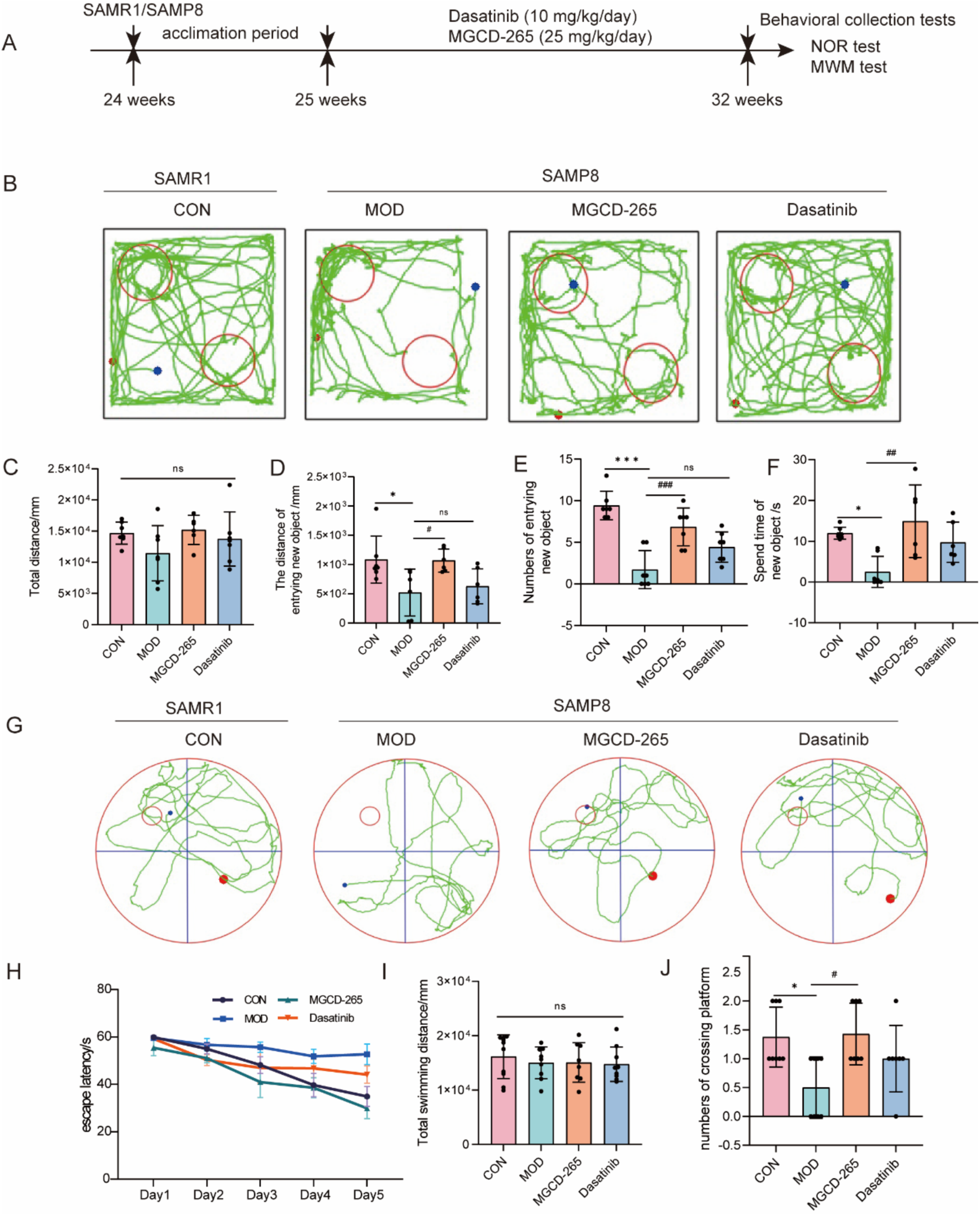
MGCD-265 and Dasatinib improved cognitive function in SAMP8 mice. **(A) Experimental design. (B) Representative exploration trajectory during the test period. (C) Total distance during the test. (D) The distance of entrying new object during the test. (E) Numbers of entrying new object. (F) Spend time of new object. (G) Representative swimming trajectories during the test period. (H) The escape latency during the training period. (I) Total swimming distance during the test period. (J) Numbers of crossing platform within 1min during the test. (n=7-9).** Data are presented as mean ± SD. Statistical analysis was performed using one-way analysis of variance (ANOVA) and followed by Tukey’s multiple comparisons test. SAMP8-MOD VS SAMR1-CON, * P < 0.05, **P < 0.01, ***P < 0.001, MGCD-265 and Dasatinib VS SAMP8-MOD, ^#^ P < 0.05, ^##^P < 0.01, ^###^P < 0.001, ns: no statistical difference.

#### MGCD-265 and Dasatinib Attenuate Glial Hyperactivation and Neuroinflammation in SAMP8 Mice

In SAMP8 mice, the activation states of microglia and astrocytes were assessed via immunofluorescence, and the release of inflammatory cytokines and chemokines was quantified using multiplex cytokine arrays. The activation levels of microglia and astrocytes were statistically analyzed based on the immunofluorescence positivity rates of IBA1 and GFAP, respectively. Results demonstrated a significant increase in GFAP-labeled astrocytes in the dentate gyrus (DG), CA1, and CA3 regions of SAMP8 mice compared to SAMR1 control mice (Figure 6B–C and Supplementary Figure S6A–D), indicating astrocytic hyperactivation. However, treatment with MGCD-265 and Dasatinib markedly ameliorated this pathological activation (Figure 6B–C). Furthermore, microglial hyperactivation in the DG region of SAMP8 mice was also significantly reversed by both compounds (Figure 6D–E). Additionally, we examined the expression of the synaptic marker PSD95. Intriguingly, SAMP8 mice exhibited varying degrees of synaptic loss, with Dasatinib showing a partial restorative effect on synaptic integrity (Figure 6F). Given that microglial and astrocytic hyperactivation typically accompanies the release of pro-inflammatory mediators, we profiled hippocampal cytokine levels. Multiplex analysis revealed elevated levels of IFN-γ, IL-1β, CCL11, and IP-10 in SAMP8 mice, which were partially suppressed by MGCD-265 and Dasatinib (Figure 6G and Supplementary Figure S2E). Similarly, whole-brain tissue lysates confirmed the reversal of aberrant cytokine overproduction in SAMP8 mice (Supplementary Figure S6F–G). In summary, our findings demonstrate that MGCD-265 and Dasatinib effectively mitigate glial hyperactivation and attenuate neuroinflammation in SAMP8 mice.

**Figure 6.**
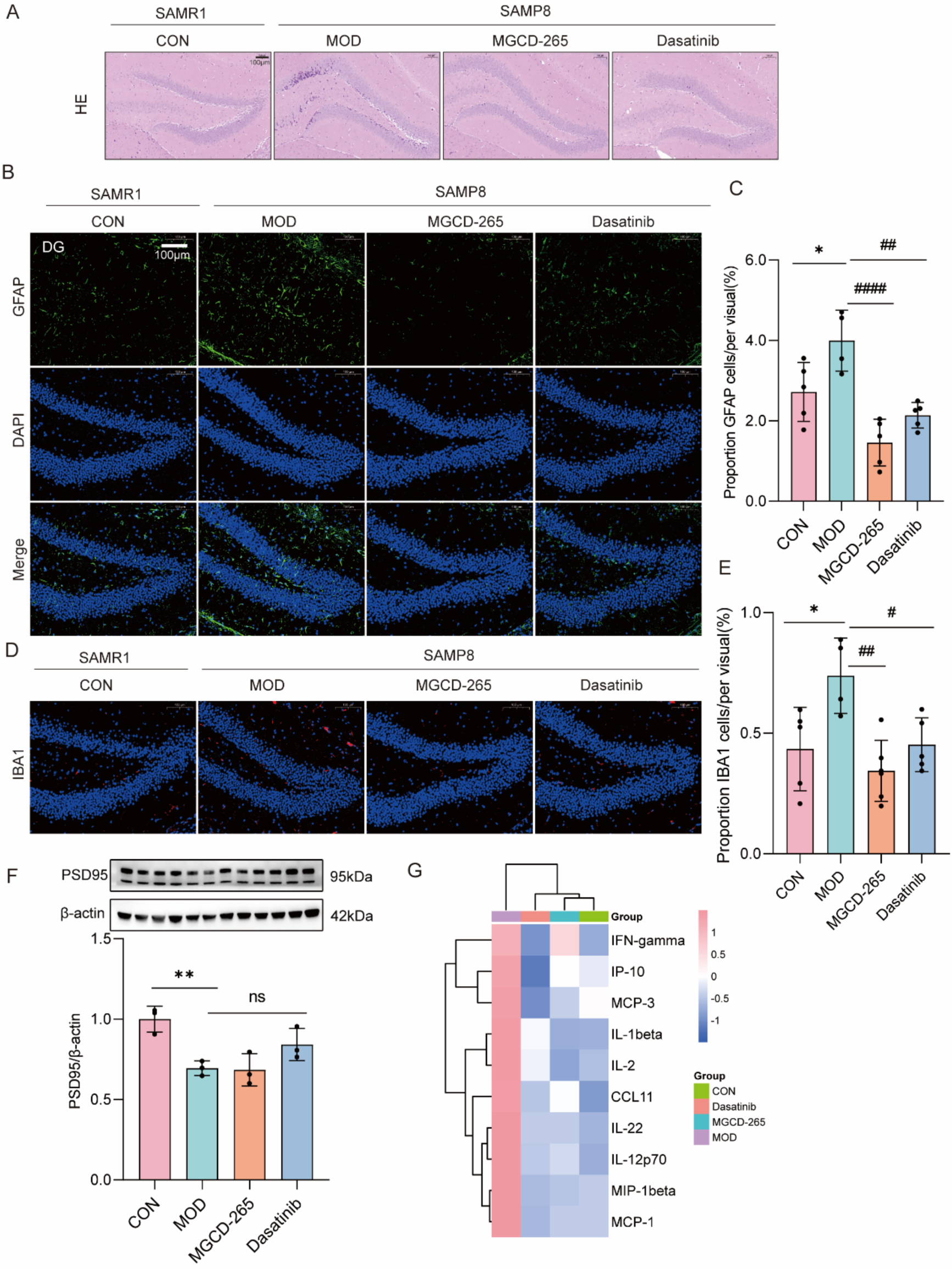
MGCD-265 and Dasatinib attenuate glial hyperactivation and neuroinflammation in SAMP8 mice. **(A) HE staining. (B-C) Immunofluorescence and quantification of GFAP in the hippocampus DG area (Scale bars, 100 μm). (D-E) Immunofluorescence and quantification of IBA1 in the hippocampus DG area (Scale bars, 100 μm), n=4-6. (F) Western bolt and quantification of PSD95 in the hippocampus, n=3. (G) Heatmap of cytokine array analysis in hippocampus.** Data are presented as mean ± SD. Statistical analysis was performed using one-way analysis of variance (ANOVA) and followed by Tukey’s multiple comparisons test. SAMP8-MOD VS SAMR1-CON, * P < 0.05, **P < 0.01, ***P < 0.001, MGCD-265 and Dasatinib VS SAMP8-MOD, ^#^ P < 0.05, ^##^P < 0.01, ^###^P < 0.001, ns: no statistical difference.

#### MGCD-265 and Dasatinib Ameliorate Tau Pathology and Suppress Aβ Deposition

AD is characterized by two prominent pathological features: tau protein pathology and Aβ deposition. We assessed tau phosphorylation levels after drug intervention and quantified Aβ content using thioflavin-S staining. Results showed that SAMP8 mice exhibited reduced Nissl body counts compared to SAMR1 mice, while MGCD-265 and Dasatinib partially restored Nissl body numbers, thereby protecting hippocampal neurons (Figure 7A). Thioflavin-S, a membrane-permeable fluorescent dye that binds amyloid fibrils, revealed significantly higher Aβ deposition in the cortex and hippocampus of SAMP8 mice than in SAMR1 controls. Notably, MGCD-265 and Dasatinib suppressed Aβ accumulation in SAMP8 mice (Figure 7B). Furthermore, SAMP8 mice displayed elevated hippocampal levels of total tau and phosphorylated tau (Ser396, Thr231) compared to SAMR1 mice, and both compounds differentially attenuated tau pathology and hyperphosphorylation. Given that tau phosphorylation is primarily regulated by kinases (e.g., GSK3β, CDK5) and phosphatases (e.g., PP2A), we examined their expression. Intriguingly, GSK3β was upregulated in SAMP8 mice, consistent with increased tau phosphorylation, whereas CDK5 and PP2A showed no significant differences between the groups (Figure 7C-I). Collectively, our study demonstrates that MGCD-265 and Dasatinib ameliorate AD-like phenotypes in SAMP8 mice by mitigating tau pathology and Aβ deposition.

**Figure 7.**
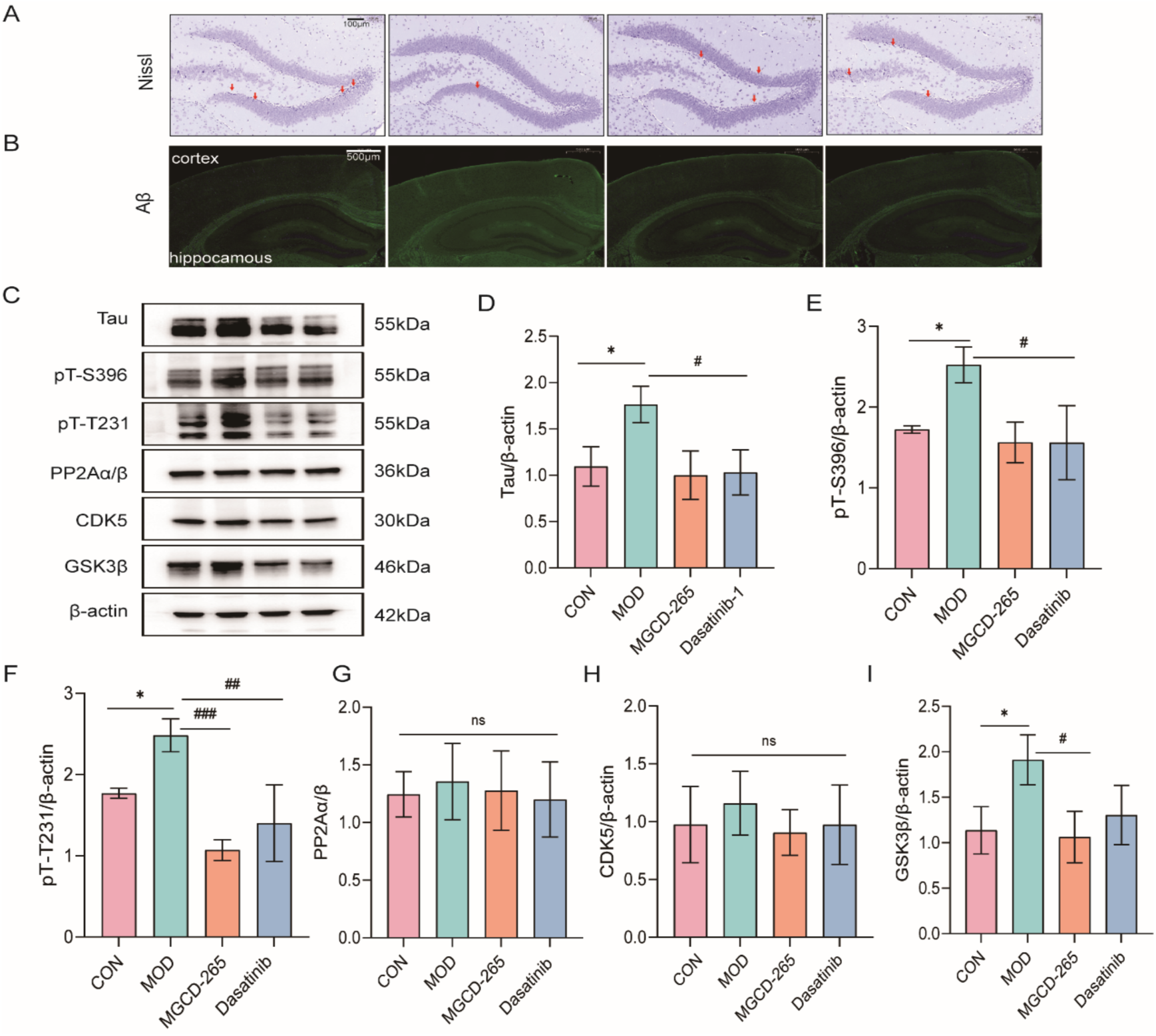
MGCD-265 and Dasatinib ameliorate Tau pathology and suppress Aβ deposition. **(A) Nissl staining. (B) Thioflavin S Immunofluorescence. (C-I) Western bolt and quantification of Tau, pT-S396, pT-T231, PP2Aα/β, CDK5 and GSK3β in the hippocampus n=3.** Data are presented as mean ± SD. Statistical analysis was performed using one-way analysis of variance (ANOVA) and followed by Tukey’s multiple comparisons test. SAMP8-MOD VS SAMR1-CON, * P < 0.05, **P < 0.01, ***P < 0.001, MGCD-265 and Dasatinib VS SAMP8-MOD, ^#^ P < 0.05, ^##^P < 0.01, ^###^P < 0.001, ns: no statistical difference.

## Discussion

### Implications for AD Pathogenesis

In this study, by integrating multi-tissue Transcriptome-Wide Association Studies (TWAS) and multimodal deep learning methods, we identified several key genes associated with AD. These genes play significant roles in the pathogenesis of AD, particularly in the regulation of emotions, cognition, and behavior. The APOE gene, a core risk gene for AD, significantly influences disease progression through its ε4 allele by regulating cholesterol metabolism, lipid transport, and amyloid plaque clearance. APOC1 may promote the pathological progression of AD by cooperating with APOE to influence lipid metabolism and trigger the neuroinflammatory cascade. The SCIMP gene is expressed in immune cells and participates in immune signal transduction, further highlighting the importance of immune responses in AD-related neuroinflammation.

Additionally, the KAT8 gene encodes a histone acetyltransferase involved in the regulation of gene expression. Mutations in KAT8 are associated with neurodevelopmental disorders, suggesting its dual role in emotional and behavioral regulation in AD. Other genes, such as RP11-196G11.6 and VKORC1, are involved in epigenetic regulation and vitamin K metabolism, respectively, suggesting that AD may involve multiple pathological pathways, including vascular pathology and cognitive impairment. PRSS53 and ZNF646 are associated with peripheral tissue lesions, particularly exhibiting regulatory potential in metabolic and inflammatory pathways. Mitochondrial dysfunction-related gene TOMM40 and cell adhesion-related gene NECTIN2 further suggest that AD may not only be a neurodegenerative disease but also involve disruptions in peripheral metabolism and the immune system.

The discovery of these genes underscores the multifaceted pathological mechanisms of AD, suggesting that its pathology is not confined to the central nervous system. Pathological changes in peripheral tissues, such as the liver and blood vessels, may also play important roles in disease progression. GO and KEGG pathway analyses further validated the central roles of pathways like cholesterol metabolism and immune responses in AD-related phenotypes, especially highlighted by the significant enrichment results in brain regions such as the anterior cingulate cortex. These findings provide new perspectives on understanding the complex pathological mechanisms of AD and offer potential targets for future personalized treatments.

### Performance and interpretability of AD-MIF

In this study, the proposed AD-MIF model demonstrated significant advantages in integrating multimodal data to predict AD-related tasks. AD-MIF enhances the model’s generalization capability and prediction accuracy by combining raw omics features with omics neighborhood features, efficiently extracting and integrating these features using autoencoders and graph autoencoders. Compared to traditional machine learning methods (such as SVM, LR, RF), deep learning methods (such as ResNet, RNN), and modality imputation-based deep learning methods (such as DeepGAMI and GenNet), AD-MIF achieved higher AUC scores in predicting multiple AD-related tasks, particularly excelling in handling high-dimensional and sparse data. This indicates that AD-MIF can effectively capture complex feature information, supporting early screening and personalized treatment of AD.

To improve the model’s interpretability and uncover the biological significance of genes in prediction tasks, AD-MIF incorporates an attention mechanism. By calculating the attention weights of each gene using Gen-G, the model gains a better understanding of their relative contributions to different phenotype prediction tasks. The gene importance score analysis in Figure 6 reveals that genes related to cognitive levels include ADRA1A, CAMKK1, and TRAPPC4, which are involved in neural transmission, regulation of cognitive functions, and protein transport; genes associated with CERAD scores include SLC30A4, KLHL5, and UCHL1, primarily involved in zinc ion transport and protein degradation; key genes for Braak stages, such as PML, WASF2, and CNTN3, are mainly related to neuroprotection and synapse formation; and genes related to the APOE genotype, including CSRNP2, OXTR, and IRAK4, are involved in the regulation of social behaviors and inflammatory responses. Notably, POLR2C and TRAPPC4 exhibit high associations across multiple phenotypes, suggesting their potentially central roles in the pathogenesis of AD. POLR2C, as a subunit of RNA polymerase II, regulates gene expression related to neural functions and neuroprotection. TRAPPC4 plays a crucial role in protein transport and vesicle formation, maintaining protein homeostasis and synaptic transmission within neurons. The multi-phenotype associations of these two genes suggest that gene expression regulation and protein metabolism may act synergistically in AD, deepening our understanding of the complex molecular mechanisms of AD.

AD-MIF not only showcases exceptional predictive capabilities in AD research but also serves as a powerful tool for understanding the disease’s multifaceted pathological mechanisms. Through gene importance score analysis, the model identifies key candidate genes associated with immunity, metabolism, and neuroinflammation, providing directions for future research. These findings can further validate the biological functions of these genes and explore their potential as therapeutic targets.

### Complementary Roles of PrediXcan and AD-MIF in Phenotypic Prediction

In this study, based on the ROSMAP dataset, we predicted the cognitive diagnosis phenotype and performed enrichment analyses of the relevant important genes using both the individual-level PrediXcan and the deep multi-layer information fusion model AD-MIF. The results revealed significant overlaps between the two methods in the following key pathways and tissue categories: GO Component: GO:0005622 -Intracellular anatomical structure, as well as multiple tissue categories including BTO:0000042 - Animal, BTO:0000142 - Brain, BTO:0001484 - Nervous system, and BTO:0001489 - Whole body. These overlaps indicate that PrediXcan and AD-MIF exhibit high consistency in identifying genes associated with intracellular anatomical structures and multiple key tissue systems, particularly highlighting the enrichment of genes in the brain and nervous system. This underscores the importance of these genes in the regulation of cognitive functions.

Furthermore, the enrichment analysis revealed the unique strengths of each method, demonstrating their complementarity. Individual-level PrediXcan showed significant gene enrichment in multiple organelle-related GO components (such as GO:0043231, GO:0005829, GO:0043229, GO:0043227, and GO:0043226) as well as in UniProt Keywords, including “Alternative splicing” (KW-0025), “Phosphoprotein” (KW-0597), and “Acetylation” (KW-0007). This highlights PrediXcan’s unique advantages in gene expression regulation and protein modification processes. On the other hand, AD-MIF demonstrated significant gene enrichment in more refined cellular substructure categories, such as GO:0005654 - Nucleoplasm and GO:0031981 - Nuclear lumen. This suggests that AD-MIF’s multi-layered information fusion capability can capture more nonlinear and complex gene interactions, identifying key genes and pathways that PrediXcan might not detect.

This complementarity indicates that the combined use of PrediXcan and AD-MIF can more comprehensively elucidate the molecular mechanisms underlying cognitive diagnosis phenotypes. By fully leveraging the strengths of each method, the depth and breadth of the research are enhanced, providing a more holistic understanding of the molecular underpinnings of cognitive functions in AD.

### Drug Repositioning Potential

In this study, through drug repositioning analysis, we identified a series of drugs that may have therapeutic potential for AD. For example, Sirolimus and Dasatinib demonstrated significant efficacy across multiple AD-related phenotypes. Sirolimus may slow neuroinflammation by modulating immune responses, while Dasatinib shows therapeutic potential by clearing senescent cells, thereby alleviating neurodegenerative changes and associated emotional symptoms. These drugs may mitigate disease progression, cognitive decline, and emotional symptoms by specifically regulating the expression of AD-related genes.

The identification of drugs such as Nintedanib, Quizartinib, and MK-2206 further suggests the potential of non-traditional medications in AD treatment. These drugs may exert neuroprotective effects through mechanisms such as reducing inflammatory responses, inhibiting RIPK1 signaling, and regulating cell survival. Although these medications have not been widely used for AD treatment, their successful application in other indications provides a basis for their potential in AD. The drug repositioning results of this study emphasize that personalized drug interventions, based on specific genetic backgrounds, may be more effective.

### Comprehensive Improvements in Cognition, Inflammation, and Neuropathology by MGCD-265 and Dasatinib in the SAMP8 Model

This study demonstrates that both MGCD-265 and Dasatinib significantly improve cognitive function in SAMP8 mice. Results from the novel object recognition and Morris water maze tests indicate that both drugs reduced escape latency and increased the number of platform crossings, suggesting their potential value in restoring spatial memory. Furthermore, immunofluorescence and multiplex cytokine assays revealed that drug treatment markedly suppressed the abnormal activation of astrocytes and microglia in the hippocampal region, while also reducing the excessive release of inflammatory mediators such as IFN-γ and IL-1β. These findings suggest that MGCD-265 and Dasatinib effectively alleviate neuroinflammation.

At the neuropathological level, drug intervention improved neuronal injury, partially restored the number of Nissl bodies, and effectively reduced both abnormal tau phosphorylation and Aβ deposition. Collectively, these results support the notion that MGCD-265 and Dasatinib promote cognitive function recovery by modulating neuroinflammation and neuropathological alterations, providing robust experimental evidence for their potential application in precision therapy for AD.

### Limitations

Although this study achieved significant findings by integrating multi-tissue Transcriptome-Wide Association Studies (TWAS) and deep learning techniques, it also has several limitations. First, while the TWAS method can reveal associations between gene expression and diseases, it cannot directly establish causality. Therefore, experimental validation is required to further confirm the functional impacts of these genes. Second, the effectiveness of the multimodal fusion algorithm depends on the quality of the input data. Future research should incorporate more high-quality multi-omics data to validate these findings. Additionally, the results of the drug repositioning analysis need to be further validated through clinical trials to ensure the efficacy and safety of these drugs in practical applications.

### Future Directions

Future research should focus on several key areas to advance our understanding of AD. First, experimental validation of significant genes is needed to establish causal relationships and clarify their roles in AD pathogenesis. Second, multimodal fusion algorithms can be enhanced by incorporating additional omics data, such as metabolomics and proteomics, to provide a more comprehensive understanding of AD’s complex mechanisms. Finally, large-scale clinical trials are essential to validate the efficacy of potential therapeutic drugs, with particular attention to the interactions between these drugs and key genes like APOE, ensuring targeted and effective treatments. Addressing these areas will help deepen our understanding of AD’s molecular mechanisms and contribute to the development of more effective, personalized treatments.

## Conclusion

This study elucidates the complex genetic mechanisms of AD through a multi-tissue transcriptome-wide association study (TWAS) combined with a multi-modal deep learning method, AD-MIF. Our findings highlight the critical roles of several genes, including APOE and APOC1, in key processes such as cholesterol metabolism, immune response, neuroinflammation, and the regulation of emotional, cognitive, and behavioral functions. Furthermore, the results suggest that AD may extend beyond neurodegenerative pathology, involving abnormalities in peripheral tissues. By integrating multi-modal data, the AD-MIF model greatly enhanced the feature extraction and predictive power of AD-related genes. This approach not only improved the prediction accuracy of multiple AD-related tasks but also provided valuable insights for the development of personalized treatment strategies. Through drug repositioning analysis, we identified several promising drug candidates with potential therapeutic effects, which were further validated in the SAMP8 AD mouse model using comprehensive behavioral, molecular, and pathological assessments. Notably, both MGCD-265 and Dasatinib were found to significantly improve cognitive function, alleviate neuroinflammation, and reduce tau hyperphosphorylation and Aβ deposition. Future research should focus on refining the AD-MIF model and validating the therapeutic effects of these key genes and candidate drugs in clinical settings to enable more precise intervention strategies.

## Methods

### Data Sources and Preprocessing

#### GWAS Summary Statistics

This study utilized GWAS summary statistics obtained from Jiang et al., who analyzed UK Biobank data using a generalized linear mixed model (GLMM) method, such as fastGWA-GLMM [23, 41, 42]. These data underwent rigorous quality control and phenotype standardization, covering 61 traits in areas including mental disorders, neurological diseases, sleep patterns, and metabolic conditions, with all samples of European ancestry. The sample sizes ranged from hundreds to hundreds of thousands of individuals, ensuring the breadth and statistical significance of the analysis (see Appendix 1 for detailed information). The data preprocessing involved the following main steps:

1. **Coordinate Conversion**: Due to differences between versions of the genome reference sequences, the GWAS data were first converted from human genome version 19 (hg19) to the latest reference version (hg38) to ensure data consistency and compatibility with modern genomic resources.
2. **Data Imputation**: To handle missing values in the GWAS data, genotype data from the 1000 Genomes reference panel were used in combination with imputation tools. We performed imputation of missing variants using a 100kb sliding window approach and utilized standardized dosage information to enhance the accuracy of the imputation.
3. **Variant Filtering**: Only variant sites that met quality control standards were retained, including variants with a minor allele frequency (MAF) > 0.01 and a genotype call rate > 0.8, to ensure high data quality and reliability.
4. **Ancestry Matching**: To reduce biases due to population structure, the data analysis was restricted to samples of European ancestry.

This series of processing steps ensured the high quality of the GWAS data, provided a reliable foundation for subsequent analyses, and helped enhance the signal detection capabilities related to the target phenotypes.

#### GTEx Dataset

This study utilized gene expression prediction models for 49 different tissues from the GTEx dataset [43]. These models were based on eQTL models constructed using the mashr method in the GTEx dataset. Each tissue-specific model provides predictions of gene expression levels in the corresponding tissue.

#### ROSMAP Dataset

This study utilized data from the Religious Orders Study and the Memory and Aging Project (ROSMAP), managed by the Rush AD Center in Chicago. ROSMAP is a longitudinal clinical-pathological study specifically focused on AD and its related pathologies. The dataset includes genotype data for 22 autosomes and gene expression data from the dorsolateral prefrontal cortex, ensuring that each subject’s genotype and gene expression data are matched for analysis. Additionally, detailed phenotype data were collected, including sex, education level, ethnicity, APOE genotype, age at death, MMSE scores, pathological scores, and cognitive diagnoses. These phenotype data provide valuable background information for gene-phenotype association analyses, contributing to a deeper understanding of the functions of AD-related genes [44].

In the data preprocessing phase, the following steps were performed:

**1. Genotype Data Filtering:** Sites that are not single nucleotide variants (SNVs) or have multiple alternate alleles were excluded to ensure data quality.
**2. SNP Matrix Conversion:** The filtered genotype data were converted into an SNP matrix for subsequent association analyses.
**3. SNP Annotation:** SNPs were annotated using the UCSC hg19 reference genome and the dbSNP137 database. The RSIDs for each SNP were extracted and added to the information table.
**4. Gene Annotation:** Gene chromosomal locations, gene IDs, and types were obtained from the Ensembl database, with genes on sex chromosomes filtered out to retain only autosomal gene annotation data.

All data underwent stringent quality control, and the analyses were based on resources provided by the AD Knowledge Portal.

#### Drug-induced gene expression profile data

The drug-induced gene expression profile data were sourced from the LINCS L1000 project [45], which is part of the Connectivity Map (CMap) dataset and focuses on the gene expression impacts of drugs and small molecule compounds. LINCS L1000 utilizes a gene expression measurement technique that captures expression values for 978 landmark genes and infers the expression of the entire genome through computational modeling. These data are used to analyze the gene expression effects of drugs, explore potential molecular mechanisms, and conduct drug repositioning studies.

In the data preprocessing phase, the following steps were performed:

**1. Data Filtering and Parsing:** Sample IDs with treatment types labeled as “trt_cp” (drug treatment) were selected from the raw data.
**2. Expression Statistics Parsing:** The cmapR package was used to parse the data, extracting and saving the expression statistics matrix for subsequent analyses.

#### Clinical trial drugs and indications data

To validate the effectiveness of the drug repositioning results, we utilized data from three drug sources: (1) This dataset includes antiepileptic, antiparkinsonian, psychotropic, antipsychotic, and antidepressant/anxiolytic drugs [46]. (2) MEDI-HPS: A dataset containing high-precision, validated drug indication information [47]. (3) ClinicalTrials.gov: Clinical trial drug lists provided by ClinicalTrials.gov. These data served as the foundation for drug enrichment testing and were used to combine p-values for different tissues using the Fisher and Tippett methods [48].

### Association between genetic prediction gene expression level and 61 brain-related characteristics

In this study, we employed the S-PrediXcan method using GWAS summary statistics provided by Jiang et al., in combination with data from the UK Biobank, to investigate the associations between 61 brain-related traits and genetically predicted gene expression levels. These GWAS summary statistics were generated using a generalized linear mixed model (GLMM) approach, such as fastGWA-GLMM. We applied gene expression prediction models from 49 different tissues in the GTEx dataset to these data and inferred gene expression levels in each tissue using the S-PrediXcan method.

For each tissue, we calculated the associations between predicted gene expression and trait risks using the prediction models, generating corresponding p-values. Based on these results, we conducted visual analyses of correlations and pleiotropy findings. We performed multiple comparison corrections on the p-values from the Transcriptome-Wide Association Studies (TWAS) of the 61 brain traits across the 49 tissues, generating corresponding q-values to identify significantly associated gene expression data.

These significant gene expression associations were then visualized using various heatmaps and normalized significance count methods to better understand the pleiotropy and correlations of genes across different brain traits. This comprehensive approach allowed us to elucidate the complex relationships between genetically predicted gene expression and a wide range of brain phenotypes, providing deeper insights into the multifaceted genetic architecture underlying AD and related cognitive functions.

### Training of gene expression prediction model and database construction

Genotype data from 553 samples of autosomal chromosomes and gene expression data from the dorsolateral prefrontal cortex were utilized within the ROSMAP project to train gene expression prediction models. The PredictDB method was adopted to convert the trained models into a format compatible with the PrediXcan software suite. Following the GTEx protocol and considering the sample size of over 350, 60 PEER factors were incorporated into the models. The model training steps included:

**1. Data Preparation:** Residuals were calculated based on PEER factors, utilizing gene expression files and covariate files for data preparation.
**2. Model Training and Database Construction:** Models were trained using the PredictDB method, and the results were stored in an SQLite database for subsequent analyses.

### Model Training and PrediXcan Analysis at Individual Level

After training the gene expression prediction models and constructing the database, the individual-level PrediXcan method was employed to predict gene expression levels based on these models. Associations with target phenotypes were evaluated using the available genotype datasets. The analysis steps for individual-level PrediXcan included:

**1. Gene Expression Prediction:** Gene expression levels of individuals in the cohort were predicted using the trained models.
**2. Phenotype Association Analysis:** Association analyses were performed between the predicted gene expression and target phenotypes (such as APOE genotype, age at death, pathological scores, and cognitive diagnoses) to assess the impact of gene

expression on these traits.

### Deep Multi-layer Information Fusion Model AD-MIF

In response to the multi-modal feature data obtained from ROSMAP samples, this study further proposes a deep multi-layer information fusion model, AD-MIF. This method first preprocesses the AD genotype data to alleviate the challenges posed by the high dimensionality and feature sparsity of genotype data. During the data preprocessing phase, the experiment constructs a multi-branch multilayer perceptron to efficiently extract the genotype data features 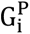:

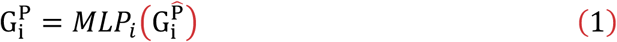

Where 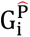 denotes the initial genotype data on the *i*-th chromosome, and *MLP_i_* represents the multilayer perceptron corresponding to the *i*-th chromosome.

High-dimensional features lead to a surge in the number of parameters in deep learning models, while sparsity limits the model’s ability to mine low-order correlations between features, thereby affecting prediction accuracy. For each type of modality data, considering the multiple interaction relationships between neighborhoods, AD-MIF uses Euclidean distance to identify the top K most similar feature data for each gene or SNP as its neighborhood. Based on this, a K-NN gene/SNP neighborhood feature graph representation (Gen-G / SNP-G) is constructed. Subsequently, AD-MIF employs Autoencoders (AE) and Graph Autoencoders (GAE) to extract latent feature information from the original omics features and the omics neighborhood feature graphs, respectively. To integrate these latent feature representations, AD-MIF combines the feature information through a linear combination to obtain a more comprehensive latent representation:

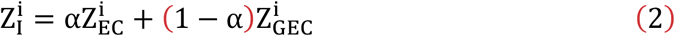

Where α denotes the learnable feature weight, initialized to 0.5.

Finally, to better utilize both the original omics features and the neighborhood features, AD-MIF introduces an information fusion module. This module aims to integrate the specific and shared features learned from the autoencoders at both local and global levels, thereby generating highly consistent feature representations. The local and global feature representations are combined through element-wise summation to obtain the final fused representation:

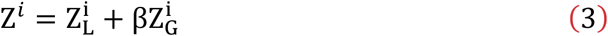

Where 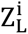 and 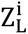 represent the local and global feature representations, respectively, and β is the feature weight parameter.

The model accomplishes various downstream AD phenotype prediction tasks to validate the application value of AD-MIF in multiple AD-related tasks. The architecture of AD-MIF is illustrated in Figure 8, and the detailed experimental procedures can be found in the Supplementary Methods. AD-MIF employs deep learning techniques to perform deep fusion of multimodal data, extracting more representative features to enhance prediction accuracy.

**Figure 8.**
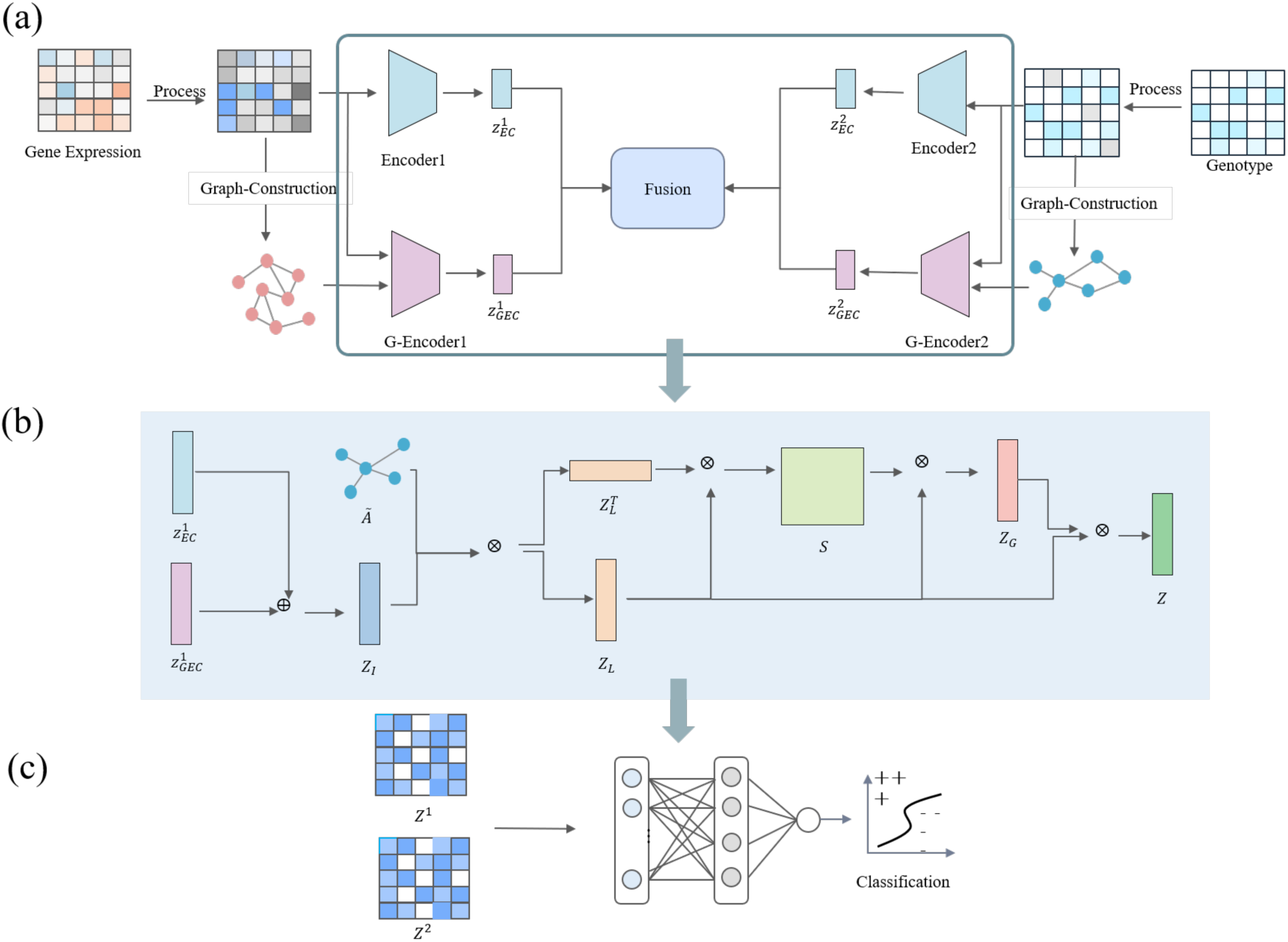
AD-MIF architecture diagram. a) Initialization of gene features and SNP features, and the construction process of neighborhood feature graphs; b) Information fusion process (using genes as an example); c) Prediction based on different downstream AD tasks.

### Gene-based and Gene-set Analyses

Based on the correlations and pleiotropic findings displayed in the charts, features with a greater number of significant genes were selected from the S-PrediXcan results, including irritability, worry/anxiety, father’s disease (AD/dementia), mother’s disease (AD/dementia), and siblings’ disease (AD/dementia). Additionally, by combining the individual-level PrediXcan analysis results using ROSMAP data, enrichment analyses were conducted on these significant genes. These analyses provided further insight into how genes regulate the occurrence and development of these traits in different tissues. The specific methods are as follows:

**1. Gene-Level Enrichment Analysis of S-PrediXcan Results:** Enrichment analyses were performed across multiple tissues for each trait. By integrating gene expression levels and q-values, genes with significant regulatory roles in specific tissues were identified.
**2. Enrichment Analysis of AD Family History Traits:** For traits related to family history of AD (such as the history of AD or dementia in fathers, mothers, and siblings), similar enrichment analyses were conducted to further examine the distribution of significant genes across different tissues.
**3. Enrichment Analysis of Individual-Level PrediXcan Phenotype Association Results:** Based on the AD-related phenotype association analysis results fro m ROSMAP data, enrichment analyses were performed to identify significant genes and regulatory pathways associated with AD. These analyses provide a better understanding of the potential roles of genes in the pathogenesis of AD and lay the groundwork for subsequent functional validations and the discover y of potential drug targets.

### Drug Repositioning via Gene Expression Correlation

This study is based on the method developed by Hon-Cheong So et al., which explores potential drug repositioning candidates by comparing disease-related gene expression changes with drug-induced gene expression profiles. Spearman and Pearson correlation coefficients were utilized to assess the correlation between gene expression profiles inferred from GWAS summary statistics and drug-induced expression profiles provided by CMap. Specifically, the Spearman correlation coefficient was used to evaluate the rank-order correlation between the two variables, while the Pearson correlation coefficient was employed to assess the linear relationship between them.Subsequently, the Kolmogorov-Smirnov test was performed to determine whether the genes that are upregulated and downregulated in the disease state exhibit similar expression changes under drug treatment. This test compares the differences between two empirical cumulative distribution functions to evaluate whether the drug can reverse the disease-related gene expression patterns.

Additionally, drug repositioning analysis was conducted on the significant genes identified in the phenotype association results of the APOE genotype. By comparing the correlation between the gene expression profiles associated with the APOE genotype and the drug-induced expression profiles, candidate drugs with potential therapeutic effects on APOE-related pathological processes were further screened. This series of analytical procedures facilitated the systematic identification of potential therapeutic drugs for AD by examining gene expression correlations across multiple dimensions. These procedures established a comprehensive approach for drug repositioning and precision treatment strategies based on the observed gene expression patterns.”

### Drug Set Repositioning Enrichment Analysis Method

To evaluate the enrichment of known drug sets in transcriptome-wide association analyses, two primary testing methods were employed:

**1. Self-Contained Test:** This test assesses the significance of specific drug sets. A one-sample t-test was used to determine whether the z-scores of the target drug sets significantly differed from zero.
**2. Competitive Test:** This test evaluates significance by comparing the rankings of drugs within the set to those outside the set. A two-sample t-test was performed to compare the differences between the target drug sets and randomly selected drug sets.

For control analyses, an equal number of drugs were randomly selected from the dataset to generate random drug sets. For each disease and brain region dataset, one-sided t-tests were conducted, and the p-values were converted to z-scores using the Probit function. Meta-analyses of p-values from different tissues were then performed using the Fisher method and Tippett’s minimum p-value method to integrate results from multiple independent experiments. These methods assess the significance of drug sets by combining multiple p-values. To control for multiple testing issues, False Discovery Rate (FDR) correction was applied using the Benjamini-Hochberg method. A significance threshold of 0.05 was set to ensure the reliability of the analysis results. This analytical workflow enables the effective evaluation of potential therapeutic roles of drug sets in AD and other complex disease studies.

### In Vivo Evaluation of Candidate Drugs in the SAMP8 Alzheimer’s Murine Model

Based on the drug repurposing analysis, the top-ranked candidates—MGCD-265 and dasatinib—were selected for in vivo validation in the SAMP8 murine model of AD. These drugs were comprehensively evaluated using molecular techniques, including Western blotting, cytokine assays, and immunofluorescence, alongside behavioral assessments such as novel object recognition and the Morris water maze tests. This integrated approach allowed for a systematic demonstration of improvements in both pathological markers and cognitive function. The combined findings provide compelling experimental evidence supporting the potential application of these candidate drugs in precision therapeutics for AD.

### Animals

Male SAMR1 and SAMP8 mice (6 months old) were obtained from Beijing Z hishan Co., Ltd. (Beijing, China). The mice were housed under standard labora tory conditions, with 3-5 animals per cage, at a controlled room temperature of 22 ± 2°C and 55 ± 5% relative humidity, on a 12-hour light/dark cycle. All animal procedures adhered to Institutional Guidelines and complied with the European Communities Council Directive (86/609/EEC) regulations.

### Group and treatment

Dasatinib and MGCD-265 were purchased from Selleck Chemicals LLC (Shanghai, China). Following a 7-day acclimation period, SAMR1 mice were assigned as the blank control group, while SAMP8 mice were randomly allocated into three experimental groups: (1) model group, (2) dasatinib-treated group, and (3) MGCD-265-treated group.

The SAMR1 control group was housed under standard conditions and received oral gavage of a vehicle solution. Similarly, the SAMP8 model group was maintained under normal conditions with vehicle administration. The SAMP8 dasatinib-treated group received daily oral administration of dasatinib (10 mg/kg/day), while the SAMP8 MGCD-265-treated group was administered MGCD-265 (25 mg/kg/day) via oral gavage. All treatments were continued for 7 consecutive weeks.

### NOR test

The novel object recognition (NOR) test was conducted following standardized procedures, comprising three phases. In the Habituation Phase, individual mice were acclimated to the testing environment in a 45 × 45 cm black opaque open-field arena for 10 minutes, allowing them to become familiar with the apparatus without objects present. In the Training Phase (24-hour interval), two identical objects, matched in all visual and tactile characteristics, were placed at symmetric locations within the arena. Each mouse was then allowed to explore these objects freely for 10 minutes to establish object familiarity. In the Testing Phase (1-hour interval), one familiar object was replaced with a novel object exhibiting distinct shape, color, and size characteristics. Mice were reintroduced to the arena for a 5-minute test session, during which exploratory behavior was video-recorded and analyzed using the Specify software (Shanghai Xinsoft Information Technology Co., Ltd) tracking system. Exploration was defined as directed sniffing or touching of objects with the nose or forepaws while oriented toward the object. The discrimination index was computed as the ratio of time spent investigating the novel object relative to the total exploration time of both objects.

### MWM test

Following 7 weeks of treatment, spatial memory was evaluated using the Morris water maze (MWM) paradigm. After a 24-hour environmental acclimation period in the testing facility, mice underwent five consecutive days of acquisition training, consisting of four daily trials (60 sec/trial) with randomized quadrant entry points to locate a submerged platform. On day 6, a 60-second probe trial was conducted with the platform removed. During this trial, swimming patterns were recorded and analyzed using video tracking software (Shanghai Xinsoft Information Technology Co., Ltd) to quantify spatial memory retention through parameters such as platform crossings and quadrant occupancy time.

### Western Blot

After extraction from the skull, the hippocampi were immediately dissected on ice, and the total protein concentration was determined using a BCA assay kit (Shanghai Epizyme Biomedical Technology Co., Ltd). For immunoblotting, 30 μg of total protein per sample was separated by 10% SDS-PAGE and transferred to PVDF membranes. The membranes were blocked with 5% BSA in TBST for 2 hours at room temperature, followed by overnight incubation at 4°C with primary antibodies against Phospho-Tau (Thr231) (Cell Signaling Technology, 7129S), Phospho-Tau (Ser396) (abcam, ab32057), total Tau (Cell Signaling Technology, 46687S), GSK-3β (Cell Signaling Technology, 12456S), CDK5 (Cell Signaling Technology, 2506S), PP2Aα/β (abcam, ab32065), and β-Actin (Proteintech, 20536-1-AP). Protein bands were visualized after incubation with an HRP-conjugated goat anti-rabbit IgG secondary antibody (Proteintech, SA00001-2).

### Measurement of cytokines

Cytokine levels in hippocampal tissue and brain homogenates were measured using the Cytokine & Chemokine 26-Plex Mouse ProcartaPlex™ (EPX260-26088-901, ThermoFisher), which includes GM-CSF, IFN γ, IL-1 β, IL-2, IL-4, IL-5, IL-6, IL-12p70, IL-13, IL-18, TNF α, IL-9, IL-10, IL-17A, IL-22, IL-23, IL-27, CCL11, GRO α, IP-10, MCP-1, MCP-3, MIP-1 α, MIP-1 β, MIP-2, and CCL5. Standard curves were calculated using parameter logistic regression, and cytokine fluorescence intensity was detected using the Luminex machine.

### Immunofluorescence assay

Brain tissue histopathology and immunofluorescence analyses were performed following standard operating procedures for reagents and antibodies. Briefly, whole brains were paraffin-embedded and sectioned into thin slices for subsequent staining.

The primary reagents and antibodies used included hematoxylin and eosin (H&E) stain, Nissl stain, AT8 antibody (Thermo Fisher, MN1020), GFAP antibody (Servicebio, GB21301), IBA1 antibody (Servicebio, GB12105), Thioflavin S (Shanghai Yuanye Bio-Technology Co., Ltd, S19293), and NeuN antibody (HuaBio, ET1602-12).

### Statistical analysis

All quantitative data are reported as means with standard deviations. Group comparisons were analyzed using parametric (ANOVA, either one-way or two-way) or non-parametric (Kruskal-Wallis) approaches, followed by appropriate post-hoc tests (Tukey’s for parametric, Dunn’s for non-parametric analyses). Statistical computations were performed using GraphPad Prism.

## Data availability

The GWAS summary statistics for 2,989 binary traits analyzed by Jiang et al. using UKB data can be accessed at http://fastgwa.info/ukbimpbin and the GWAS Catalog (GCP ID: GCP000224). The GTEx data is publicly available through the GTEx Portal (https://gtexportal.org). Gene expression, genotype, and phenotypic data from the dorsolateral prefrontal cortex were provided by the Rush Alzheimer’s Disease Center, Rush University Medical Center, Chicago. Data collection was supported by National Institute on Aging (NIA) grants P30AG10161 (ROS), R01AG15819 (ROSMAP; genomics and RNAseq), R01AG17917 (MAP), and U01AG46152 (ROSMAP AMP-AD). Additional phenotypic data can be requested at www.radc.rush.edu. The LINCS L1000 Level 5 data is accessible via NCBI GEO (https://www.ncbi.nlm.nih.gov/geo/query/acc.cgi?acc=GSE70138). The MEDI resource can be found at https://www.vumc.org/cpm/center-precision-medicine-blog/medi-ensemble-medication-indication-resource. A list of drugs included in clinical trials (according to ClinicalTrials.gov) is available at https://doi.org/10.15363/thinklab.d212. The Anatomical Therapeutic Chemical (ATC) Classification from KEGG can be accessed via http://www.genome.jp/kegg-bin/get_htext?br08303.keg. Lastly, the MetaXcan tool is available at https://github.com/hakyimlab/MetaXcan.

The key results of this study, including gene expression predictions, significant trait associations, and drug repositioning outcomes, are available at https://drive.google.com/drive/folders/1K6qg5YY2nGCXbXfzBr5hg6aKWwowsd2e.

## Code availability

The code used for the analyses in this study is available at the following GitHub repository: https://github.com/LearnDrugAI/Multi-Tissue_Analysis_DL_AD.

## Acknowledgements

The authors express their deep gratitude to the patients and their families for their generous contributions to this research. This study utilized data obtained from the AD Knowledge Portal (https://adknowledgeportal.org), specifically gene expression, genotype, and phenotypic data from the dorsolateral prefrontal cortex, provided by the Rush Alzheimer’s Disease Center at Rush University Medical Center, Chicago. Data collection was supported by grants from the National Institute on Aging (NIA), including P30AG10161, R01AG15819, R01AG17917, and U01AG46152. The vital role of research volunteers and the valuable contributions of collaborating researchers are also acknowledged. Additionally, this work was supported by the scientific and technological project “Research and Development of Key Technologies for the Production and Quality Control of Sipaiyi Intelligent Preparations,” led by Xiaoyi Lv in 2023.

