## supplementary information for "Integrating GWAS and Transcriptomic Data Using PrediXcan and Multimodal Deep Learning Reveals Genetic Basis and Drug Repositioning Opportunities for Alzheimer’s Disease" (from another version of this preprint: version 1, 2025-01-02)

#### Supplementary Tables

1. Supplementary Table S1 is provided as an **Excel file** and includes detailed GWAS summary data for 61 brain and psychiatric-related traits.
2. Supplementary Table S12, S3, and S4 represent Gene-Trait Associations Among Irritability, Anxiety, and Alzheimer's Risk Factors at q-Value Thresholds of 0.5, 0.1, and 0.05, respectively.
3. Supplementary Table S5 is provided as an **Excel file** and includes Gene Information related to AD for Brain and Psychiatric-related Traits with a q-Value Threshold of 0.05.
4. Supplementary Table S6 contains annotation information for AD-related phenotypes analyzed using individual-level PrediXcan methodology. The analysis is based on gene expression and genotype data from 553 dorsolateral prefrontal

cortex samples in the ROSMAP database, aiming to associate predicted gene expression levels with Alzheimer's disease (AD) phenotypes.

5. Supplementary Table S7 is provided as an **Excel file** and includes Gene Information Related to Phenotypes Predicted by the Individual-level PrediXcan Method.
6. Supplementary Table S8 represents the Enrichment analysis of phenotype-related genes in cognitive diagnosis
7. Supplementary Table S9 represents the Enrichment of Hub Genes Associated with Cognitive Diagnostic Phenotypes Predicted by the AD-MIF Model
8. Supplementary Table S10 represents the Top Repositioning Hits for Father's Alzheimer's Disease/Dementia (ranked by Q-values and P-values).
9. Supplementary Table S11 represents the Top Repositioning Hits for Siblings 's Alzheimer's Disease/Dementia (Ranked by Q-values and P-values)
10. Supplementary Table S12 and Table S13 represent the Top Repositioning Hits for Irritability and the Top Repositioning Hits for Worrier/Anxious Feelings, respectively, ranked by Q-values and P-values.
11. Supplementary Table S14 represents the Drug enrichment analyses P values with drug-sets defined by MEDI-HPS.
12. Supplementary Table S15 represents the Drug enrichment analyses P values with drug-sets defined by ClinicalTrials.

#### Supplementary Figures

1. Supplementary Figure S1 stands for Standardized Counting Results of Brain and Psychological Traits: 18 Remaining after screening from 61.
2. Supplementary Figure S2 represents the Distribution of Significant Genes for Five Brain and Psychiatric Traits (Including AD Family History and Emotional Traits) Across 49 Tissues Identified via S-PrediXcan Analysis.
3. Supplementary Figure S3 represents the Gene Expression Network for Brain- and Psychiatric-Related Traits at a q-value Threshold of 0.5.
4. Supplementary Figure S4 represents the Gene Association Network of Five Significant Traits with Q Threshold of 0.5.
5. Supplementary Figure S5 represents the Gene Expression Network for Brain- and Psychiatric-Related Traits at a q-value Threshold of 0.1.
6. Supplementary Figure S6 represents the Shared Gene Network between Alzheimer's Disease-Related Phenotypes.
7. Supplementary Figure S7 represents the Cellular Component (Gene Ontology) Enrichment Analysis of Genes Associated with Cognitive Diagnosis (similarity ⩾ 0.8).
8. Supplementary Figure S8 represents the Cellular Component (Gene Ontology) Enrichment Analysis of Hub Genes Associated with Cognitive Diagnostic Phenotypes Predicted by the AD-MIF Model (similarity ⩾ 0.8).
9. Supplementary Figure S9 represents the Intersection Analysis of Common Drugs and Tissues Across AD Family History Traits: Siblings, Father and Mother.
10. Supplementary Figure S10 represents the Top 50 Repurposed Drugs for Illnesses of Father - Alzheimer's Disease/Dementia Based on Q-value and P-value in Adrenal Gland.
11. Supplementary Figure S11 represents the Top 50 Repurposed Drugs for Illnesses of Siblings - Alzheimer's Disease/Dementia Based on Q-value and P-value in Stomach.
12. Supplementary Figure S12 and Supplementary Figure S13 represent the Top 50 Repurposed Drugs for Irritability Based on Q-value in Significant Tissues and the

Top 50 Repurposed Drugs for Worrier/Anxious Feelings Based on Q-value in Significant Tissues, respectively.

13. Supplementary Figure S14 represents the Father's Alzheimer's Disease: Trait-Tissue-Gene-Drug Interaction Network.
14. Supplementary Figure S15 represents the Network of Siblings' Alzheimer's Disease with Tissue-Gene-Drug Interactions for Drug Repurposing.
15. Supplementary Figure S16 represents the Trait-Tissue-Gene-Drug Interaction Network for Irritability.
16. Supplementary Figure S17 represents the Trait-Tissue-Gene-Drug Interaction Network for Worrier/Anxious Feelings.
17. Supplementary Figure S18 represents the Top 50 Repurposed Drugs for Alzheimer's Disease Based on P-value in Dorsolateral Prefrontal Cortex.
18. Supplementary Figure S19 represents the Intersection Analysis of Shared Drugs and Tissues Between Irritability and Worrier/Anxious Feelings.
19. Supplementary Figure S20 represents the Irritability: GO Biological Process and KEGG Pathway Enrichment Analysis Across Significant Tissues.
20. Supplementary Figure S21 represents the Worrier/Anxious Feelings: GO Biological Process and KEGG Pathway Enrichment Analysis Across Significant Tissues.
21. Supplementary Figure S22 represents the Worrier/Anxious Feelings: GO biological process enrichment analysis in other significant tissues.
22. Supplementary Figure S23 represents the Worrier/Anxious Feelings: GO Biological Process and KEGG Pathway Enrichment Analysis in Brain Anterior Cingulate Cortex (BA24) and Artery Aorta (q-value $\leqslant$ 0.01).

### Supplementary Methods

AD-MIF Model Based on Information Fusion: Gene Feature Analysis and Prediction for Alzheimer's Disease.

#### Supplementary Tables

Table S1 GWAS Summary Data for 61 Brain and Psychiatric-related traits. Detailed information is available in Supplementary Table S1.xlsx.

Table S2 Gene-Trait Associations Among Irritability, Anxiety, and Alzheimer's Risk Factors at a q-Value Threshold of 0.5.

| **Gene** | **Shared Status** | **Shared Trait Combination** | **Regulation (1down/2up)** | **Total Edge Width** |
|---|---|---|---|---|
| KAT8 | 4 | FA, IR, MA and W/A | 1 | 429 |
| RP11-196G11.2 | 3 | IR, MA and W/A | 1 | 216 |
| ZNF646 | 3 | IR, MA and W/A | 1 | 174 |
| APOE | 3 | FA, MA and SA | 2 | 168 |
| RP11-196G11.6 | 3 | IR, MA and W/A | 2 | 116 |
| APOC1 | 3 | FA, MA and SA | 1 | 104 |
| VKORC1 | 3 | IR, MA and W/A | 1;2 | 94 |
| PRSS53 | 3 | IR, MA and W/A | 2 | 86 |
| SCIMP | 3 | FA, IR and MA | 2 | 36 |
| BCKDK | 2 | MA and W/A | 1 | 213 |
| HSD3B7 | 2 | IR and W/A | 1 | 186 |
| STX4 | 2 | IR and W/A | 2 | 166 |
| SETD1A | 2 | IR and W/A | 1 | 160 |
| STX1B | 2 | IR and W/A | 1 | 144 |
| NECTIN2 | 2 | FA and MA | 1 | 118 |
| PRSS8 | 2 | MA and W/A | 1 | 112 |
| PRSS36 | 2 | MA and W/A | 1 | 96 |
| RP11-131L12.4 | 2 | IR and W/A | 1 | 96 |
| BORCS7 | 2 | IR and W/A | 1;2 | 94 |
| DOPEY1 | 2 | IR and W/A | 1 | 91 |
| CEACAM19 | 2 | FA and MA | 2 | 86 |
| CLDN23 | 2 | IR and W/A | 1 | 85 |
| MTMR9 | 2 | IR and W/A | 1 | 84 |
| RP11-1072A3.4 | 2 | IR and W/A | 2 | 81 |
| CTA-223H9.9 | 2 | IR and W/A | 2 | 80 |
| CHMP3 | 2 | IR and W/A | 2 | 79 |
| RP11-131L12.3 | 2 | IR and W/A | 1 | 78 |
| NEIL2 | 2 | IR and W/A | 1 | 77 |
| RP11-78O7.2 | 2 | IR and W/A | 1 | 76 |
| MEI1 | 2 | IR and W/A | 2 | 71 |
| AS3MT | 2 | IR and W/A | 1;2 | 70 |
| SLC35G5 | 2 | IR and W/A | 2 | 68 |
| APOM | 2 | MA and W/A | 2 | 67 |
| RWDD2A | 2 | IR and W/A | 1 | 67 |
| DOC2A | 2 | MA and W/A | 2 | 66 |
| ERI1 | 2 | IR and W/A | 2 | 64 |
| NCOA6 | 2 | IR and W/A | 2 | 63 |
| GGT7 | 2 | IR and W/A | 1 | 62 |
| AKAP10 | 2 | IR and W/A | 2 | 58 |

| | | | | |
|---|---|---|---|---|
| SELENOH | 2 | IR and W/A | 1 | 58 |
| WNT3 | 2 | IR and W/A | 2 | 58 |
| BLK | 2 | IR and W/A | 2 | 56 |
| CLPTM1 | 2 | FA and MA | 1;2 | 56 |
| NT5C2 | 2 | IR and W/A | 1;2 | 56 |
| MFHAS1 | 2 | IR and W/A | 1 | 54 |
| LINCR-0001 | 2 | IR and W/A | 1 | 52 |
| ACO2 | 2 | IR and W/A | 2 | 51 |
| FAM167A | 2 | IR and W/A | 2 | 51 |
| PINX1 | 2 | IR and W/A | 1 | 51 |
| PPP1R3B | 2 | IR and W/A | 2 | 50 |
| PTCH1 | 2 | IR and W/A | 2 | 50 |
| ZDHHC5 | 2 | IR and W/A | 1 | 48 |
| ARHGAP27 | 2 | IR and W/A | 2 | 46 |
| CTSB | 2 | IR and W/A | 2 | 46 |
| CYB561D2 | 2 | IR and W/A | 1;2 | 46 |
| FBXL19 | 2 | IR and W/A | 1 | 46 |
| ZFP36L2 | 2 | FA and IR | 1;2 | 46 |
| C18orf8 | 2 | IR and MA | 2 | 45 |
| ZC3H7B | 2 | IR and W/A | 2 | 45 |
| SUDS3 | 2 | IR and W/A | 2 | 44 |
| LINC00493 | 2 | IR and W/A | 2 | 43 |
| MSH5 | 2 | MA and W/A | 2 | 43 |
| PVR | 2 | FA and MA | 1 | 43 |
| AF131215.8 | 2 | IR and W/A | 1 | 42 |
| CELF4 | 2 | IR and W/A | 2 | 42 |
| NDUFS2 | 2 | FA and MA | 1 | 42 |
| SLC25A17 | 2 | IR and W/A | 2 | 42 |
| XRCC6 | 2 | IR and W/A | 1 | 41 |
| NEGR1 | 2 | IR and W/A | 2 | 40 |
| RCN1 | 2 | IR and W/A | 2 | 40 |
| ST3GAL3 | 2 | IR and W/A | 2 | 39 |
| FDFT1 | 2 | IR and W/A | 1 | 38 |
| PAX6 | 2 | IR and W/A | 2 | 38 |
| POLR3H | 2 | IR and W/A | 2 | 38 |
| SOX7 | 2 | IR and W/A | 2 | 37 |
| ST6GALNAC3 | 2 | IR and W/A | 2 | 37 |
| TGOLN2 | 2 | FA and W/A | 1;2 | 35 |
| RP11-297N6.4 | 2 | IR and W/A | 2 | 34 |
| TRAPPC5 | 2 | IR and W/A | 2 | 34 |
| TMEM106B | 2 | IR and MA | 1;2 | 33 |
| CHADL | 2 | IR and W/A | 2 | 32 |
| EP300 | 2 | IR and W/A | 2 | 32 |
| HEXIM1 | 2 | IR and W/A | 1 | 32 |

| | | | | |
|---|---|---|---|---|
| MICB | 2 | MA and W/A | 1 | 32 |
| MMAB | 2 | IR and W/A | 1 | 32 |
| PGM3 | 2 | IR and W/A | 1 | 32 |
| PHF5A | 2 | IR and W/A | 1 | 32 |
| RP11-188D8.1 | 2 | IR and W/A | 1 | 32 |
| TOB2 | 2 | IR and W/A | 2 | 32 |
| DLST | 2 | IR and W/A | 1 | 31 |
| KCTD10 | 2 | IR and W/A | 1;2 | 30 |
| PROX2 | 2 | IR and W/A | 1 | 30 |
| MAP1LC3B | 2 | IR and W/A | 1 | 29 |
| CELF2 | 2 | IR and W/A | 1 | 28 |
| CYP17A1 | 2 | IR and W/A | 1;2 | 28 |
| MAPT | 2 | IR and W/A | 1 | 28 |
| RP11-456K23.1 | 2 | IR and W/A | 2 | 28 |
| PMVK | 2 | IR and W/A | 2 | 27 |
| C17orf107 | 2 | IR and MA | 2 | 26 |
| TIMP4 | 2 | FA and W/A | 1;2 | 26 |
| AREL1 | 2 | IR and W/A | 1 | 24 |
| BIN1 | 2 | FA and MA | 1 | 24 |
| DEFB134 | 2 | IR and W/A | 1 | 24 |
| KCNN3 | 2 | IR and W/A | 1 | 24 |
| NBEAL1 | 2 | FA and IR | 2 | 23 |
| TLR9 | 2 | IR and W/A | 1;2 | 23 |
| UBE3D | 2 | IR and W/A | 2 | 23 |
| LINC00461 | 2 | IR and W/A | 1 | 22 |
| LTBP2 | 2 | IR and W/A | 1 | 22 |
| RP11-10A14.4 | 2 | IR and W/A | 1 | 22 |
| AC078842.3 | 2 | IR and W/A | 2 | 21 |
| L3MBTL2 | 2 | IR and W/A | 2 | 21 |
| RP11-981G7.6 | 2 | IR and W/A | 1 | 21 |
| SPECC1 | 2 | IR and W/A | 1;2 | 21 |
| FBXL5 | 2 | IR and W/A | 2 | 20 |
| FAM66A | 2 | IR and W/A | 1 | 19 |
| DFNA5 | 2 | IR and W/A | 2 | 18 |
| PRDX2 | 2 | SA and W/A | 2 | 18 |
| TAOK2 | 2 | MA and W/A | 2 | 18 |
| YLPM1 | 2 | IR and W/A | 2 | 18 |
| MVK | 2 | IR and W/A | 2 | 17 |
| TCAIM | 2 | IR and W/A | 1 | 16 |
| RP11-10A14.5 | 2 | IR and W/A | 2 | 15 |
| ADAM10 | 2 | FA and MA | 2 | 14 |
| CTD-2561B21.11 | 2 | IR and W/A | 2 | 14 |
| HCG27 | 2 | IR and MA | 1 | 14 |
| PBXIP1 | 2 | IR and W/A | 1 | 14 |

| | | | | |
|---|---|---|---|---|
| PIGU | 2 | IR and W/A | 2 | 13 |
| FCF1 | 2 | IR and W/A | 2 | 11 |
| TRAPPC6A | 2 | FA and MA | 1 | 11 |
| MSR1 | 2 | IR and W/A | 2 | 10 |
| ARHGAP12 | 2 | IR and W/A | 2 | 9 |
| CTC-467M3.3 | 2 | IR and W/A | 1 | 9 |
| CASC10 | 2 | IR and W/A | 1 | 8 |
| ARPP21 | 2 | IR and W/A | 2 | 7 |
| CHST8 | 2 | IR and W/A | 1 | 7 |
| GRM3 | 2 | IR and W/A | 1 | 6 |
| RP11-490G2.2 | 2 | IR and W/A | 2 | 6 |
| RP11-90P13.1 | 2 | IR and W/A | 2 | 6 |

**Abbreviations:** IR: Irritability; W/A: Worrier/Anxious feelings; FA: Father's Alzheimer's/Dementia; SA: Siblings' Alzheimer's/Dementia; MA: Mother's Alzheimer's/Dementia

**TotalEdgeWidth** is a quantitative measure used to assess the overall importance of a gene across all studied traits and tissues. It is calculated by summing the edgeWidth values for each gene across all associated traits, where edgeWidth represents the number of tissues in which the gene shows a strong association with a specific trait. A higher TotalEdgeWidth indicates that the gene has consistent and significant associations across multiple traits and tissues, suggesting it may play a key role in the biological mechanisms underlying these traits. This metric helps prioritize the most influential genes in the study, guiding further biological validation or functional research.

Table S3 Gene-Trait Associations Among Irritability, Anxiety, and Alzheimer's Risk Factors at a q-Value Threshold of 0.1.

| Gene | Shared Status | Shared Trait Combination | Regulation (1down/2up) | Total Edge Width |
|---|---|---|---|---|
| APOE | 3 | FA, MA and SA | 2 | 154 |
| ZNF646 | 3 | IR, MA and W/A | 1 | 140 |
| NECTIN2 | 3 | FA, MA and SA | 1 | 108 |
| ZNF668 | 3 | IR, MA and W/A | 2 | 106 |
| RP11-196G11.6 | 3 | IR, MA and W/A | 2 | 84 |
| TOMM40 | 3 | FA, MA and SA | 1 | 66 |
| STX4 | 2 | IR and W/A | 1 | 164 |
| STX1B | 2 | IR and W/A | 1 | 140 |
| RP11-196G11.2 | 2 | IR and W/A | 1 | 98 |
| RP11-131L12.4 | 2 | IR and W/A | 1 | 92 |
| HSD3B7 | 2 | IR and W/A | 1 | 91 |
| MTMR9 | 2 | IR and W/A | 1 | 84 |
| SETD1A | 2 | IR and W/A | 1 | 80 |
| RP11-1072A3.4 | 2 | IR and W/A | 2 | 78 |
| CLDN23 | 2 | IR and W/A | 1 | 77 |
| KAT8 | 2 | MA and W/A | 1 | 75 |
| SLC35G5 | 2 | IR and W/A | 2 | 67 |
| ERI1 | 2 | IR and W/A | 2 | 62 |
| NEIL2 | 2 | IR and W/A | 1 | 59 |
| CEACAM19 | 2 | FA and MA | 2 | 57 |
| BLK | 2 | IR and W/A | 2 | 56 |
| SELENOH | 2 | IR and W/A | 1 | 56 |
| WNT3 | 2 | IR and W/A | 2 | 56 |
| PRSS53 | 2 | IR and W/A | 2 | 50 |
| PPP1R3B | 2 | IR and W/A | 2 | 49 |
| FBXL19 | 2 | IR and W/A | 1 | 46 |
| LINCR-0001 | 2 | IR and W/A | 1 | 46 |
| FAM167A | 2 | IR and W/A | 2 | 45 |
| PTCH1 | 2 | IR and W/A | 2 | 44 |
| AF131215.8 | 2 | IR and W/A | 1 | 42 |
| BCKDK | 2 | MA and W/A | 1 | 42 |
| DOPEY1 | 2 | IR and W/A | 1 | 40 |
| NCOA6 | 2 | IR and W/A | 2 | 40 |
| RCN1 | 2 | IR and W/A | 2 | 40 |
| DENND1B | 2 | IR and W/A | 1 | 38 |
| PAX6 | 2 | IR and W/A | 2 | 38 |
| SUDS3 | 2 | IR and W/A | 2 | 38 |
| BORCS7 | 2 | IR and W/A | 1;2 | 37 |
| SLC25A17 | 2 | IR and W/A | 2 | 37 |

Formatted Table

| | | | | |
|---|---|---|---|---|
| ARHGAP27 | 2 | IR and W/A | 1;2 | 36 |
| MSRA | 2 | IR and W/A | 2 | 36 |
| ZC3H7B | 2 | IR and W/A | 2 | 35 |
| MFHAS1 | 2 | IR and W/A | 1 | 34 |
| APOC4 | 2 | FA and MA | 2 | 32 |
| RP11-131L12.3 | 2 | IR and W/A | 1 | 31 |
| PROX2 | 2 | IR and W/A | 1 | 28 |
| RP11-78O7.2 | 2 | IR and W/A | 1 | 25 |
| DEFB134 | 2 | IR and W/A | 1 | 24 |
| CHMP3 | 2 | IR and W/A | 2 | 22 |
| AREL1 | 2 | IR and W/A | 1 | 21 |
| NT5C2 | 2 | IR and W/A | 1;2 | 21 |
| ZDHHC5 | 2 | IR and W/A | 1 | 19 |
| FDFT1 | 2 | IR and W/A | 1 | 18 |
| LINC00493 | 2 | IR and W/A | 2 | 18 |
| LTBP2 | 2 | IR and W/A | 2 | 18 |
| YLPM1 | 2 | IR and W/A | 2 | 18 |
| EP300 | 2 | IR and W/A | 2 | 16 |
| FAM66A | 2 | IR and W/A | 1 | 14 |
| L3MBTL2 | 2 | IR and W/A | 2 | 14 |
| RP11-10A14.5 | 2 | IR and W/A | 2 | 12 |
| RP11-981G7.6 | 2 | IR and W/A | 1 | 12 |
| FCF1 | 2 | IR and W/A | 2 | 10 |

Table S4 Gene-Trait Associations Among Irritability, Anxiety, and Alzheimer's Risk Factors at a q-Value Threshold of 0.05.

| Gene | Shared Status | Shared Trait Combination | Regulation (1down/2up) | Total Edge Width |
|---|---|---|---|---|
| APOE | 3 | FA, MA and SA | 2 | 154 |
| APOC1 | 3 | FA, MA and SA | 1 | 96 |
| NECTIN2 | 3 | FA, MA and SA | 1 | 96 |
| RP11-196G11.2 | 2 | IR and W/A | 1 | 98 |
| HSD3B7 | 2 | IR and W/A | 1 | 89 |
| MTMR9 | 2 | IR and W/A | 1 | 84 |
| STX4 | 2 | IR and W/A | 1 | 82 |
| RP11-1072A3.4 | 2 | IR and W/A | 2 | 78 |
| SETD1A | 2 | IR and W/A | 1 | 78 |
| CLDN23 | 2 | IR and W/A | 1 | 77 |
| STX1B | 2 | IR and W/A | 1 | 70 |
| KAT8 | 2 | MA and W/A | 1 | 64 |
| RP11-131L12.4 | 2 | IR and W/A | 1 | 64 |
| ERI1 | 2 | IR and W/A | 2 | 62 |
| SLC35G5 | 2 | IR and W/A | 2 | 61 |
| ZNF646 | 2 | IR and W/A | 1 | 54 |
| BLK | 2 | IR and W/A | 2 | 53 |
| NEIL2 | 2 | IR and W/A | 1 | 53 |
| WNT3 | 2 | IR and W/A | 2 | 53 |
| PPP1R3B | 2 | IR and W/A | 2 | 48 |
| FBXL19 | 2 | IR and W/A | 1 | 46 |
| FAM167A | 2 | IR and W/A | 2 | 43 |
| PTCH1 | 2 | IR and W/A | 2 | 43 |
| AF131215.8 | 2 | IR and W/A | 1 | 42 |
| LINCR-0001 | 2 | IR and W/A | 1 | 41 |
| RCN1 | 2 | IR and W/A | 2 | 40 |
| PAX6 | 2 | IR and W/A | 2 | 38 |
| BCKDK | 2 | MA and W/A | 2 | 34 |
| SLC25A17 | 2 | IR and W/A | 2 | 33 |
| ZC3H7B | 2 | IR and W/A | 2 | 33 |
| ARHGAP27 | 2 | IR and W/A | 1;2 | 32 |
| MFHAS1 | 2 | IR and W/A | 1 | 32 |
| PRAG1 | 2 | IR and W/A | 1 | 32 |
| SUDS3 | 2 | IR and W/A | 2 | 28 |
| MSRA | 2 | IR and W/A | 2 | 27 |
| DEFB134 | 2 | IR and W/A | 1 | 24 |
| PRSS53 | 2 | IR and W/A | 2 | 23 |
| AREL1 | 2 | IR and W/A | 1 | 21 |
| RP11-196G11.6 | 2 | IR and W/A | 2 | 21 |

| | | | | |
|---|---|---|---|---|
| CTA-398F10.2 | 2 | IR and W/A | 1 | 19 |
| FDFT1 | 2 | IR and W/A | 1 | 18 |
| LTBP2 | 2 | IR and W/A | 2 | 18 |
| YLPM1 | 2 | IR and W/A | 2 | 18 |
| XKR6 | 2 | IR and W/A | 2 | 16 |
| EP300 | 2 | IR and W/A | 2 | 15 |
| L3MBTL2 | 2 | IR and W/A | 2 | 13 |
| ZNF668 | 2 | IR and W/A | 2 | 13 |
| FAM66A | 2 | IR and W/A | 1 | 12 |
| RP11-981G7.6 | 2 | IR and W/A | 1 | 12 |
| FCF1 | 2 | IR and W/A | 2 | 10 |

Table S5 Gene Information for Brain and Psychiatric-related Traits with a q-Value Threshold of 0.05. Detailed information related to AD is available in Supplementary Table S5.xlsx.

**Result Analysis**

Based on experimental results and related reports, multiple genes associated with Alzheimer's Disease (AD) and related emotional traits (such as anxiety and irritability) have been identified. Among these, genes such as **APOE**, **CLU**, **CD2AP**, **FBXL19**, **SETD1A**, and **NEGR1** exhibit significant overlap across different traits. These genes primarily participate in key biological processes including neuroinflammation, synaptic plasticity, lipid metabolism, and epigenetic regulation, revealing potential shared mechanisms between AD and emotional traits. For example, **APOE** significantly influences AD risk by regulating lipid metabolism and neuroinflammation, while **NEGR1** plays an important role in synaptic maintenance and neuroprotection.

These shared genes not only help deepen the understanding of the genetic foundations of AD and its related emotional traits but also provide potential biomarkers and therapeutic targets for early diagnosis, risk assessment, and precision treatment. Additionally, the research underscores the importance of genetic diversity, such as the differing risk effects of **TOMM40** across various populations, suggesting that future studies should consider racial and genetic background diversity to achieve more personalized intervention measures. Overall, these findings offer valuable insights and research directions for the prevention and treatment of AD and its associated neuropsychiatric disorders.

Table S6 Annotation of AD-Related Phenotypes in ROSMAP PrediXcan Analysis

<table>
<tr><th>Phenotypic</th><th>Description</th></tr>
<tr><td>gender</td><td>1 = Male<br>0 = Female</td></tr>
<tr><td>educ</td><td>The years of education variable is based on the number of years of regular school reported at baseline cognitive testing.</td></tr>
<tr><td>race</td><td>1：White<br>2：Black or African American<br>3：American Indian or Alaska Native</td></tr>
<tr><td>spanish</td><td>1 ： Yes<br>2 ： No</td></tr>
<tr><td>apoe_genotype</td><td><table>
<tr><td>value</td><td>coding</td></tr>
<tr><td>22</td><td>E2E2</td></tr>
<tr><td>23</td><td>E2E3</td></tr>
<tr><td>24</td><td>E2E4</td></tr>
<tr><td>33</td><td>E3E3</td></tr>
<tr><td>34</td><td>E3E4</td></tr>
<tr><td>44</td><td>E4E4</td></tr>
</table></td></tr>
<tr><td>age_at_visit_max</td><td>The age of the patient at the last follow-up.</td></tr>
<tr><td>age_first_ad_dx</td><td>The age at first Alzheimer's dementia diagnosis</td></tr>
<tr><td>age_death</td><td>Age of death is calculated from subtracting date of birth from date of death and dividing the difference by days</td></tr>
</table>

**Formatted Table**

<table>
<tr><td>cts_mmse30_first_ad_dx</td><td>MMSE (mini-mental state examination) score of patients with Alzheimer's disease at the first diagnosis.</td></tr>
<tr><td>cts_mmse30_lv</td><td>Cts_mmse30_lv: MMSE score of the patient at the last follow-up. The data is 26, with a full score of 30. The higher the score, the better the cognitive function.</td></tr>
<tr><td>pmi</td><td>Post-mortem interval (PMI) refers to the interval between death and tissue preservation in hours</td></tr>
<tr><td>braaksc</td><td>Braak Stage is a semiquantitative measure of severity of neurofibrillary tangle (NFT) pathology.
<table>
<tr><td>value</td><td>coding</td></tr>
<tr><td>0</td><td>0</td></tr>
<tr><td>1</td><td>I</td></tr>
<tr><td>2</td><td>II</td></tr>
<tr><td>3</td><td>III</td></tr>
</table></td></tr>
<tr><td>ceradsc</td><td>CERAD score is a semiquantitative measure of neuritic plaques.</td></tr>
<tr><td>cogdx</td><td>
<table>
<tr><td>value</td><td>coding</td><td>if using a binary variable, recommendation is</td></tr>
<tr><td>1</td><td>Definite</td><td>yes</td></tr>
<tr><td>2</td><td>Probable</td><td>yes</td></tr>
<tr><td>3</td><td>Possible</td><td>no</td></tr>
<tr><td>4</td><td>No AD</td><td>no</td></tr>
</table></td></tr>
</table>

<table>
<tr>
<td>dcfdx_lv</td>
<td>A clinical diagnosis of cognitive status is rendered at every assessment based on a three-stage process including computer scoring of cognitive tests, clinical judgment by a neuropsychologist, and diagnostic classification by a clinician.
<table>
<tr><td>Value</td><td>Coding</td></tr>
<tr><td>1</td><td>NCI: No cognitive impairment</td></tr>
<tr><td>2</td><td>MCI: Mild cognitive impairment, no other condition contributing to CI</td></tr>
<tr><td>3</td><td>MCI+: Mild cognitive impairment AND another condition contributing to CI</td></tr>
<tr><td>4</td><td>AD: Alzheimer’s dementia, no other condition contributing to CI (NINCDS/ADRDA Probable AD)</td></tr>
<tr><td>5</td><td>AD+: Alzheimer’s dementia AND other condition contributing to CI (NINCDS/ADRDA Possible AD)</td></tr>
<tr><td>6</td><td>Other dementia: Other primary cause of dementia, no clinical evidence of Alzheimer’s</td></tr>
</table>
</td>
</tr>
</table>

| | | | |
|---|---|---|---|
| | | | dementia |

Table S7 Gene Information Related to Phenotypes Predicted by the Individual-level PrediXcan Method. Detailed information is available in Supplementary Table S7.xlsx.

**Result Analysis**

Based on the analysis using the personalized PrediXcan method on the ROSMAP dataset, multiple genes have been identified as significantly associated with Alzheimer's Disease (AD) and its related phenotypes (such as cognitive impairment, mortality, MMSE scores, etc.). Among these, genes including **PPARGC1A**, **HERC1**, **WDR41**, **ATG5**, **EGFR**, **NPTXR**, **ACAT2**, **NPNT**, **RAB27B**, **ECD**, **MS4A6A**, **GPR55**, **ACACB**, **PTPRZ1**, **SMIM24**, **POT1**, **SYT2**, **CTNND1**, **CIART**, **NCALD**, and **CERS6** have been confirmed to play critical roles in the pathological processes of AD. These genes are involved in various biological processes, including neuroinflammation, synaptic plasticity, autophagy, lipid metabolism, protein degradation, cell cycle regulation, and neuroprotection, further revealing potential links between AD and these biological processes. For example, **PPARGC1A** demonstrates neuroprotective effects by improving AD-like behavioral abnormalities (such as deficits in spatial memory and working memory impairment), while **ATG5** and **EGFR** play important roles in autophagy and cell signaling, helping to alleviate neuroinflammation and promote cell survival.

Additionally, some genes in the study (such as **PPARGC1A**, **HERC1**, **WDR41**, **ATG5**, **EGFR**, etc.) have been directly reported in the literature to be associated with AD, reinforcing their value as potential biomarkers and therapeutic targets. These findings not only deepen our understanding of the genetic foundations of AD but also provide new directions for early diagnosis, risk assessment, and precision treatment. Furthermore, certain genes (such as **GPR55**, **SMIM24**, **CIART**, etc.) have not yet been sufficiently studied in the context of AD, indicating the need for future investigations into their specific roles in the pathogenesis of AD. Overall, these results offer valuable insights into the complex genetic network of AD and lay the groundwork for developing targeted intervention strategies.

Table S8 Enrichment analysis of phenotype-related genes in cognitive diagnosis

| GO-term | Description | Count in Network | Strength | Signal | False Discovery Rate |
|---|---|---|---|---|---|
| **Cellular Component (Gene Ontology)** | | | | | |
| GO:0005622 | Intracellular anatomical structure | 271 of 14891 | 0.07 | 0.43 | 9.90e-05 |
| GO:0005829 | Cytosol | 121 of 5438 | 0.15 | 0.36 | 0.0049 |
| GO:0043231 | Intracellular membrane-bounded organelle | 228 of 12149 | 0.08 | 0.35 | 0.0038 |
| GO:0043229 | Intracellular organelle | 241 of 13231 | 0.07 | 0.32 | 0.0072 |
| GO:0043227 | Membrane-bounded organelle | 239 of 13188 | 0.06 | 0.3 | 0.0114 |
| GO:0005737 | Cytoplasm | 223 of 12056 | 0.07 | 0.3 | 0.0114 |
| GO:0043226 | Organelle | 250 of 14017 | 0.06 | 0.3 | 0.012 |
| **Tissue Expression (TISSUES)** | | | | | |
| BTO:0000042 | Animal | 277 of 15148 | 0.07 | 0.47 | 5.36e-06 |
| BTO:0000000 | Tissues, cell types and enzyme sources | 278 of 15199 | 0.07 | 0.47 | 5.36e-06 |
| BTO:0000227 | Central nervous system | 128 of 5825 | 0.15 | 0.37 | 0.0036 |
| BTO:0000142 | Brain | 127 of 5733 | 0.15 | 0.37 | 0.0036 |
| BTO:0001484 | Nervous system | 129 of 6016 | 0.14 | 0.33 | 0.0094 |

Formatted Table

| | | | | | |
|---|---|---|---|---|---|
| BTO:0000282 | Head | 138 of 6642 | 0.12 | 0.3 | 0.0163 |
| BTO:0001488 | Endocrine gland | 132 of 6403 | 0.12 | 0.27 | 0.0351 |
| BTO:0000522 | Gland | 142 of 7004 | 0.11 | 0.26 | 0.0351 |
| BTO:0001489 | Whole body | 235 of 13099 | 0.06 | 0.25 | 0.0351 |
| **Subcellular Localization (COMPARTMENTS)** | | | | | |
| GOCC:0005622 | Intracellular | 221 of 11512 | 0.09 | 0.36 | 0.0024 |
| GOCC:0005737 | Cytoplasm | 165 of 8195 | 0.11 | 0.29 | 0.0203 |
| **Annotated Keywords (UniProt)** | | | | | |
| KW-0597 | Phosphoprotein | 163 of 8122 | 0.11 | 0.3 | 0.0178 |
| KW-0025 | Alternative splicing | 196 of 10313 | 0.08 | 0.29 | 0.0178 |
| KW-0007 | Acetylation | 78 of 3362 | 0.17 | 0.27 | 0.041 |

Table S9 Enrichment of Hub Genes Associated with Cognitive Diagnostic Phenotypes Predicted by the AD-MIF Model

| GO-term | Description | Count in Network | Strength | Signal | False Discovery Rate |
|---|---|---|---|---|---|
| **Cellular Component (Gene Ontology)** | | | | | |
| GO:0005622 | Intracellular anatomical structure | 240 | 0.06 | 0.26 | 0.0289 |
| GO:0005654 | Nucleoplasm | 89 | 0.18 | 0.29 | 0.0289 |
| GO:0031981 | Nuclear lumen | 93 | 0.16 | 0.27 | 0.0402 |
| **Tissue Expression (TISSUES)** | | | | | |
| BTO:0000042 | Animal | 251 | 0.07 | 0.44 | 3.92e-05 |
| BTO:0001489 | Whole body | 221 | 0.08 | 0.36 | 0.0019 |
| BTO:0000142 | Brain | 116 | 0.15 | 0.37 | 0.0042 |
| BTO:0001484 | Nervous system | 118 | 0.14 | 0.33 | 0.0101 |

Table S10 Top Repositioning Hits for Father's Alzheimer's Disease/Dementia (Ranked by Q-values and P-values)

| Drug | Tissue | Rank | P-value | Q-value | Z-score | Brief description |
|---|---|---|---|---|---|---|
| Ibudilast | Adrenal Gland | 1 | 2.23E-06 | 0.232 | 1.748 | No psychiatric relevance. Attenuated astroglial reactivity and inflammation in a PD mouse model[1]. |
| ORG-9768 | Adrenal Gland | 2 | 4.47E-06 | 0.232 | 1.748 | No relevant psychiatric or AD-related findings. |
| Dutasteride | Adrenal Gland | 3 | 6.70E-06 | 0.232 | 1.748 | Potential treatment for PMDD[2]. |
| LY-2228820 | Adrenal Gland | 4 | 8.94E-06 | 0.232 | 1.748 | No psychiatric relevance. p38 MAPK inhibitor developed for cancer therapy[3]. |
| Talazoparib | Adrenal Gland | 5 | 1.12E-05 | 0.232 | 1.748 | No psychiatric relevance. Oral PARP inhibitor with minimal non-hematologic toxicity[4]. |
| TPCA-1 | Adrenal Gland | 6 | 1.34E-05 | 0.232 | 1.748 | No psychiatric relevance. Suppresses inflammatory gene expression under cytokine stimuli[5]. |
| XMD11-85H | Adrenal Gland | 7 | 1.56E-05 | 0.232 | 1.748 | No relevant psychiatric or AD-related findings. |
| LY-2090314 | Adrenal Gland | 8 | 1.79E-05 | 0.232 | 1.748 | No psychiatric |

| | | | | | | |
|---|---|---|---|---|---|---|
| | | | | | | relevance. May exhibit antiviral activity through a GSK-3-independent mechanism[6]. |
| STA-5326 | Adrenal Gland | 9 | 2.01E-05 | 0.232 | 1.748 | No psychiatric relevance. Well-tolerated in Crohn's disease patients at 28 mg and above[7]. |
| Ambroxol | Adrenal Gland | 10 | 2.23E-05 | 0.232 | 1.748 | No psychiatric relevance. Exhibits antioxidant and anti-inflammatory properties[8]. |
| Tranylcypromine | Adrenal Gland | 11 | 2.46E-05 | 0.232 | 1.748 | Clinically used for depression[9]. |
| RG-108 | Adrenal Gland | 12 | 2.68E-05 | 0.232 | 1.748 | No psychiatric relevance. Attenuates cisplatin-induced cochlear apoptosis[10]. |
| Avanafil | Adrenal Gland | 13 | 2.90E-05 | 0.232 | 1.748 | No psychiatric relevance. A PDE-5 inhibitor for erectile dysfunction[11]. |
| Ethinyl-Estradiol | Adrenal Gland | 14 | 3.13E-05 | 0.232 | 1.748 | Contains thrombogenic risks and may adversely affect mood[12]. |
| Ticlopidine | Adrenal Gland | 15 | 3.35E-05 | 0.232 | 1.748 | No psychiatric relevance. Antithrombotic |

| | | | | | | |
|---|---|---|---|---|---|---|
| | | | | | | agent[13]. |
| Losartan | Adrenal Gland | 16 | 3.58E-05 | 0.232 | 1.748 | Exhibits antidepressant effects in rodent models[14]. |
| Colforsin-Daproate | Adrenal Gland | 17 | 3.80E-05 | 0.232 | 1.748 | No psychiatric relevance. Improved heart failure symptoms without serious adverse events[15]. |
| D-Serine | Adrenal Gland | 18 | 4.02E-05 | 0.232 | 1.748 | Restores excitation/inhibition balance and improves cognitive function[16]. |
| Dacomitinib | Adrenal Gland | 19 | 4.25E-05 | 0.232 | 1.748 | Shows promising efficacy in glioblastoma studies[17]. |
| CYT387 | Adrenal Gland | 20 | 4.47E-05 | 0.232 | 1.748 | No psychiatric relevance. Inhibits cell proliferation and autophagy[18]. |

Table S11 Top Repositioning Hits for Siblings 's Alzheimer's Disease/Dementia (Ranked by Q-values and P-values)

| Drug | Tissue | Rank | P-value | Q-value | Z-score | Brief description |
|---|---|---|---|---|---|---|
| AS-605240 | Stomach | 1 | 1.02E-06 | 0.104 | 1.734 | PI3Kγ inhibitor improves insulin resistance in obese diabetic models[19]. |
| Phenylacetylglutamine | Stomach | 2 | 2.05E-06 | 0.104 | 1.734 | Potential psychiatric relevance via the gut-brain axis. Linked to neurological and systemic diseases[20]. |
| BMS-777607 | Stomach | 3 | 3.07E-06 | 0.104 | 1.734 | No psychiatric relevance. Combined with anti–PD-1, decreases tumor growth[21]. |
| Bortezomib | Stomach | 4 | 4.10E-06 | 0.104 | 1.734 | Promising therapy for severe cNPSLE with psychosis[22]. |
| AZD-4547 | Stomach | 5 | 5.12E-06 | 0.104 | 1.734 | FGFR inhibitor, limited findings in psychiatric relevance. |
| Senicapoc | Stomach | 6 | 6.15E-06 | 0.104 | 1.734 | No psychiatric relevance. Slows mammary tumor growth[23]. |
| AZD-1480 | Stomach | 7 | 7.17E-06 | 0.104 | 1.734 | No psychiatric relevance. Regulates proliferation and immunity in HL[24]. |
| Oxybuprocaine | Stomach | 8 | 8.19E-06 | 0.104 | 1.734 | Used as an anesthetic, no psychiatric relevance. |
| Linagliptin | Stomach | 9 | 9.22E-06 | 0.104 | 1.734 | No psychiatric relevance. A DPP-4 inhibitor primarily used for the treatment of type 2 |

| | | | | | | |
|---|---|---|---|---|---|---|
| | | | | | | diabetes[25]. |
| LCQ-908 | Stomach | 10 | 1.02E-05 | 0.104 | 1.734 | No psychiatric relevance, DGAT-1 inhibitor. |
| HG-14-10-04 | Stomach | 11 | 1.13E-05 | 0.104 | 1.734 | No psychiatric relevance. |
| Zofenopril-Calcium | Stomach | 12 | 1.23E-05 | 0.104 | 1.734 | ACE inhibitor, no psychiatric relevance. |
| Zileuton | Stomach | 13 | 1.33E-05 | 0.104 | 1.734 | No psychiatric relevance. LOX inhibitor with anti-ferroptotic and anti-inflammatory activity[26]. |
| OSI-027 | Stomach | 14 | 1.43E-05 | 0.104 | 1.734 | No psychiatric relevance, potential sex-specific HNSC drug[27]. |
| Mazindol | Stomach | 15 | 1.54E-05 | 0.104 | 1.734 | Dopamine and norepinephrine reuptake inhibitor for ADHD[28]. |
| QL-X-138 | Stomach | 16 | 1.64E-05 | 0.104 | 1.734 | No psychiatric relevance. Potent antiproliferative agent for lymphoma and leukemia[29]. |
| AS-601245 | Stomach | 17 | 1.74E-05 | 0.104 | 1.734 | No psychiatric relevance, still in development. |
| Pirfenidone | Stomach | 18 | 1.84E-05 | 0.104 | 1.734 | No psychiatric relevance. Potential adjunct to immunotherapy for tumor-related pulmonary fibrosis[30]. |
| KIN001-269 | Stomach | 19 | 1.95E-05 | 0.104 | 1.734 | No psychiatric relevance. |
| Meloxicam | Stomach | 20 | 2.05E-05 | 0.104 | 1.734 | Adjuvant therapy, improves cognitive function in |

schizophrenia patients[31].

Table S12 Top Repositioning Hits for Irritability (Ranked by Q-values and P-values)

| Drug | Tissue | Rank | P-value | Q-value | Z-score | Brief description |
|---|---|---|---|---|---|---|
| PD-153035 | Brain Frontal Cortex BA9 | 1 | 4.66E-07 | 0.049 | -1.741 | Improves prepulse inhibition (PPI) deficits in animal models [32, 33]. |
| LY-2874455 | Brain Cerebellum | 1 | 5.59E-07 | 0.060 | -1.732 | No relevant findings. |
| Belinostat | Uterus | 1 | 6.52E-07 | 0.068 | -1.739 | Shows antidepressant effects in animal models but mixed efficacy in later studies[34]. |
| LCL-161 | Skin Not Sun Exposed Suprapubic | 1 | 7.45E-07 | 0.079 | -1.738 | No relevant findings. |
| LCQ-908 | Cells EBV-transformed lymphocytes | 1 | 8.38E-07 | 0.084 | -1.769 | No relevant findings. |
| Pranlukast | Cells Cultured fibroblasts | 1 | 1.12E-06 | 0.115 | -1.746 | Long-term use associated with increased depression risk [35]. |
| Brivudine | Muscle Skeletal | 1 | 1.30E-06 | 0.126 | -1.759 | No relevant findings. |
| AZ-20 | Muscle Skeletal | 2 | 2.51E-06 | 0.126 | -1.758 | No relevant findings. |
| Penfluridol | Lung | 1 | 1.30E-06 | 0.135 | -1.741 | Effective for schizophrenia, well-studied over decades [36]. |
| A-769662 | Brain Substantia nigra | 1 | 2.23E-06 | 0.148 | -1.735 | Limited efficacy in psychiatric disorders, used as a control in studies. |
| Simeprevir | Brain Substantia nigra | 2 | 2.79E-06 | 0.148 | -1.735 | No psychiatric relevance, some safety studies. |
| Ambrisentan | Cells Cultured | 2 | 2.89E-06 | 0.148 | -1.746 | Shows potential in protecting |

| | | | | | | |
|---|---|---|---|---|---|---|
| | fibroblasts | | | | | blood-brain barrier post-injury. |
| I-BET-762 | Brain Anterior cingulate cortex BA24 | 1 | 1.68E-06 | 0.156 | -1.737 | No relevant findings. |
| Budipine | Brain Anterior cingulate cortex BA24 | 2 | 2.98E-06 | 0.156 | -1.737 | Effective for tremor in Parkinson's, less for other symptoms [37]. |
| Zaldaride | Adipose Subcutaneous | 1 | 1.58E-06 | 0.164 | -1.749 | No relevant findings. |
| UNC-1215 | Stomach | 1 | 1.68E-06 | 0.18 | -1.723 | No relevant findings. |
| Etanidazole | Brain Nucleus accumbens basal ganglia | 1 | 2.98E-06 | 0.227 | -1.737 | No relevant findings. |
| BTS-54505 | Brain Nucleus accumbens basal ganglia | 2 | 4.28E-06 | 0.227 | -1.737 | No relevant findings. |
| VX-661 | Lung | 2 | 4.56E-06 | 0.236 | -1.741 | Studies focus on cystic fibrosis, some psychiatric safety data available. |
| XMD-16144 | Adipose Visceral Omentum | 1 | 6.24E-06 | 0.245 | -1.751 | No relevant findings. |

Table S13 Top Repositioning Hits for Worrier/anxious feelings (Ranked by Q-values and P-values)

| Drug | Tissue | Rank | P-value | Q-value | Z-score | Brief description |
|---|---|---|---|---|---|---|
| NVP-TAE226 | Brain Putamen basal ganglia | 1 | 3.44E-06 | 0.16 | -1.720 | No relevant findings. |
| Pinitol | Brain Putamen basal ganglia | 2 | 3.44E-06 | 0.16 | -1.720 | Shows potential for treating cognitive impairment and sensorimotor gating deficits in schizophrenia [38]. |
| Rimexolone | Brain Putamen basal ganglia | 3 | 4.47E-06 | 0.16 | -1.720 | No relevant findings. |
| PF-573228 | Heart Left Ventricle | 1 | 1.58E-06 | 0.17 | -1.734 | No relevant findings. |
| AS-703026 | Prostate | 1 | 1.68E-06 | 0.18 | -1.718 | No relevant findings. |
| AS-703026 | Lung | 1 | 1.96E-06 | 0.21 | -1.721 | No relevant findings. |
| BX-795 | Brain Cerebellum | 1 | 2.05E-06 | 0.217 | -1.742 | Promising neuroprotective effects, potential treatment for Parkinson's disease and other protein conformational disorders [39]. |
| Balaglitazone | Brain Caudate basal ganglia | 1 | 2.05E-06 | 0.220 | -1.734 | No relevant findings. |
| Avanafil | Heart Atrial Appendage | 1 | 2.23E-06 | 0.231 | -1.752 | Primarily used for erectile dysfunction, rapid onset of action observed [40]. |
| AS-703026 | Breast Mammary Tissue | 1 | 2.23E-06 | 0.24 | -1.714 | No relevant findings. |

| | | | | | | |
|---|---|---|---|---|---|---|
| Pazopanib | Heart Left Ventricle | 2 | 6.89E-06 | 0.247 | -1.734 | No relevant findings. |
| NVP-BHG712 | Heart Left Ventricle | 3 | 6.89E-06 | 0.247 | -1.734 | No relevant findings. |
| Pelitinib | Skin Not Sun Exposed Suprapubic | 1 | 2.51E-06 | 0.27 | -1.726 | No relevant findings. |
| XMD-885 | Brain Hippocampus | 1 | 6.61E-06 | 0.328 | -1.727 | Identified via statistical analysis, similar to current study methods [41]. |
| SB-939 | Brain Hippocampus | 2 | 6.61E-06 | 0.328 | -1.727 | No relevant findings. |
| Barasertib-HQPA | Brain Hippocampus | 3 | 1.22E-05 | 0.328 | -1.727 | No relevant findings. |
| SB-939 | Brain Hippocampus | 4 | 1.22E-05 | 0.328 | -1.727 | No relevant findings. |
| Resveratrol | Brain Hippocampus | 5 | 1.72E-05 | 0.37 | -1.727 | Known for its antioxidant and neuroprotective properties, improves mood and cognition [42]. |
| Mitoxantrone | Adipose_Visceral_Omentum | 1 | 6.98E-06 | 0.374 | -1.730 | Shows benefit in CNS treatment; reduces glutamate production in neurodegenerative diseases [43, 44]. |
| Daunorubicin | Adipose Visceral Omentum | 2 | 6.98E-06 | 0.374 | -1.730 | Ongoing studies are validating its potential for repurposing [45]. |

Table S14 Drug enrichment analyses P values with drug-sets defined by MEDI-HPS

| Disorder | Self (Fisher) | Compet (Fisher) | Self (minP) | Compet (minP) |
|---|---|---|---|---|
| **Enrichment for indicated drugs of each disorder** | | | | |
| Irritability | 0 | 0.373 | 3.26E-62 | 0.576 |
| Worrier / anxious feelings | 0 | 0.490 | 1.58E-59 | 0.499 |
| Illnesses of father: Alzheimer's disease/dementia | 0 | 0.830 | 5.67E-79 | 0.788 |
| Illnesses of mother: Alzheimer's disease/dementia | 0 | 0.939 | 4.37E-82 | 0.843 |
| Illnesses of siblings: Alzheimer's disease/dementia | 4.13E-237 | 0.438 | 2.28E-75 | 0.359 |
| **Enrichment for drugs for depression or anxiety** | | | | |
| Irritability | 0 | 5.14E-2 | 1.22E-39 | 0.527 |
| Worrier / anxious feelings | 0 | 0.686 | 2.91E-36 | 0.572 |
| Illnesses of father: Alzheimer's disease/dementia | 0 | 0.770 | 3.09E-49 | 0.391 |
| Illnesses of mother: Alzheimer's disease/dementia | 0 | 0.841 | 2.25E-53 | 0.700 |
| Illnesses of siblings: Alzheimer's disease/dementia | 2.84E-144 | 0.556 | 4.12E-45 | 0.301 |
| **Enrichment for all psychiatric drug** | | | | |
| Irritability | 0 | 0.769 | 2.98E-149 | 0.948 |
| Worrier / anxious feelings | 0 | 0.990 | 1.46E-47 | 0.477 |
| Illnesses of father: Alzheimer's disease/dementia | 0 | 0.782 | 1.14E-63 | 0.697 |
| Illnesses of mother: Alzheimer's disease/dementia | 0 | 0.957 | 8.49E-66 | 0.879 |
| Illnesses of siblings: Alzheimer's disease/dementia | 1.01E-186 | 0.770 | 2.60E-60 | 0.752 |

Table S15 Drug enrichment analyses P values with drug-sets defined by ClinicalTrials

| Disorder | Self (Fisher) | Compet (Fisher) | Self (minP) | Compet (minP) |
|---|---|---|---|---|
| **Enrichment for indicated drugs of Irritability** | | | | |
| Irritability | 0 | 0.589 | 8.15E-158 | 0.446 |
| Worrier | 1.00 | 1.78E-123 | 1.00 | 1.00 |
| AD | 0 | 0.631 | 2.81E-125 | 0.836 |

### Supplementary Figures

#### S1: Standardized Gene Count Results for Brain and Psychiatric Traits: 18 Remaining After Screening from an Initial 61.

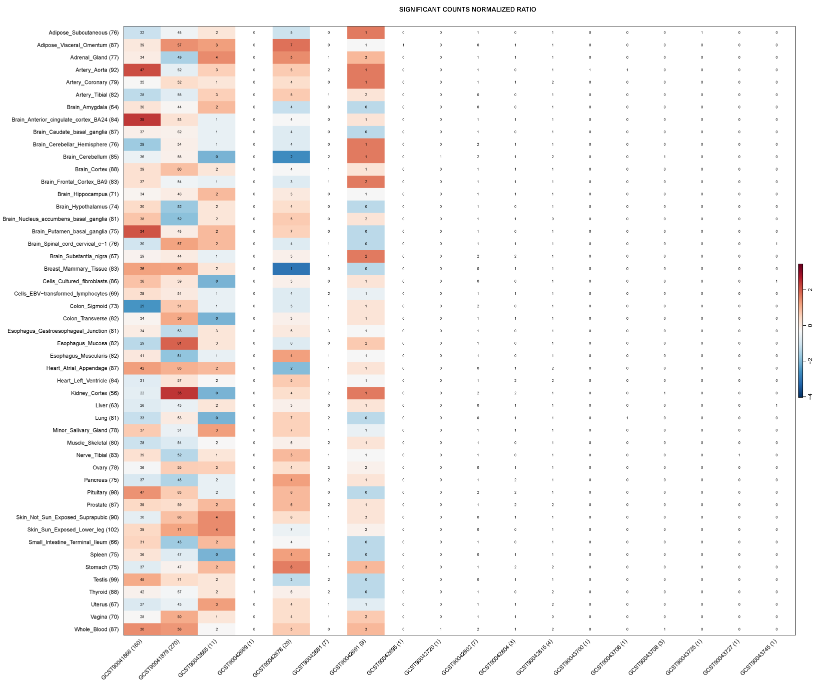

**Supplementary Figure S1: Standardized Gene Count Results for Brain and Psychiatric Traits: 18 Remaining After Screening from an Initial 61.**The heatmap displays the standardized gene counts for 18 brain and psychiatric traits, identified through S-PrediXcan analysis, across 49 different tissues. Red indicates a higher count of significant genes, while blue indicates a lower count, reflecting the pleiotropy of genes across various tissues. Brain regions such as the hippocampus (Brain_Hippocampus) and frontal cortex (Brain_Frontal_Cortex_BA9) show significant associations with AD-related traits, while non-brain tissues like the liver (Liver), lung (Lung), and aorta (Artery_Aorta) also exhibit notable gene associations. The data have been q-value corrected, with an FDR threshold of 0.01 applied.

S2: Distribution of Significant Genes for Five Brain and Psychiatric Traits (Including AD Family History and Emotional Traits) Across 49 Tissues Identified via S-PrediXcan Analysis.

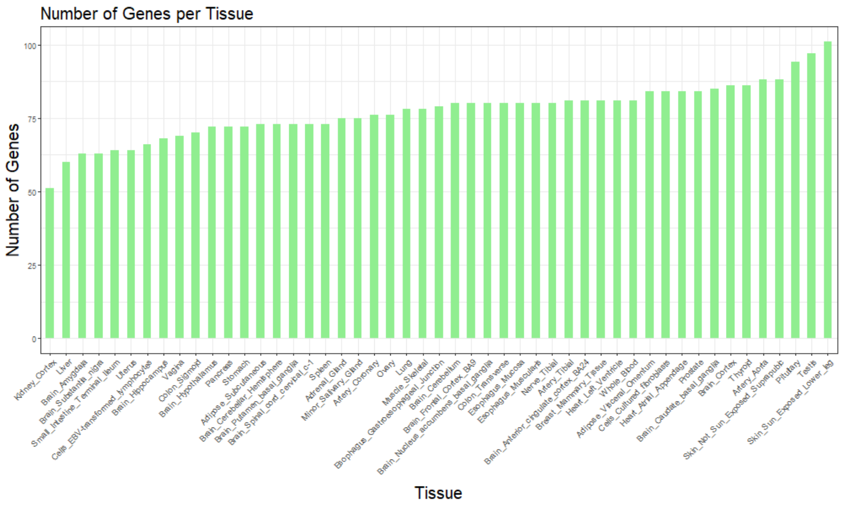

**Supplementary Figure S2: Distribution of Significant Genes for Five Brain and Psychiatric Traits (Including AD Family History and Emotional Traits) Across 49 Tissues Identified via S-PrediXcan Analysis.** This figure illustrates the distribution of the number of significant genes for five key traits across 49 different tissues. The horizontal axis represents the tissue names, and the vertical axis shows the number of significant genes. It can be observed that certain tissues, such as the Testis and Spleen, have a higher number of significant genes, suggesting that these tissues may play a potential role in Alzheimer's disease (AD)-related characteristics. Brain-related tissues, such as the Hippocampus (Brain_Hippocampus) and the Frontal Cortex (Brain_Frontal_Cortex_BA9), demonstrate significant gene enrichment in AD-related traits, further supporting the connection between these brain regions and AD pathology.

S3: Gene Expression Network for Brain- and Psychiatric-Related Traits at a q-value Threshold of 0.5.

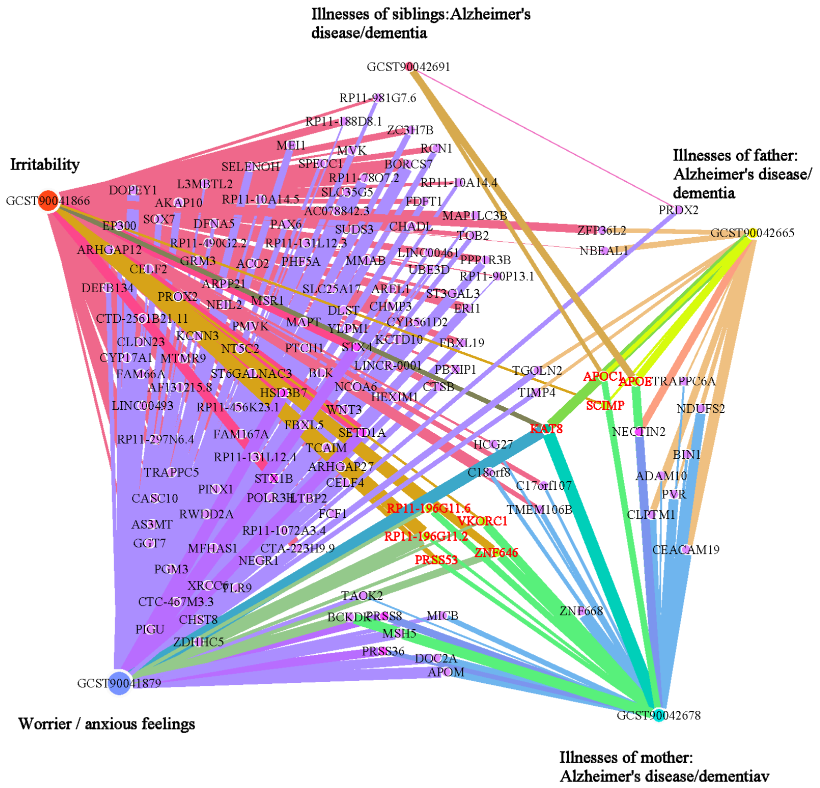

**Supplementary Figure S3. Gene Expression Network for Brain- and Psychiatric-Related Traits at a q-value Threshold of 0.5.** This figure displays the gene expression association networks for five significantly related traits at a q-value (pva.qval) threshold of 0.5. This threshold filters a larger number of trait-associated genes, and the figure visualizes the results of shared genes, highlighting the potential roles of these genes in regulating complex polygenic mechanisms.

S4: Gene association network of five significant traits with q threshold of 0.5.

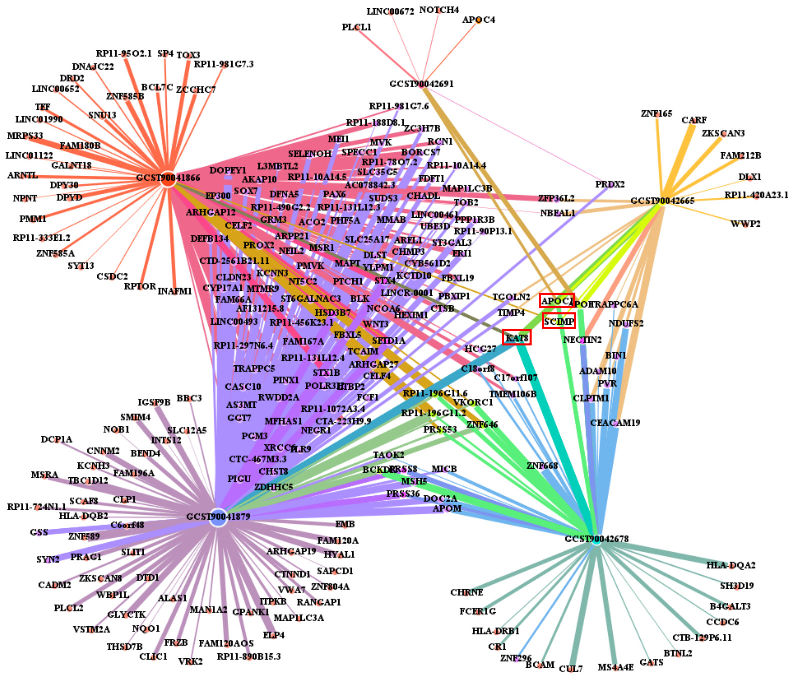

**Supplementary Figure S4: Gene Association Network of Five Significant Traits with Q Threshold of 0.5.** The figure shows the gene expression association network for five significant traits at a q-value (pva.qval) threshold of 0.5. This threshold filters out a larger number of genes associated with the traits. The gene expression in the red box is related to four traits.

S5: Gene Expression Network for Brain- and Psychiatric-Related Traits at a q-value Threshold of 0.1.

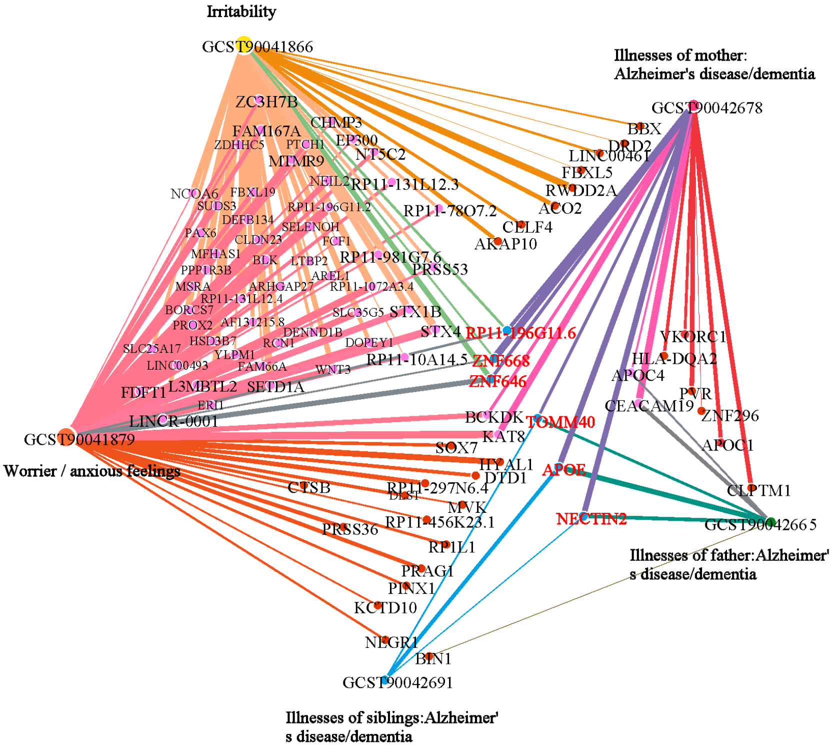

**Supplementary Figure S5: Gene Expression Network for Brain- and Psychiatric-Related Traits at a q-value Threshold of 0.1.** This figure illustrates that at a q-value (pva.qval) threshold of 0.1, the gene expression association network becomes more concentrated. The selected genes show a stronger association with AD-related traits, indicating their central role in the genetic mechanisms of the disease.

S6: Shared Gene Network between Alzheimer's Disease-Related Phenotypes.

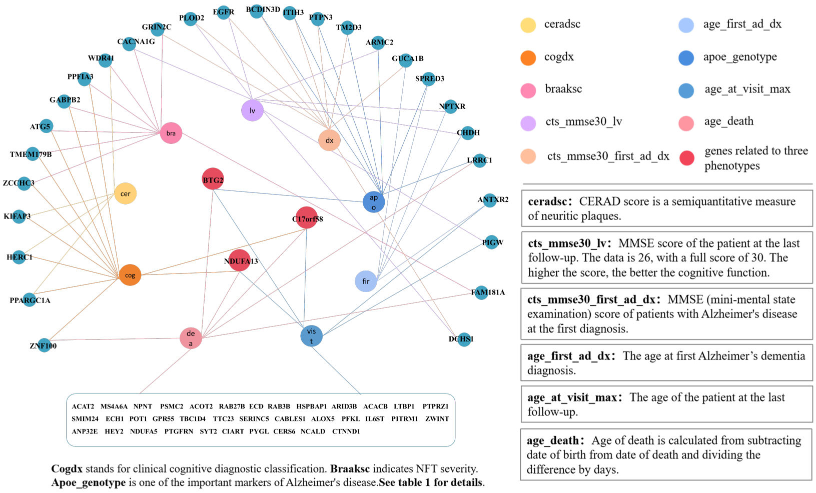

**Cogdx** stands for clinical cognitive diagnostic classification. **Braaksc** indicates NFT severity. **Apoe_genotype** is one of the important markers of Alzheimer's disease.**See table 1 for details**.

**Supplementary Figure S6: Shared Gene Network between Alzheimer's Disease-Related Phenotypes.** These phenotypes include age at first AD diagnosis (age_first_ad_dx), age at death (age_death), MMSE score at first AD diagnosis (cts_mmse30_first_ad_dx), MMSE score at last follow-up (cts_mmse30_lv), Braak pathology stage (braaksc), CERAD score (ceradsc), AD cognitive diagnosis (cogdx) dx, APOE genotype (apoe_genotype), and age at last follow-up (age_at_visit_max). Due to the significant genetic overlap between age at last follow-up and age at death, we only visualized the genes among these that have been proven to be related to brain research.

S7: Cellular Component (Gene Ontology) Enrichment Analysis of Genes Associated with Cognitive Diagnosis (similarity ⩾ 0.8).

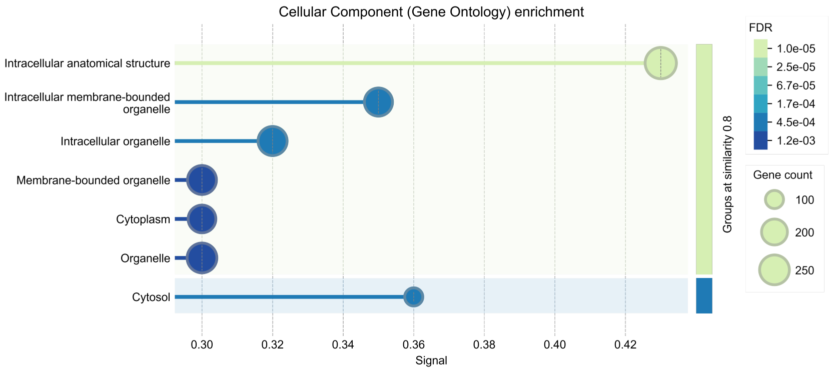

**Supplementary Figure S7: Cellular Component (Gene Ontology) Enrichment Analysis of Genes Associated with Cognitive Diagnosis (similarity ⩾ 0.8).**

S8: Cellular Component (Gene Ontology) Enrichment Analysis of Hub Genes Associated with Cognitive Diagnostic Phenotypes Predicted by the AD-MIF Model (similarity ⩾ 0.8).

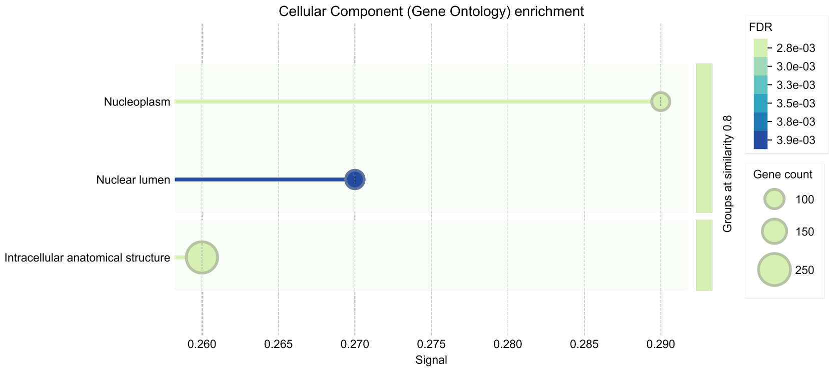

**Supplementary Figure S8: Cellular Component (Gene Ontology) Enrichment Analysis of Hub Genes Associated with Cognitive Diagnostic Phenotypes Predicted by the AD-MIF Model (similarity ⩾ 0.8).**

S9: Intersection Analysis of Common Drugs and Tissues Across AD Family History Traits: Siblings, Father and Mother.

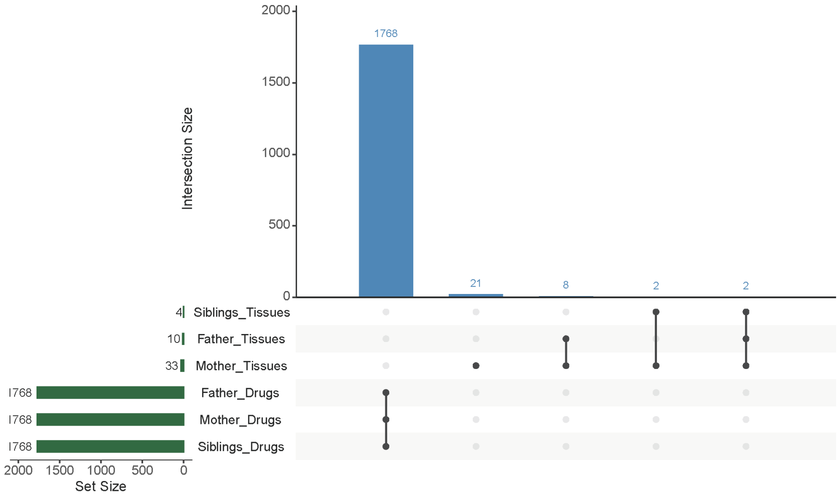

**Supplementary Figure S9: Intersection Analysis of Common Drugs and Tissues Across AD Family History Traits: Siblings, Father and Mother.**

S10: Top 50 Repurposed Drugs for Illnesses of Father - Alzheimer's Disease/Dementia Based on Q-value and P-value in Adrenal Gland.

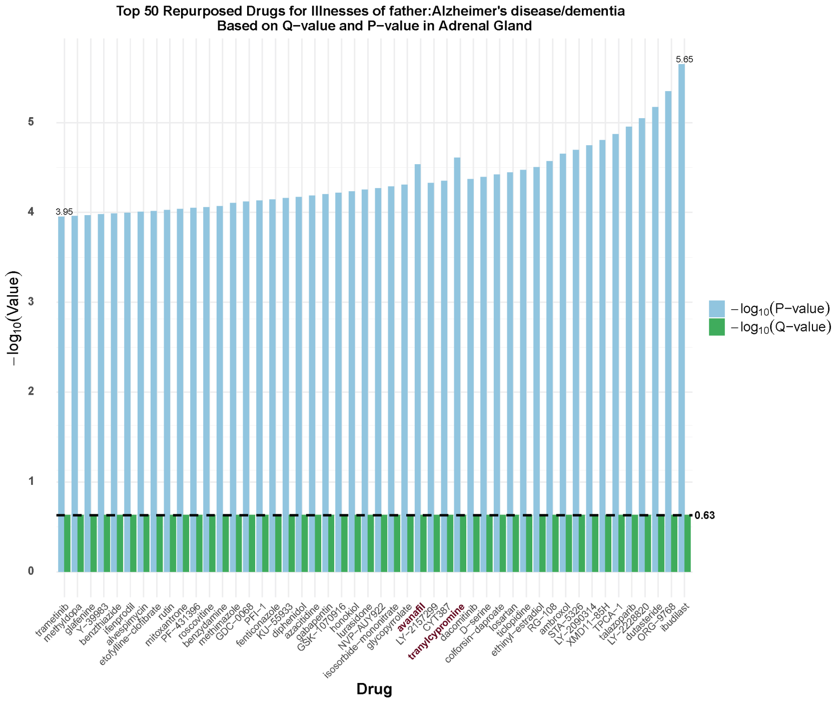

**Supplementary Figure S10: Top 50 Repurposed Drugs for Illnesses of Father - Alzheimer's Disease/Dementia Based on Q-value and P-value in Adrenal Gland.** This figure displays the top 50 potential repurposed drugs identified in adrenal gland tissues from individuals with a paternal history of AD/dementia. The visualization of drug effects is based on q-values and p-values, and duplicate drugs are highlighted using bold red font.

#### S11: Top 50 Repurposed Drugs for Illnesses of Siblings - Alzheimer's Disease/Dementia Based on Q-value and P-value in Stomach.

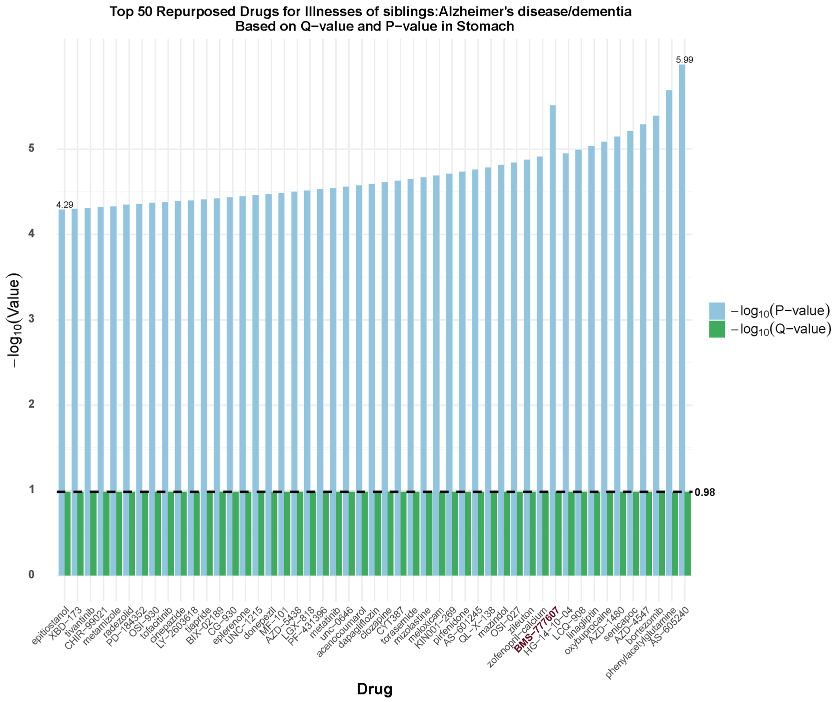

**Supplementary Figure S11:Top 50 Repurposed Drugs for Illnesses of Siblings - Alzheimer's Disease/Dementia Based on Q-value and P-value in Stomach.** This figure displays the top 50 potential repurposed drugs identified in stomach tissues from individuals with a sibling history of AD/dementia. The visualization of drug effects is based on q-values and p-values, and duplicate drugs are highlighted using bold red font.

S12: Top 50 Repurposed Drugs for Irritability Based on Q-value in Significant Tissues.

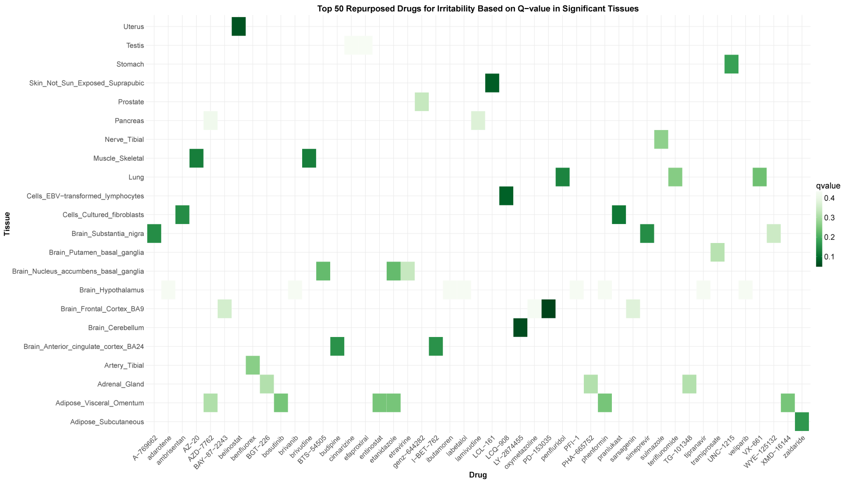

**Supplementary Figure S12: Top 50 Repurposed Drugs for Irritability Based on Q-value in Significant Tissues.** The analysis shows that some drugs display higher significance in specific tissues, particularly in brain and adipose-related regions. For instance, BX-795 shows high significance in brain regions such as the "Brain_Cerebellum" and "Brain_Putamen_basal_ganglia," suggesting its potential therapeutic role in these neurofunctional areas. Meanwhile, Avanafil exhibits notable significance in "Adipose_Subcutaneous," hinting at a potential link to fat metabolism or inflammatory responses. Pantoprazole shows significance across multiple tissues, indicating potential cross-system biological effects.Overall, the significance of these drugs in brain and adipose tissues suggests their possible role in regulating irritability by modulating the function of these tissues, providing new directions for drug repurposing research. The high significance in brain-related tissues, in particular, points to the nervous system as a key target for regulating irritability.

### S13: Top 50 Repurposed Drugs for Worrier/anxious feelings Based on Q-value in Significant Tissues

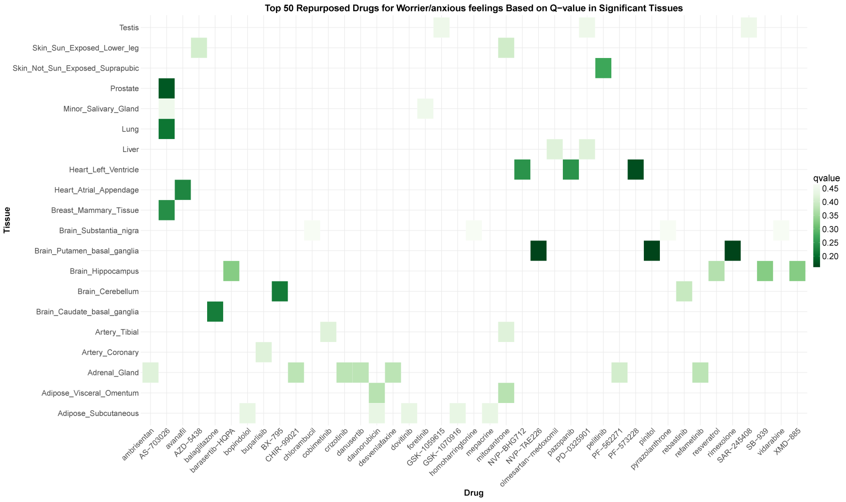

**Supplementary Figure S13: Top 50 Repurposed Drugs for Worrier/anxious feelings Based on Q-value in Significant Tissues.** This chart presents the top 50 drugs associated with Worrier/Anxious feelings across different tissues based on their significance. Balaglitazone and Mitoxantrone show high significance in multiple tissues, such as the Brain_Putamen_basal_ganglia, suggesting their potential role in regulating neurological functions. Brain-related regions, including the Brain_Cerebellum and Brain_Putamen_basal_ganglia, appear significantly across several drugs, indicating their critical role in the regulation of anxiety. Additionally, some drugs show high significance in the heart (Heart_Atrial_Appendage) and liver (Liver), suggesting that anxiety symptoms may be linked to cardiovascular and metabolic functions. Drugs like Balaglitazone and Mitoxantrone demonstrate broad activity across various tissues, while drugs like XMD-885 show more localized effects in specific tissues. Overall, these findings highlight the brain, heart, and liver as key targets for anxiety symptoms, offering valuable insights for potential drug repurposing strategies.

S14: Father's Alzheimer's Disease: Trait-Tissue-Gene-Drug Interaction Network.

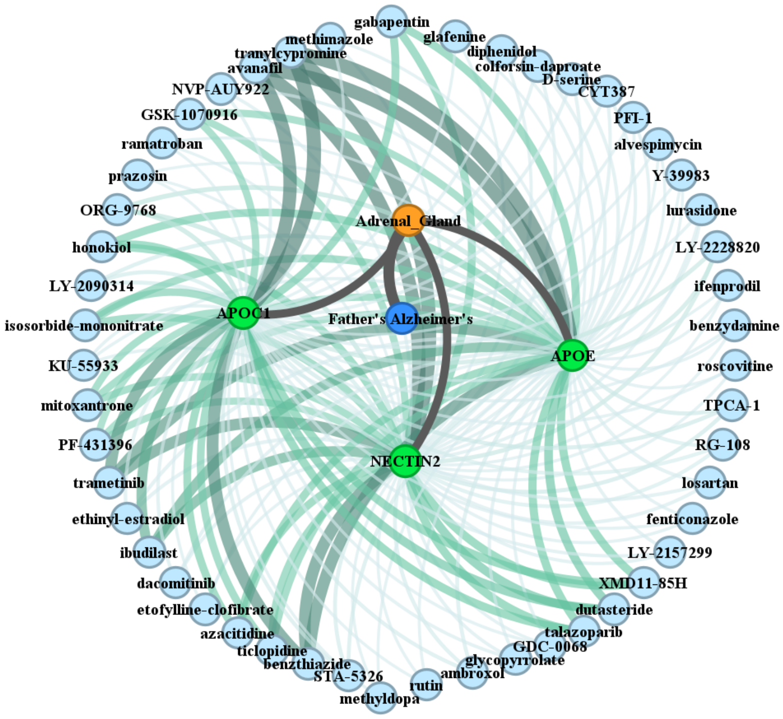

**Supplementary Figure S14: Father's Alzheimer's Disease: Trait-Tissue-Gene-Drug Interaction Network.** This network visualizes the interactions between Father's Alzheimer's Disease, the Adrenal Gland tissue, key genes, and potential repurposed drugs. The key genes involved are APOE, APOC1, and NECTIN2, all associated with Alzheimer's disease. Nodes represent the trait, tissue, genes, and drugs, while edges highlight the associations between them. Drug-Gene interactions are weighted by $-log_{10}(p)$, reflecting the strength of these interactions, and Tissue-Gene interactions have a fixed weight of 8, representing established links between the adrenal gland and the genes. This network provides insight into potential drug repurposing strategies for Alzheimer's disease.

S15: Network of Siblings' Alzheimer's Disease with Tissue-Gene-Drug Interactions for Drug Repurposing.

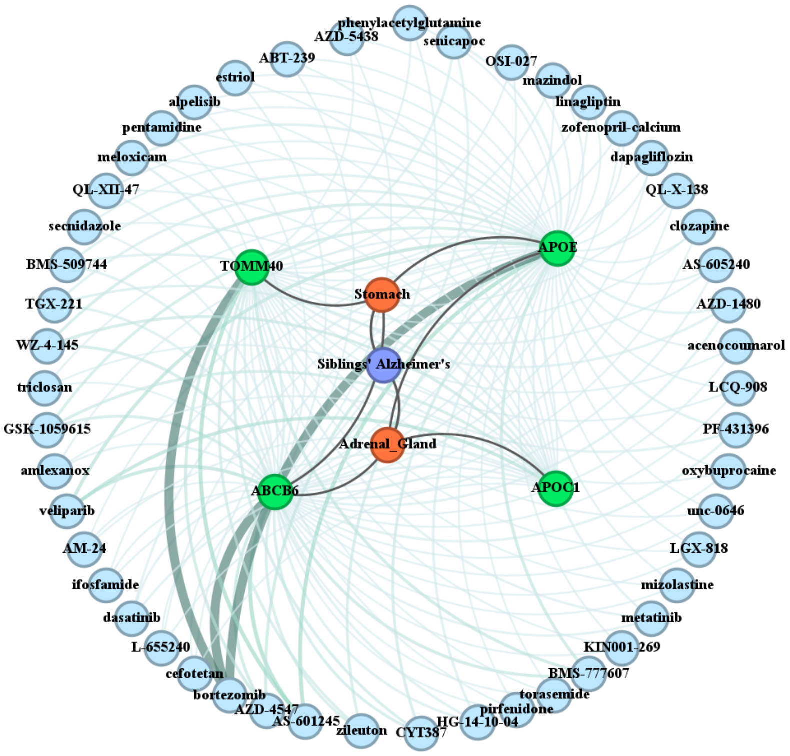

**Supplementary Figure S15: Network of Siblings' Alzheimer's Disease with Tissue-Gene-Drug Interactions for Drug Repurposing.** This network visualizes the interactions between Siblings' Alzheimer's Disease, the relevant tissues (stomach and adrenal gland), key genes, and potential repurposed drugs. The genes involved include TOMM40, APOE, APOC1, and ABCB6, all of which are linked to Alzheimer's disease. Nodes represent the trait, tissues, genes, and drugs, while edges indicate the associations between them. Drug-Gene interactions are weighted by $-log_{10}(p)$, reflecting the strength of the associations, while Tissue-Gene interactions are given a fixed weight of 8, representing known connections between the tissues and these genes. This network provides valuable insights for exploring drug repurposing strategies for Alzheimer's disease

S16: Irritability: Trait-Tissue-Gene-Drug Interaction Network.

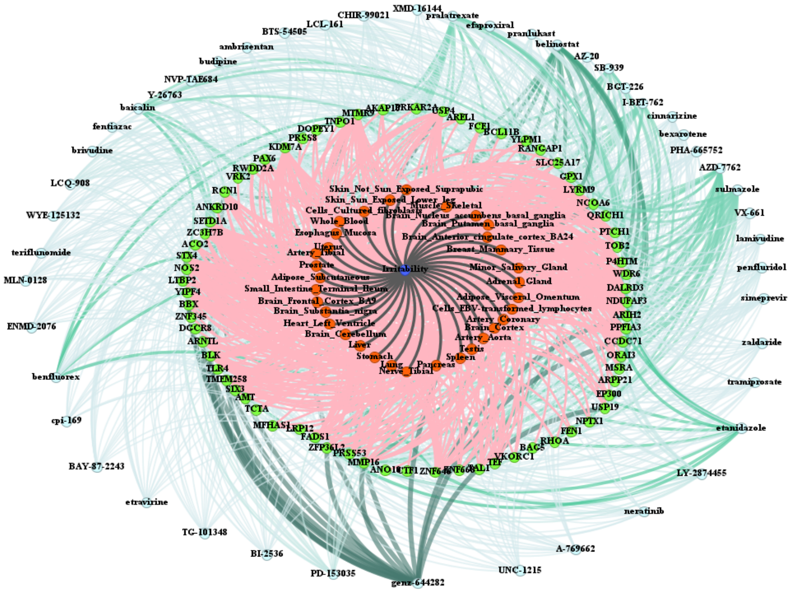

**Supplementary Figure S16: Irritability: Trait-Tissue-Gene-Drug Interaction Network.** This network visualizes the multi-level interactions among traits (from genetic research), tissues (with emphasis on significant tissues), genes and recycled drugs. Nodes represent four different biological entities:

**Trait**: Derived from genome-wide association studies (GWAS), indicating specific conditions or phenotypes related to genes. **Tissue**: represents the organ or tissue that is studying gene expression and drug interaction. **Gene**: Encoding a specific protein affected by genetic variation or regulated by drugs. **Drugs**: drugs that represent potential changes in use, targeting genes identified from genetic association.

Edges (connections) are weighted according to the importance of their association:

**Drug-gene edge**: weighted by negative log10 transformation arrangement P value $-log_{10}(p)$, indicating the association strength between drug and gene. **Tissue-gene edge:** fixed weight of 8, representing the established interaction within the tissue of interest.

Based on genetic and tissue-specific data, especially focusing on significant intra-tissue interactions, the network provides insights into the potential drug reuse of irritability.

S17: Worrier/Anxious Feelings: Trait-Tissue-Gene-Drug Interaction Network.

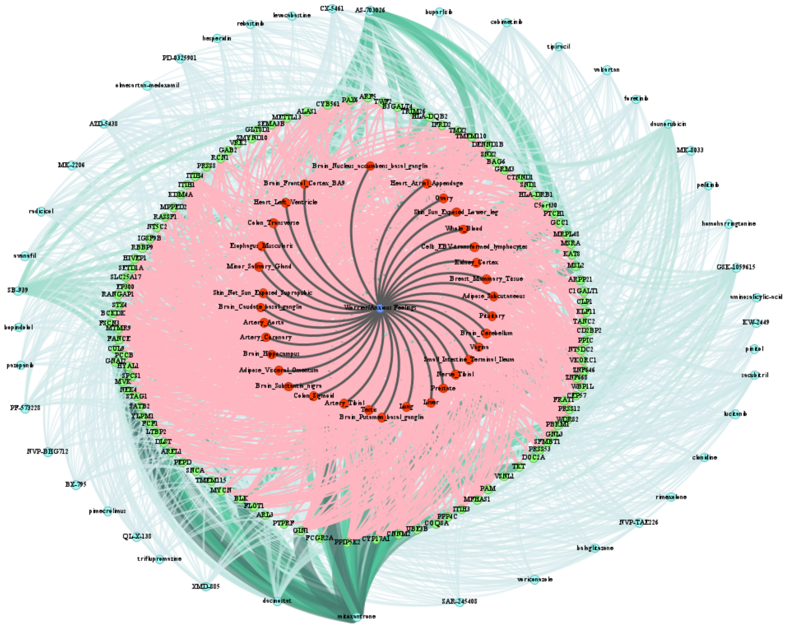

**Supplementary Figure S17: Worrier/Anxious Feelings: Trait-Tissue-Gene-Drug Interaction Network.** This network visualizes the multi-level interactions among traits (from genetic research), tissues (with emphasis on significant tissues), genes and recycled drugs. Nodes represent four different biological entities:

**Trait**: Derived from genome-wide association studies (GWAS), indicating specific conditions or phenotypes related to genes. **Tissue**: represents the organ or tissue that is studying gene expression and drug interaction. **Gene**: Encoding a specific protein affected by genetic variation or regulated by drugs. **Drugs**: drugs that represent potential changes in use, targeting genes identified from genetic association.

Edges (connections) are weighted according to the importance of their association:

**Drug-gene edge**: weighted by negative log10 transformation arrangement P value $-log_{10}(p)$, indicating the association strength between drug and gene. **Tissue-gene edge:** fixed weight of 8, representing the established interaction within the tissue of interest.

Based on genetic and tissue-specific data, especially focusing on significant intra-tissue interactions, the network provides insights into the potential drug reuse of worrier/anxious Feelings.

S18: Top 50 Repurposed Drugs for Alzheimer's Disease Based on P-value in Dorsolateral Prefrontal Cortex.

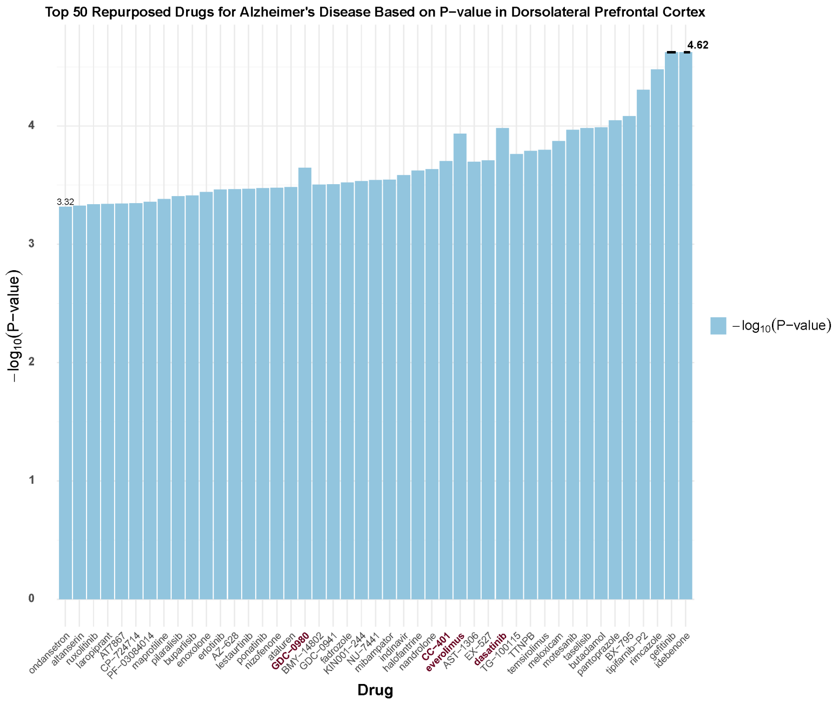

**Supplementary Figure S18: Top 50 Repurposed Drugs for Alzheimer's Disease Based on P-value in Dorsolateral Prefrontal Cortex.** We highlighted duplicate drugs using bold red font.

S19: Intersection Analysis of Shared Drugs and Tissues Between Irritability and Worrier/Anxious Feelings

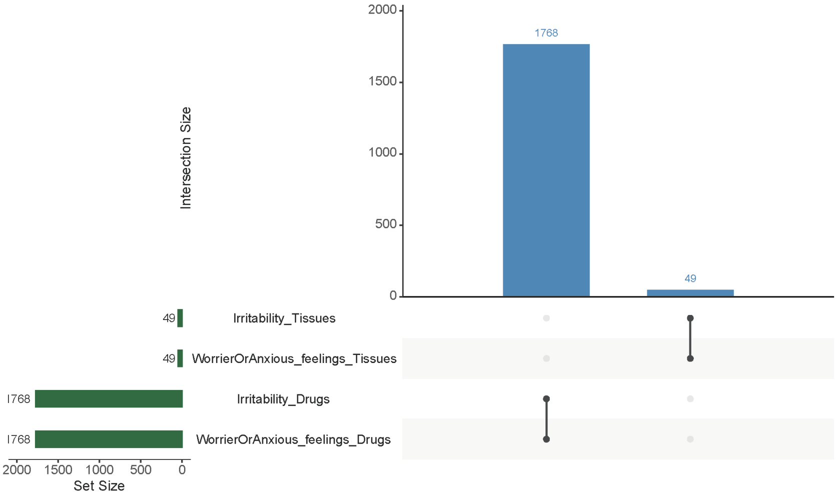

**Supplementary Figure S19: Intersection Analysis of Shared Drugs and Tissues Between Irritability and Worrier/Anxious Feelings.**

S20: Irritability: GO Biological Process and KEGG Pathway Enrichment Analysis Across Significant Tissues.

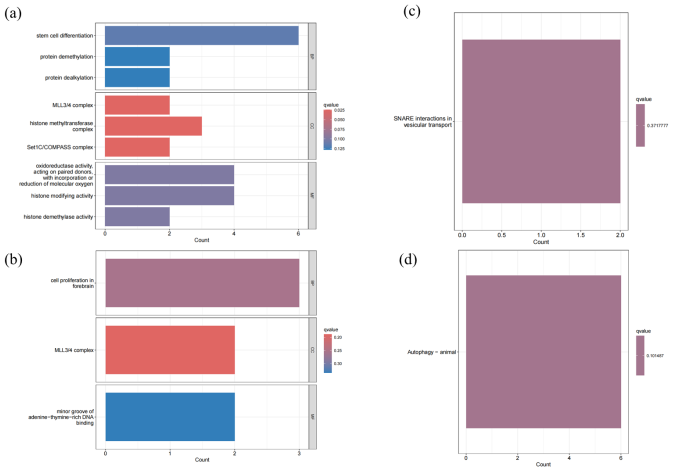

**Supplementary Figure S20:** Irritability: GO Biological Process and KEGG Pathway Enrichment Analysis Across Significant Tissues. (p-value and q-value ⩽ 0.05). a) GO biological process enrichment analysis in key tissues of interest. b) GO biological process enrichment analysis in other significant tissues. c) KEGG pathway enrichment analysis in key tissues of interest. d) GO biological process enrichment analysis in other significant tissues.

S21: Worrier/Anxious Feelings: GO Biological Process and KEGG Pathway Enrichment Analysis Across Significant Tissues.

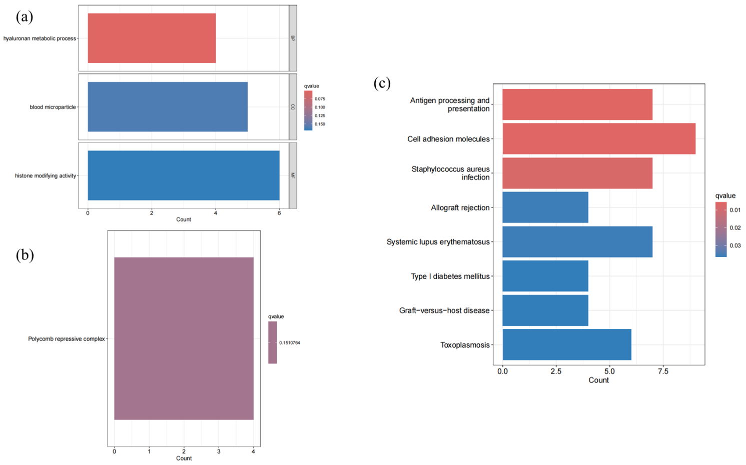

**Supplementary Figure S21:** Worrier/Anxious Feelings: GO Biological Process and KEGG Pathway Enrichment Analysis Across Significant Tissues. (p-value and q-value $\leqslant$ 0.05). a) GO biological process enrichment analysis in key tissues of interest. b) KEGG pathway enrichment analysis in key tissues of interest. d) KEGG pathway enrichment analysis in key tissues of interest.

S22: Worrier/Anxious Feelings: GO biological process enrichment analysis in other significant tissues.

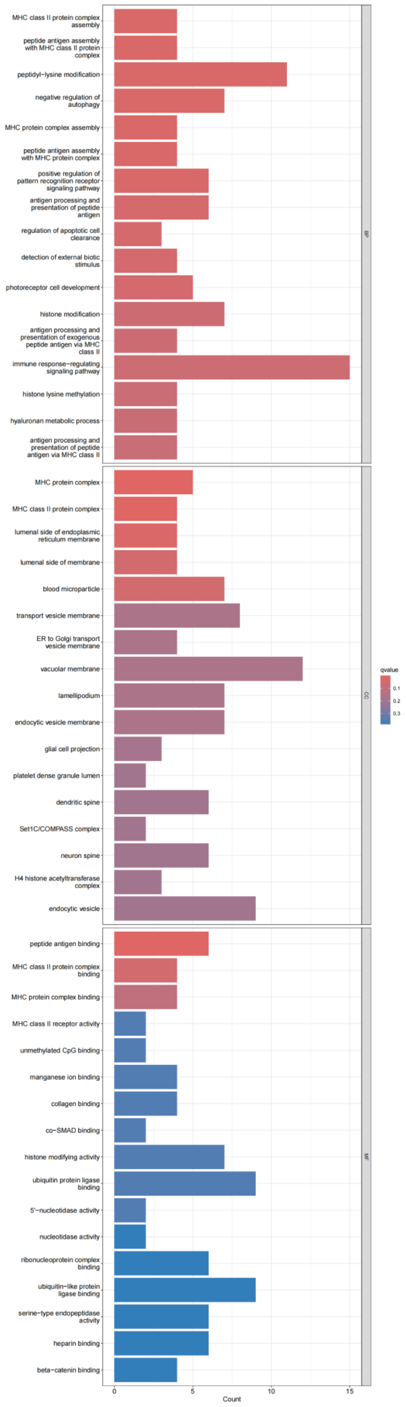
MHC class II protein complex assembly
peptide antigen assembly with MHC class II protein complex
peptidyl−lysine modification
negative regulation of autophagy
MHC protein complex assembly
peptide antigen assembly with MHC protein complex
positive regulation of pattern recognition receptor signaling pathway
antigen processing and presentation of peptide antigen
regulation of apoptotic cell clearance
detection of external biotic stimulus
photoreceptor cell development
histone modification
antigen processing and presentation of exogenous peptide antigen via MHC class II
immune response−regulating signaling pathway
histone lysine methylation
hyaluronan metabolic process
antigen processing and presentation of peptide antigen via MHC class II
BP
MHC protein complex
MHC class II protein complex
lumenal side of endoplasmic reticulum membrane
lumenal side of membrane
blood microparticle
transport vesicle membrane
ER to Golgi transport vesicle membrane
vacuolar membrane
lamellipodium
endocytic vesicle membrane
glial cell projection
platelet dense granule lumen
dendritic spine
Set1C/COMPASS complex
neuron spine
H4 histone acetyltransferase complex
endocytic vesicle
CC
peptide antigen binding
MHC class II protein complex binding
MHC protein complex binding
MHC class II receptor activity
unmethylated CpG binding
manganese ion binding
collagen binding
co−SMAD binding
histone modifying activity
ubiquitin protein ligase binding
5'−nucleotidase activity
nucleotidase activity
ribonucleoprotein complex binding
ubiquitin−like protein ligase binding
serine−type endopeptidase activity
heparin binding
beta−catenin binding
MF
0
5
10
15
Count
qvalue
0.1
0.2
0.3

**Supplementary Figure S22:** Worrier/Anxious Feelings: GO biological process enrichment analysis in other significant tissues.

S23: Worrier/Anxious Feelings: GO Biological Process and KEGG Pathway Enrichment Analysis in Brain Anterior Cingulate Cortex (BA24) and Artery Aorta.

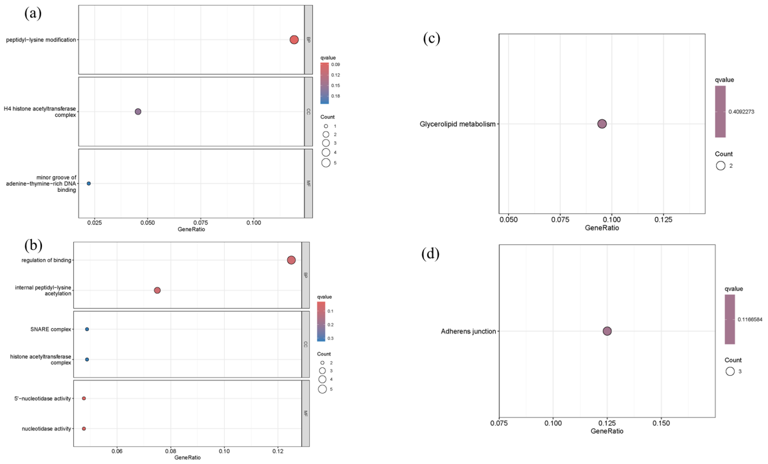

**Supplementary Figure S23:** Worrier/Anxious Feelings: GO Biological Process and KEGG Pathway Enrichment Analysis in Brain Anterior Cingulate Cortex (BA24) and Artery Aorta (q-value ⩽ 0.01). **a)** GO Biological Process Enrichment Analysis in Brain Anterior Cingulate Cortex (BA24). **b)** GO Biological Process Enrichment Analysis in Artery Aorta. **c)** KEGG Pathway Enrichment Analysis in Brain Anterior Cingulate Cortex (BA24). **d)** KEGG Pathway Enrichment Analysis in Artery Aorta.

S24: Irritability: GO Biological Process and KEGG Pathway Enrichment Analysis in Brain Anterior Cingulate Cortex (BA24) and Artery Aorta.

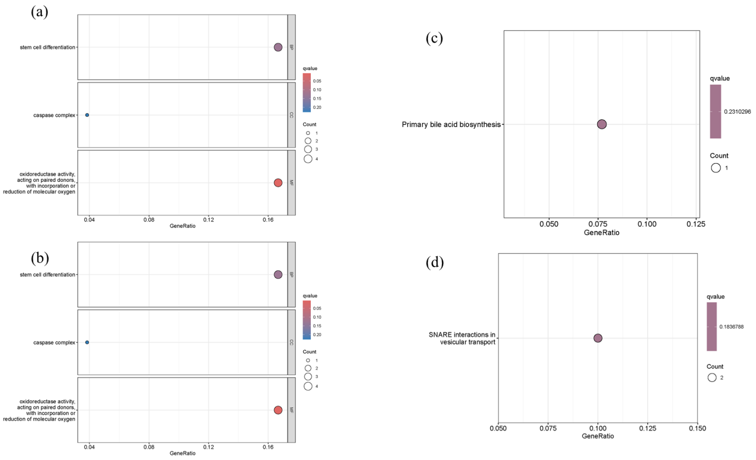

**Supplementary Figure S24:** Irritability: GO Biological Process and KEGG Pathway Enrichment Analysis in Brain Anterior Cingulate Cortex (BA24) and Artery Aorta (q-value ⩽ 0.01). **a)** GO Biological Process Enrichment Analysis in Brain Anterior Cingulate Cortex (BA24). **b)** GO Biological Process Enrichment Analysis in Artery Aorta. **c)** KEGG Pathway Enrichment Analysis in Brain Anterior Cingulate Cortex (BA24). **d)** KEGG Pathway Enrichment Analysis in Artery Aorta.

Supplementary Methods

AD-MIF Model Based on Information Fusion: Gene Feature Analysis and Prediction for Alzheimer's Disease.

#### 1.1 Overview of AD-MIF

The AD-MIF model consists of three components: the data preprocessing and gene neighborhood feature map construction module, the information fusion module, and the prediction module. Firstly, during the data preprocessing stage, the model preprocesses AD gene-phenotype data to address issues of high dimensionality and feature sparsity. Using Euclidean distance, the model identifies the K most similar features for each gene or SNP and constructs K-NN gene/SNP neighborhood feature maps (Gen-G/SNP-G). Subsequently, in the information fusion module, AD-MIF utilizes autoencoders and graph autoencoders to extract latent feature information, integrating specific and shared features to generate a highly consistent feature representation. Finally, the prediction module performs classification based on the processed features, making it suitable for various Alzheimer's disease-related applications.

#### 1.2 Genotype Data Preprocessing

Since the AD genotype data used involves multiple chromosomes, this results in challenges of high feature dimensionality and sparsity in the genotype data. To address these limitations, the experiment constructs multi-branch multilayer perceptrons tailored to different chromosomes to efficiently extract genotype data features $\mathrm{G}_{\mathrm{i}}^{\mathrm{P}}$:

$$\mathrm{G}_{\mathrm{i}}^{\mathrm{P}} = MLP_i\left(\mathrm{G}_{\mathrm{i}}^{\mathrm{P}}\right) \tag{1}$$

where $\mathrm{G}_{\mathrm{i}}^{\mathrm{P}}$ represents the initial genotype data on the $i$-th chromosome, and $MLP_i$ denotes the multilayer perceptron corresponding to the $i$-th chromosome. Finally, the processed genotype features are concatenated and fused to obtain high feature density AD genotype data $\mathrm{G}^{\mathrm{P}}$.

#### 1.3 Construction of Omics Neighborhood Feature Maps

For the various omics data in Alzheimer's disease (AD), multiple interaction relationships between gene neighborhoods and SNP domains are considered. In addition to the original gene and SNP feature matrices, the experiment also constructs gene neighborhood feature maps and SNP domain feature maps to capture the interaction relationships between omics data neighborhoods. For each omics feature, AD-MIF identifies the top K most similar genes/SNPs based on Euclidean distance to define the neighborhood, and then connects them within the neighborhood. Subsequently, the experiment constructs a K-Nearest Neighbors (K-NN) gene neighborhood feature map (Gen-G) and an SNP domain feature map (SNP-G) based on the identified neighborhood sets to represent the interaction networks of gene neighborhoods and SNP neighborhoods.

In Gen-G, nodes represent gene data, and edges represent neighborhood relationships between genes. In SNP-G, nodes represent genotype data, and edges represent neighborhood relationships between SNPs. This structure allows interaction information between genes/SNPs to be transmitted between nodes and edges, facilitating effective feature sharing and enhancement. Notably, the parameter K is treated as a training parameter and is dynamically adjusted based on the model's downstream tasks. This dynamic adjustment accommodates different application needs, enhances the understanding of gene interactions, and improves the model's predictive performance across various AD prediction tasks.

#### 1.4 Information Fusion Module

To better utilize both the raw features of omics data and their neighborhood features, AD-MIF introduces an information fusion module. This module aims to integrate the specific features and shared features learned from autoencoders at both local and global levels, thereby generating highly consistent feature representations. Taking Gen-G as an example, the experiment first employs Autoencoders (AE) and Graph Autoencoders (GAE) to extract latent feature information from the original gene features $\mathrm{G_o^i}$and the adjacency matrix $\mathrm{A^i}$ of the gene neighborhood feature map.

Specifically, for the $i$-th modality data, the original gene features are extracted using an autoencoder:

$$\mathrm{Z_{EC}^i} = f_{enc^i}(\mathrm{G^i}) \tag{2}$$

where $f_{enc^i}$ represents the autoencoder for the $i$-th modality, and $\mathrm{G^i}$ denotes the original gene features. The graph autoencoder adopts a symmetric structure and performs multiple reconstructions based on the feature matrix and adjacency matrix in

Gen-G. The neighborhood feature representation obtained from the $i$-th reconstruction is:

$$\mathrm{Z}^{\mathrm{i}}_{\mathrm{GEC}_{(l)}} = f_{g-enc^{i}}\left(\mathrm{Tanh}\left(\tilde{\mathrm{A}}^{\mathrm{i}}\mathrm{Z}^{\mathrm{i}}_{\mathrm{GEC}_{(l-1)}}\mathrm{W}_{\mathrm{l}}\right)\right) \tag{3}$$

where $f_{g-enc^{i}}$ represents the graph autoencoder for the $i$-th modality, $\tilde{\mathrm{A}}^{\mathrm{i}}$ denotes the normalized adjacency matrix of Gen-G for the iii-th modality, $\mathrm{W}_{\mathrm{l}}$ is the weight matrix of the l-th layer of the GAE, and $\tanh(\cdot)$ is the activation function.

To integrate the latent feature information learned by the aforementioned autoencoder and graph autoencoder, the experiment first combines these two representations through a linear combination to obtain more comprehensive latent information:

$$\mathrm{Z}^{\mathrm{i}}_{\mathrm{I}} = \alpha\mathrm{Z}^{\mathrm{i}}_{\mathrm{EC}} + (1-\alpha)\mathrm{Z}^{\mathrm{i}}_{\mathrm{GEC}} \tag{4}$$

where $\alpha$ is a learnable feature weight, initially set to 0.5. During training, $\alpha$ is automatically adjusted through gradient descent to dynamically allocate weights based on downstream tasks. Based on the updated latent features, the experiment further employs message passing to capture the gene neighborhood interaction feature information in the Gen-G graph, resulting in the local feature representation of gene neighborhood interactions:

$$\mathrm{Z}^{\mathrm{i}}_{\mathrm{L}} = \tilde{\mathrm{A}}^{\mathrm{i}}\mathrm{Z}^{\mathrm{i}}_{\mathrm{I}} \tag{5}$$

To better represent the correlations of gene interactions, the experiment introduces a feature correlation layer to calculate the global relationship scores of each gene relative to other genes:

$$\mathrm{S}^{i}_{n,m} = \frac{\exp\left(\mathrm{Z}^{\mathrm{i}}_{\mathrm{L}}\left(\mathrm{Z}^{\mathrm{i}}_{\mathrm{L}}\right)^{T}\right)_{n,m}}{\sum_{\mathrm{k}\in\mathrm{N(i)}}\exp\left(\mathrm{Z}^{\mathrm{i}}_{\mathrm{L}}\left(\mathrm{Z}^{\mathrm{i}}_{\mathrm{L}}\right)^{T}\right)_{n,k}} \tag{6}$$

Based on the local gene interaction representation $\mathrm{Z}^{\mathrm{i}}_{\mathrm{L}}$ and the global relationship scores $\mathrm{S}^{\mathrm{i}}$, the experiment further computes the global feature representation of gene interaction representation $\mathrm{Z}^{\mathrm{i}}_{\mathrm{G}}$, defined as follows:

$$\mathrm{Z}^{\mathrm{i}}_{\mathrm{G}} = \mathrm{S}^{\mathrm{i}}\mathrm{Z}^{\mathrm{i}}_{\mathrm{L}} \tag{7}$$

Finally, by introducing a feature weight parameter $\beta$ and performing element-wise summation between the local and global representations, the final fused representation is obtained:

$$Z^{i} = Z_{L}^{i} + \beta Z_{G}^{i} \tag{8}$$

For genotype data, the construction process of SNP-G is similar to the aforementioned procedure.

#### 1.5 Prediction Module

In the experiment, data obtained from different modalities are fused and processed using a constructed Artificial Neural Network (ANN) to make predictions for various downstream tasks. A binary cross-entropy loss function is employed to calculate the task loss:

$$\mathrm{Loss} = \frac{1}{\lceil N \rceil} \sum_{(i,j)\in N} \left( y_{(i-j)} \log(\hat{y}_{(i-j)}) + \left(1 - y_{(i-j)} \log(1 - \hat{y}_{(i-j)}\right)\right) \tag{9}$$

During the training and optimization phase, the experiment utilizes the Adam optimizer with a learning rate of 0.001 and sets the batch size to 128 for each epoch to ensure efficient and stable model training. The AD-MIF model undergoes 1,000 iterations of training, and the model parameters with the best performance are retained by monitoring the Area Under the Curve (AUC) and Mean Squared Error (MSE) metrics on the validation set.

The AD-MIF model is developed using the PyTorch framework and trained on an NVIDIA GeForce RTX 4090 GPU.
